# Clinically reported covert cerebrovascular disease and risk of neurological disease: a whole-population cohort of 367,988 people using natural language processing

**DOI:** 10.1101/2025.06.12.25329472

**Authors:** Matthew H Iveson, Mome Mukerjee, Emma M Davidson, Huayu Zhang, Laura Sherlock, Emily L Ball, Grant Mair, Alice Hosking, Heather Whalley, Michael T C Poon, Joanna M Wardlaw, David Kent, Richard Tobin, Claire Grover, Beatrice Alex, William N Whiteley

**Affiliations:** Centre for Clinical Brain Sciences, University of Edinburgh; Centre for Clinical Informatics, Usher Institute, University of Edinburgh; Nuffield Department of Primary Care Health Sciences, University of Oxford; UK Dementia Research Institute, University of Edinburgh; Graduate School of Biomedical Sciences, Tufts University; School of Informatics, University of Edinburgh; School of Literatures, Languages and Cultures, University of Edinburgh; Edinburgh Futures Institute, University of Edinburgh; Advanced Care Research Centre, University of Edinburgh; British Heart Foundation Data Science Centre, HDRUK, London

**Keywords:** Stroke, dementia, cerebrovascular disease, radiology reports, natural language processing

## Abstract

**Background:** The relevance of covert cerebrovascular disease (CCD) in practice is uncertain, in part because estimation of risk in whole clinical populations is difficult. To address this gap, we measured clinically reported CCD in a large clinical cohort using natural language processing (NLP) and estimated subsequent disease risk in linked health record data.

**Methods:** From all people with brain imaging in Scotland from 2010 to 2018, we selected people with no prior hospitalisation for neurological disease. Four phenotypes were identified with NLP of imaging reports: white matter hypoattenuation or hyperintensities (WMH), lacunes, cortical infarcts and cerebral atrophy. Hazard ratios (aHR) for stroke, dementia, and Parkinson’s disease (conditions previously associated with CCD), epilepsy (a brain-based control condition) and colorectal cancer (a non-brain control condition), adjusted for age, sex, deprivation, region, scan modality, and pre-scan healthcare, were calculated for each phenotype.

**Findings:** From 367,988 people with brain imaging and no history of neurological disease, 129,199 (35%) had ≥1 phenotype. For each phenotype, the aHR of any stroke was: WMH 1·4 (95%CI: 1·3–1·4), lacunes 1·6 (1·5–1·6), cortical infarct 1·8 (1·7–1·9), and cerebral atrophy 1·1 (1·0–1·1). The aHR of any dementia was: WMH 1·3 (1·3–1·3), lacunes 1·0 (0·9–1·0), cortical infarct 1·1 (1·1–1·2) and cerebral atrophy 1·7 (1·7–1·8). The aHR of Parkinson’s disease was, in people with a report of: WMH 1·1 (1·0–1·2), lacunes 1·1 (0·9–1·2), cortical infarct 0·7 (0·6–0·9) and cerebral atrophy 1·4 (1·3–1·5). The aHRs between CCD phenotypes and epilepsy and colorectal cancer overlapped the null.

**Interpretation:** NLP identified CCD and atrophy phenotypes from routine clinical image reports, and these had important associations with future stroke, dementia and Parkinson’s disease. Prevention of neurological disease in people with CCD should be a priority for healthcare providers and policymakers.

**Funding:** The Chief Scientist’s Office, the Medical Research Council, the Alzheimer’s Society, Health Data Research UK, the Wellcome Trust, Research Data Scotland, MQ – Transforming Mental Health, The Alan Turing Institute, the National Institute for Health Research, and the Stroke Association.

**Research in context:** *Evidence before this study:* Systematic reviews of magnetic resonance imaging (MRI) in cohort studies show that people with asymptomatic, covert cerebrovascular disease (CCD) have an increased risk of stroke, dementia and Parkinson’s disease. Covert brain infarcts, identified with natural language processing (NLP) show similar associations in one US-based insurance system. However, there is a lack of data on clinically-reported CCD from whole populations.

*Added value of this study:* This study used a validated NLP algorithm to identify CCD and cerebral atrophy from both MRI and computed tomography (CT) imaging reports generated during routine healthcare in >367K people in Scotland. It also distinguished between three CCD phenotypes – white matter hypoattenuation/hyperintensities, lacunes, cortical infarcts – and cerebral atrophy, and their associations with stroke and dementia and their subtypes. In adjusted models, we demonstrate higher risk of dementia (particularly Alzheimer’s disease) in people with atrophy, and higher risk of stroke in people with cortical infarcts. However, associations with an age-associated control outcome (colorectal cancer) were neutral, supporting a causal relationship. It also highlights differential associations between cerebral atrophy and dementia and cortical infarcts and stroke risk.

*Implications of all the available evidence:* CCD or atrophy on brain imaging reports in routine clinical practice is associated with a higher risk of stroke or dementia. Evidence is needed to support treatment strategies to reduce this risk. NLP can identify these important, otherwise uncoded, disease phenotypes, allowing research at scale into imaging-based biomarkers of dementia and stroke.

## Introduction

Covert cerebrovascular disease (CCD) is a common incidental finding after brain imaging. CCD imaging phenotypes include subcortical and periventricular white matter hypoattenuation and hyperintensities (WMH), small subcortical (lacunar) infarcts and large artery atherothrombo- or cardio-embolic (cortical) infarcts in people who have never reported symptoms of stroke or transient ischaemic attack (TIA). Systematic reviews of cohorts screened by magnetic resonance imaging (MRI) show that CCD prevalence is much higher than symptomatic stroke,^1–3^ and that findings of CCD are independently associated with stroke and dementia in population-based cohorts^4^. However, these findings may not generalise to clinical practice, because participants in cohort studies are not representative of people seeking healthcare, ^5^ and because neuroimaging reports generated during research are more standardised than clinical reports. Furthermore, while most studies identify CCD with MRI, computed tomography (CT) is more commonly used in clinical practice, particularly for those with minor symptoms for whom there is clinical uncertainty. Therefore, the clinical relevance of clinically reported CCD is often uncertain.

CCD does not have a diagnostic code in common classifications (e.g., International Classification of Diseases (ICD)-11), hence it is difficult to study in populations representative of clinical practice. Natural language processing (NLP) can help to identify hard to find phenotypes in medical notes and clinical reports. We have developed and validated a rules-based NLP algorithm (EdIE-R) with excellent recall and precision to identify brain imaging phenotypes from brain imaging radiology reports.^6^ This algorithm performed with high accuracy, compared with a reference standard of expert reads of brain imaging reports or brain images.^6–9^ Therefore NLP could reveal CCD and atrophy phenotypes in whole populations of patients who have had brain imaging during routine healthcare.

Here, we used NLP to identify clinically reported CCD and cerebral atrophy on CT and MRI brain imaging reports from routine healthcare in the Scottish population for up to 12 years after index imaging. We estimated the absolute risks and association of clinically reported CCD and atrophy with incident stroke, dementia, and Parkinson’s disease, and with epilepsy (a brain-based control condition not directly related to CCD) and colorectal cancer (a non-brain control condition).

## Methods

### Study design and participants

We used de-identified individual-level data, accessed through the National Health Service (NHS) Scotland’s National Safe Haven provided by Public Health Scotland (PHS). PHS deterministically linked general hospital inpatient records (Scottish morbidity record (SMR) 01), mental health inpatient records (SMR 04), community prescribing (Prescribing Information System (PIS)), cancer registrations (SMR 06), NRS death records and brain imaging reports from Scottish Medical Imaging with the community health index (CHI) number, a unique patient identifier, between 1st January 2008 and 31st December 2020.^10^

We extracted data from the whole population of Scotland with at least one NHS Scotland hospitalisation, outpatient or prescribing record (5,667,954 people). We selected all 785,331 people with a brain MRI or CT scan between January 2010 and August 2018. We excluded people with missing demographic data, an imaging date that conflicted with emigration or death, reports from research studies (e.g., clinical trials), reports with no usable information, and reports of non-brain imaging. We excluded people with records of stroke of any type, dementia, epilepsy, colorectal cancer, multiple sclerosis (MS), Parkinson’s disease, transient ischaemic attack (TIA), subarachnoid haemorrhage, subdural haemorrhage and extradural haemorrhage, prior to the date of scan or 6 months after the date of scan (Supplementary Material). We then removed individuals who died or transferred out within 6 months after the date of scan. The process of selecting the analytic cohort is shown in Figure 1.

**Figure 1.**
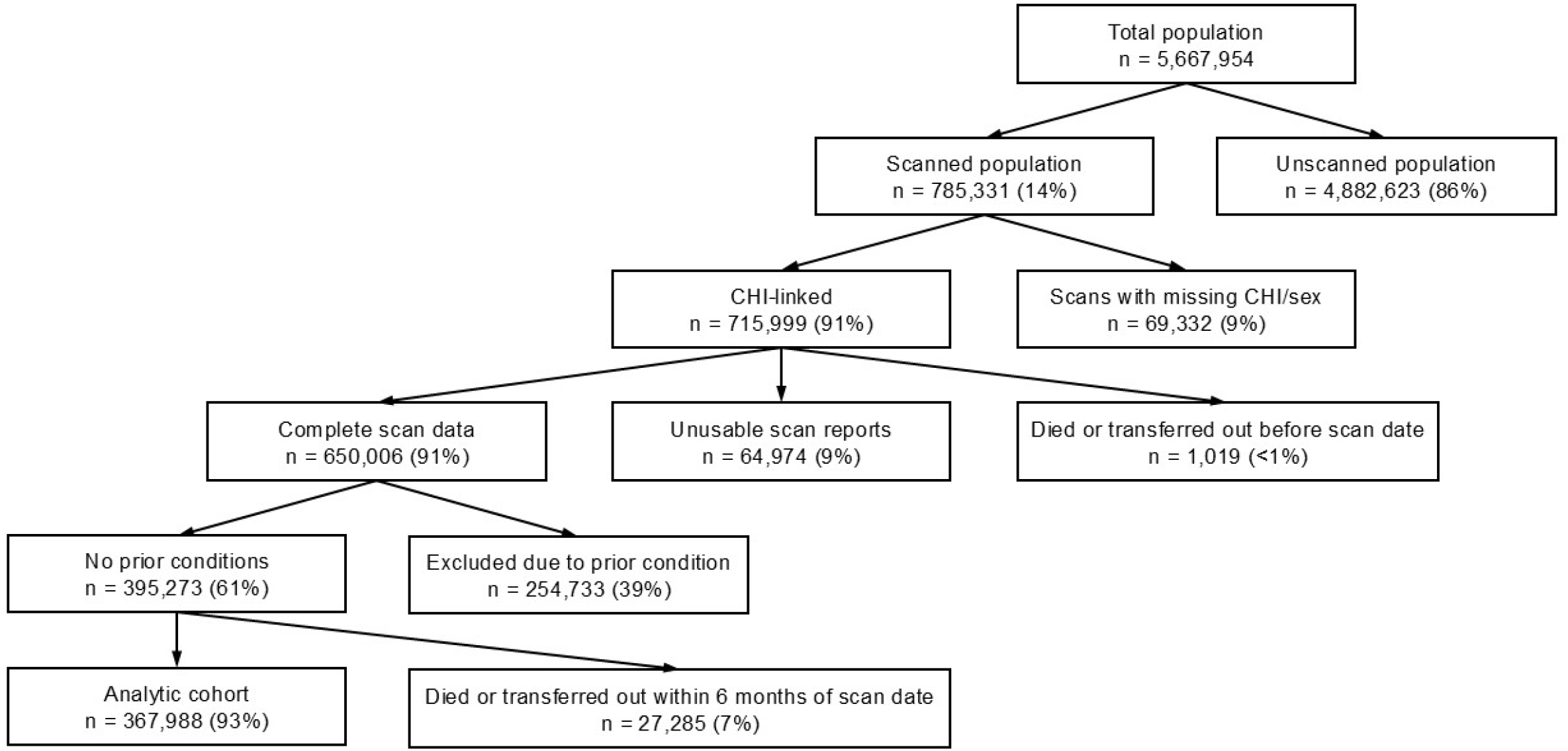
Flow diagram showing cohort selection from the whole population of Scotland who interacted with the Scottish NHS (N = 5,667,954) to the analytic cohort (N = 367,988). Percentages indicate the proportion of the node above. CHI = Community Health Index

### Study definitions

CCD and atrophy imaging phenotypes were white matter hypoattenuation (on CT) or hyperintensities (on MRI) (WMH), lacunes (small subcortical infarcts), cortical infarcts, and cerebral atrophy. Cerebral atrophy was included because it is a common co-finding of CCD and has potentially important disease associations. Rules-based NLP identified CCD or atrophy on first-available MRI or CT brain imaging report for each participant (https://www.ltg.ed.ac.uk/software/EdIE-R/). The tool has been validated against expert reads of Scottish brain imaging reports, and against an expert independent rater reading underlying images^6,9^ Additionally, we created a variable summing the number of CCD and atrophy phenotypes present at the first brain image.

We defined outcomes as the first recorded date of: any stroke (and subtypes ischaemic stroke, intracerebral haemorrhage and uncertain stroke); all dementia (and subtypes Alzheimer’s dementia, vascular dementia, and unspecified or rare dementia); Parkinson’s disease; epilepsy; and colorectal cancer. Epilepsy and colorectal cancer was included as brain-based and non-brain negative control outcomes, respectively. Outcomes were defined with established ICD10 coding and British National Formulary (BNF) medication lists (Supplementary Material) taken from the Health Data Research UK Phenotype Library (https://phenotypes.healthdatagateway.org/) with SMR01, SMR04, SMR06, PIS and national death records.

Covariates included age (in years) at first-available imaging report date, split into 5-year bands, sex, area-level measures of deprivation at first brain imaging (Scottish Index for Multiple Deprivation (SIMD) decile (2012 classification),^11^ Urban-Rural Classification (8-fold 2011 classification)^12^ and Health Board of scan (a proxy for region). Ethnicity data was not available for the cohort.

We used all data sources to define co-morbidities prior to first imaging: myocardial infarction (MI), angina, peripheral vascular disease, renal impairment, heart failure, non-colorectal cancer and diabetes (Supplementary Material). We measured healthcare usage in the year prior to first scan with the number of hospital admissions, the number of unique medicines dispensed, the number of statin prescriptions dispensed, and the number of antihypertensive prescriptions dispensed.

Follow-up started from 6 months after the date of the first brain imaging report (index date), to allow time for diagnosis made with or because of the first brain imaging to appear in records. For each outcome separately, survival time was estimated to an event, or right-censored on death, migration out of Scotland, or study end (31/12/2020). We report 1-year, 5-year, and overall cumulative incidence with the Kaplan-Meier method for each outcome by scan phenotypes (WMH, lacunes, cortical infarcts, cerebral atrophy), and survival estimates relative to the wider Scottish population (the analytic cohort plus the unscanned population alive on 01/07/2010; N = 4,637,231). Survival analyses were conducted with Cox proportional hazards models and the proportional hazards assumption was tested statistically and visually with Schoenfeld residual plots. The variables ‘scan modality’, and the Scottish Health Board violated this assumption and were included as stratification variables. For each outcome (and its subtypes), and for each CCD and atrophy phenotype we estimated maximally adjusted hazard ratios (aHRs), adjusting for other phenotypes, age, sex, deprivation, rurality, co-morbidities and healthcare usage measured by prescriptions and admissions in the year prior to scan. We also examined adjusted HRs for number of CCD and atrophy phenotypes at scan (max 4). Full model estimates, including univariate HRs, are shown in Supplementary Material.

Sensitivity analyses for each outcome (and subtype) tested whether diagnosis preceded scan by excluding the first 1 and 5 years of follow-up. While the first 6 months of follow-up are excluded in the primary analyses, excluding larger time periods provides a stricter test of potential asymptomatic time, particularly for conditions with more insidious onset. Subgroup analyses tested aHR by age, sex, and brain imaging type.

Analyses were conducted within the NHS Scotland National Safe Haven, using R (v4.3.2)^13^ and R Studio (v2023.12.0)^14^ with the package ‘survival’.^15^ Missing data were not imputed.

The study was approved by the NRES Committee Northwest - Greater Manchester East ethics committee (15/NW/0719), and the Public Benefit and Privacy Panel for Health and Social Care (1516-0219) of NHS Scotland.

## Results

Of 5·67 million people in Scotland between 2008 and 2018, 0·65 million (11%) had at least one brain imaging report, of whom 0·36 million (57%) had no recorded history of neurological disease and were alive 6 months after scan. Of this 0.36 million, most (54%) were over 55 years old, 22% had a report of WMH, 8% of a lacune, 2% of a cortical infarct, 22% of cerebral atrophy and 65% had none of these scan phenotypes. There was a total of 2·4-million-person years of follow-up time, with median follow-up time of people with CCD or atrophy of 5·0 years, and of scanned individuals without CCD or atrophy of 7·0 years (Supplementary Material).

Compared with people without, those with CCD or atrophy were older (mean 73·4 vs 48·2 years), had more admissions with MI (13·2% vs 3·8%), renal impairment (9·2% vs 1·9%), heart failure (5·1% vs 1·0%), cancer (10·7% vs 5·4%), and diabetes (9·7% vs 3·3%), had a greater number of hospitalisations (mean 1·1 vs 0·7) and prescribed medicines (3·1 vs 1·6) in the year prior to scan (Table 1). Compared with the Scottish population (with or without a brain imaging), people who had brain imaging were older, had more vascular and cancer comorbidities, and had fewer antihypertensive and statin dispensing events in the year prior to scan.

**Table 1.** Demographic characteristics of those with and without clinically reported covert cerebrovascular (CCD) or cerebral atrophy phenotype and of the whole Scottish population (analytic cohort plus unscanned individuals alive at 01/07/2010).

|  |  | Analytic cohort |  |  | Scottish population |
| --- | --- | --- | --- | --- | --- |
|  |  | Covert cerebrovascular disease or cerebral atrophy<br>n= 129,199 | No covert cerebrovascular disease or cerebral atrophy<br>n= 238,789 | All<br>N = 367,988 | N = 4,637,231 |
| **Sex** | Female | 69,246 (53.6%) | 129,957 (54.4%) | 199,203 (54.1%) | 2,457,323 (53.0%) |
| **Age (years)** | Mean (SD) | 73.4 (12.6) | 48.2 (17.1) | 57.0 (19.8) | 40.9 (22.5) |
| **Age group** | <=25 | 324 (0.3%) | 26,526 (11.1%) | 26,850 (7.3%) | 613,158 (13.2%) |
|  | 26-35 | 896 (0.7%) | 38,404 (16.0%) | 39,300 (10.7%) | 616,072 (13.3%) |
|  | 36-45 | 2,675 (2.1%) | 41,770 (17.5%) | 44,445 (12.1%) | 684,817 (14.8%) |
|  | 46-55 | 8,395 (6.5%) | 49,818 (20.8%) | 58,213 (15.8%) | 703,974 (15.2%) |
|  | 56-65 | 17,788 (13.8%) | 40,537 (17.0%) | 58,325 (15.9%) | 589,770 (12.7%) |
|  | 66-75 | 34,841 (27.0%) | 26,853 (11.2%) | 61,694 (16.8%) | 400,205 (8.6%) |
|  | 76-85 | 44,257 (34.2%) | 12,051 (5.1%) | 56,308 (15.3%) | 227,668 (4.9%) |
|  | 86-95 | 18,958 (14.7%) | 2,711 (1.2%) | 21,669 (5.9%) | 77,073 (1.7%) |
|  | >=96 | 1,065 (0.8%) | 119 (<0.1%) | 1,184 (0.3%) | 6,638 (0.1%) |
| **Scan modality** | CT | 105,977 (82.0%) | 167,623 (70.2%) | 273,600 (74.4%) | - |
|  | MR | 23,222 (18.0%) | 71,166 (29.8%) | 94,388 (25.6%) | - |
| **SIMD\* quintile** | 1 Most deprived | 29,800 (23.1%) | 58,365 (24.4%) | 88,165 (24.0%) | - |
|  | 2 | 26,887 (20.8%) | 48,793 (20.4%) | 75,680 (20.6%) | - |
|  | 3 | 23,863 (18.5%) | 44,398 (18.6%) | 68,261 (18.5%) | - |
|  | 4 | 23,043 (17.8%) | 41,734 (17.5%) | 64,777 (17.6%) | - |
|  | 5 Least deprived | 23,204 (18.0%) | 38,449 (16.1%) | 61,653 (16.8%) | - |
|  | Missing | 2,402 (1.8%) | 7,050 (3.0%) | 9,452 (2.5%) | - |
| **Urban-Rural\*\*** | 1 Large Urban | 53,271 (41.2%) | 96,467 (40.4%) | 149,738 (40.7%) | - |
|  | 2 | 38,468 (29.8%) | 70,018 (29.3%) | 108,486 (29.5%) | - |
|  | 3 | 10,649 (8.3%) | 18,752 (7.9%) | 29,401 (8.0%) | - |
|  | 4 | 3,100 (2.4%) | 4,878 (2.0%) | 7,978 (2.2%) | - |
|  | 5 | 1,127 (0.9%) | 2,210 (0.9%) | 3,337 (0.9%) | - |
|  | 6 | 12,831 (9.9%) | 24,106 (10.1%) | 36,937 (10.0%) | - |
|  | 7 | 3,669 (2.8%) | 6,427 (2.7%) | 10,096 (2.7%) | - |
|  | 8 Very Remote Rural | 2,928 (2.3%) | 5,443 (2.3%) | 8,371 (2.3%) | - |
|  | Missing | 3,156 (2.4%) | 10,488 (4.4%) | 13,644 (3.7%) | - |
| **Comorbidities** | MI, angina, or peripheral vascular disease | 17,085 (13.2%) | 8,978 (3.8%) | 26,063 (7.1%) | 81,427 (1.8%) |
|  | Renal impairment | 11,922 (9.2%) | 4,555 (1.9%) | 16,477 (4.5%) | 35,510 (0.8%) |
|  | Heart failure | 6,639 (5.1%) | 2,283 (1.0%) | 8,922 (3.2%) | 25,586 (0.6%) |
|  | Any cancer | 13,886 (10.7%) | 13,000 (5.4%) | 26,886 (7.3%) | 84,033 (1.8%) |
|  | Diabetes | 12,511 (9.7%) | 7,916 (3.3%) | 20,427 (5.6%) | 58,600 (1.3%) |
| **Care in year before scan** | Hospital admissions (mean, SD) | 1.1 (2.4) | 0.7 (1.9) | 0.8 (2.1) | 0.8 (1.8) |
|  | Unique medicines (mean, SD) | 3.1 (2.7) | 1.6 (2.1) | 2.1 (2.4) | 2.8 (2.5) |
|  | Antihypertensive dispensing events (mean, SD) | 2.3 (3.6) | 0.8 (2.3) | 1.3 (2.9) | 3.7 (6.9) |
|  | Statin dispensing events (mean, SD) | 1.0 (1.6) | 0.3 (1.0) | 0.6 (1.3) | 1.5 (3.1) |

\*2012 \*\*2011-2012

The five most frequent combinations of CCD and atrophy phenotypes were: cerebral atrophy alone; WMH and cerebral atrophy; WMH alone; WMH, cerebral atrophy and lacunes; and lacunes. The proportion of people who had an MRI rather than a CT was higher in people with no phenotype and for WMH only, although for all phenotype combinations, most brain imaging was performed with CT (Figure 2).

**Figure 2.**
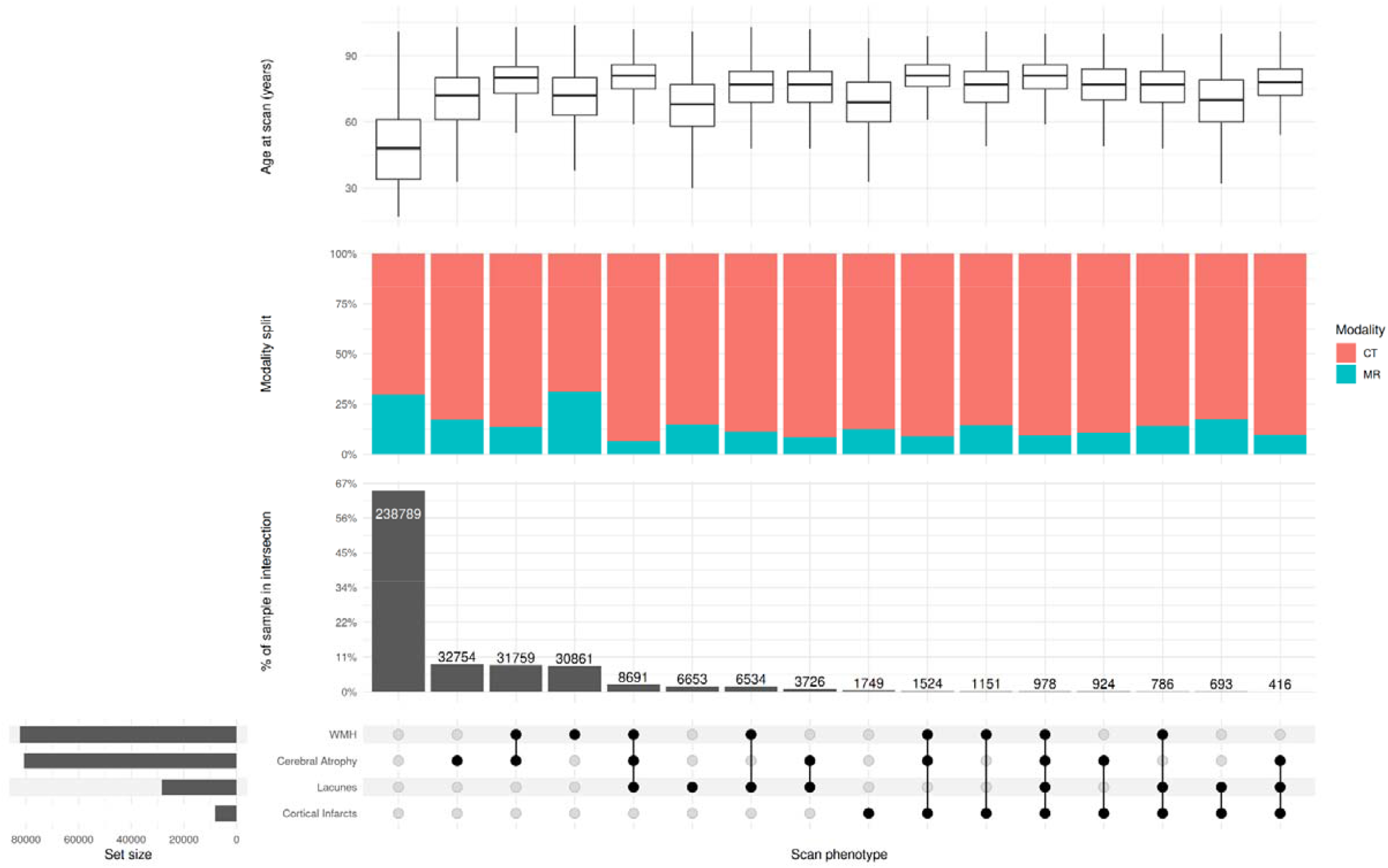
Co-occurrence of covert cerebrovascular disease phenotypes, with summary of imaging modality and age at imaging per intersection. Box and whisker plots represent range, interquartile range and median. WMH = white matter hypoattenuation/hyperintensities

The incidence rate, absolute risk difference, and relative risk (RR) of outcomes between people with a brain image report and the wider Scottish population are shown in Supplementary Material. Incidence was proportionally higher in those with a brain image report than in the wider population for all outcomes except colorectal cancer, though RR were lower with increasing age (Supplementary Material). At one and five years, people with a CCD or atrophy phenotype were more likely than the wider Scottish population to experience stroke (1-year: RR 6·4, 95%CI: 6·1–6·7; 5-year: RR 30·3, 29·6–31·0), dementia (5·1, 5·0–5·2; 16·8, 16·6–17·0), Parkinson’s disease (3·5, 3·3–3·8; 18·1, 17·4–18·9) and epilepsy (1·9, 1·8–1·9; 7·9, 7·8–7·9) but not colorectal cancer (1·1, 0·9–1·2; 5·1, 4·9–5·3). At one year, the cumulative incidence (accounting for the competing risk of death) of stroke and dementia were similar and low among those with WMH, lacunes, cortical infarcts, cerebral atrophy or no scan phenotype (around 2% and 9%; Table 2). At five years, cumulative stroke incidence was highest among those with cortical infarcts (11%), followed by lacunes (9%), WMH (7%) and cerebral atrophy (6%), while dementia incidence was highest among those with cerebral atrophy (26%), followed by WMH (23%), cortical infarcts (22%) and lacunes (22%).

**Table 2.** 1-year and 5-year cumulative incidence (%) for each scan phenotype and outcome, accounting for competing risk of death.

|  | WMH |  | Lacunes |  | Cortical infarcts |  | Cerebral atrophy |  | None |  | Analytic cohort |  | Scottish population |  |
| --- | --- | --- | --- | --- | --- | --- | --- | --- | --- | --- | --- | --- | --- | --- |
| Outcome | 1-year | 5-year | 1-year | 5-year | 1-year | 5-year | 1-year | 5-year | 1-year | 5-year | 1-year | 5-year | 1-year | 5-year |
| Stroke | 2.0<br>[2.0-2.1] | 6.8<br>[6.6-7.0] | 2.6<br>[2.4-2.8] | 8.7<br>[8.4-9.0] | 3.5<br>[3.1-3.9] | 11.0<br>[10.0-12.0] | 1.8<br>[1.7-1.9] | 6.2<br>[6.0-6.4] | 0.3<br>[0.3-0.3] | 1.2<br>[1.1-1.2] | 0.8<br>[0.8-0.8] | 2.9<br>[2.9-3.0] | 0.1<br>[0.1-0.1] | 0.2<br>[0.2-0.2] |
| Dementia | 9.6<br>[9.4-9.8] | 23.0<br>[23.0-24.0] | 8.4<br>[8.1-8.7] | 22.0<br>[21.0-22.0] | 9.1<br>[8.5-9.8] | 22.0<br>[22.0-23.0] | 11.0<br>[11.0-11.0] | 26.0<br>[25.0-26.0] | 0.8<br>[0.8-0.9] | 2.3<br>[2.2-2.3] | 3.7<br>[3.6-3.8] | 9.5<br>[9.4-9.6] | 0.5<br>[0.5-0.5] | 1.0<br>[1.0-1.0] |
| Parkinson's | 0.5<br>[0.5-0.6] | 1.9<br>[1.8-2.0] | 0.5<br>[0.5-0.6] | 1.9<br>[1.8-2.1] | 0.3<br>[0.2-0.5] | 1.4<br>[1.1-1.7] | 0.6<br>[0.5-0.6] | 2.2<br>[2.1-2.3] | 0.1<br>[<0.1-0.1] | 0.4<br>[0.4-0.4] | 0.2<br>[0.2-0.3] | 1.0<br>[1.0-1.0] | 0.1<br>[<0.1-0.1] | 0.1<br>[0.1-0.1] |
| Epilepsy | 3.2<br>[3.1-3.3] | 9.2<br>[9.0-9.4] | 3.2<br>[3.0-3.4] | 9.2<br>[8.9-9.6] | 3.4<br>[3.0-3.8] | 9.4<br>[8.8-10.0] | 3.2<br>[3.0-3.3] | 9.4<br>[9.2-9.6] | 4.1<br>[4.0-4.1] | 12.0<br>[12.0-13.0] | 3.9<br>[3.8-4.0] | 12.5<br>[12.4-12.6] | 2.0<br>[2.0-2.0] | 5.0<br>[5.0-5.0] |
| Colorectal Cancer | 0.3<br>[0.3-0.3] | 1.0<br>[0.9-1.1] | 0.3<br>[0.3-0.4] | 1.1<br>[0.9-1.2] | 0.4<br>[0.3-0.5] | 1.2<br>[0.9-1.4] | 0.3<br>[0.3-0.4] | 1.1<br>[1.0-1.2] | 0.1<br>[<0.1-0.1] | 0.4<br>[0.3-0.4] | 0.2<br>[0.2-0.2] | 0.6<br>[0.6-0.7] | 0.2<br>[0.2-0.2] | 0.5<br>[0.5-0.5] |

WMH = white matter hypoattenuation/hyperintensities

Compared to those with no CCD or atrophy phenotype, those with each phenotype (accounting for the presence of other phenotypes) had a higher risk of any stroke over the 12-year follow-up period. The aHRs were, for cortical infarct 1·8 (95% CI: 1·7–1·9), lacunes 1·6 (1·5–1·6), WMH 1·4 (1·3–1·4), and cerebral atrophy 1·1 (1·0–1·1) (Figure 3). The aHR for ischaemic stroke was highest in people with cortical infarct (1·9, 1·7–2·0) and aHR for intracerebral haemorrhage was highest in people with cortical infarct (1·7, 1·5–1·9) and lacunes (1·6, 1·5–1·7). People with cerebral atrophy had an increased risk of unspecified stroke (1·2, 1·1–1·3), but not ischaemic stroke (1·0, 0·9–1·1) or haemorrhagic stroke (1·1, 0·9–1·1) (Figure 4). People with more CCD or atrophy phenotypes had a higher aHR for all stroke (1·4, 1·3–1·4), with the highest aHR (versus those without phenotypes) in those with all four phenotypes (3·0, 2·5–3·6) (Supplemental Material).

**Figure 3.**
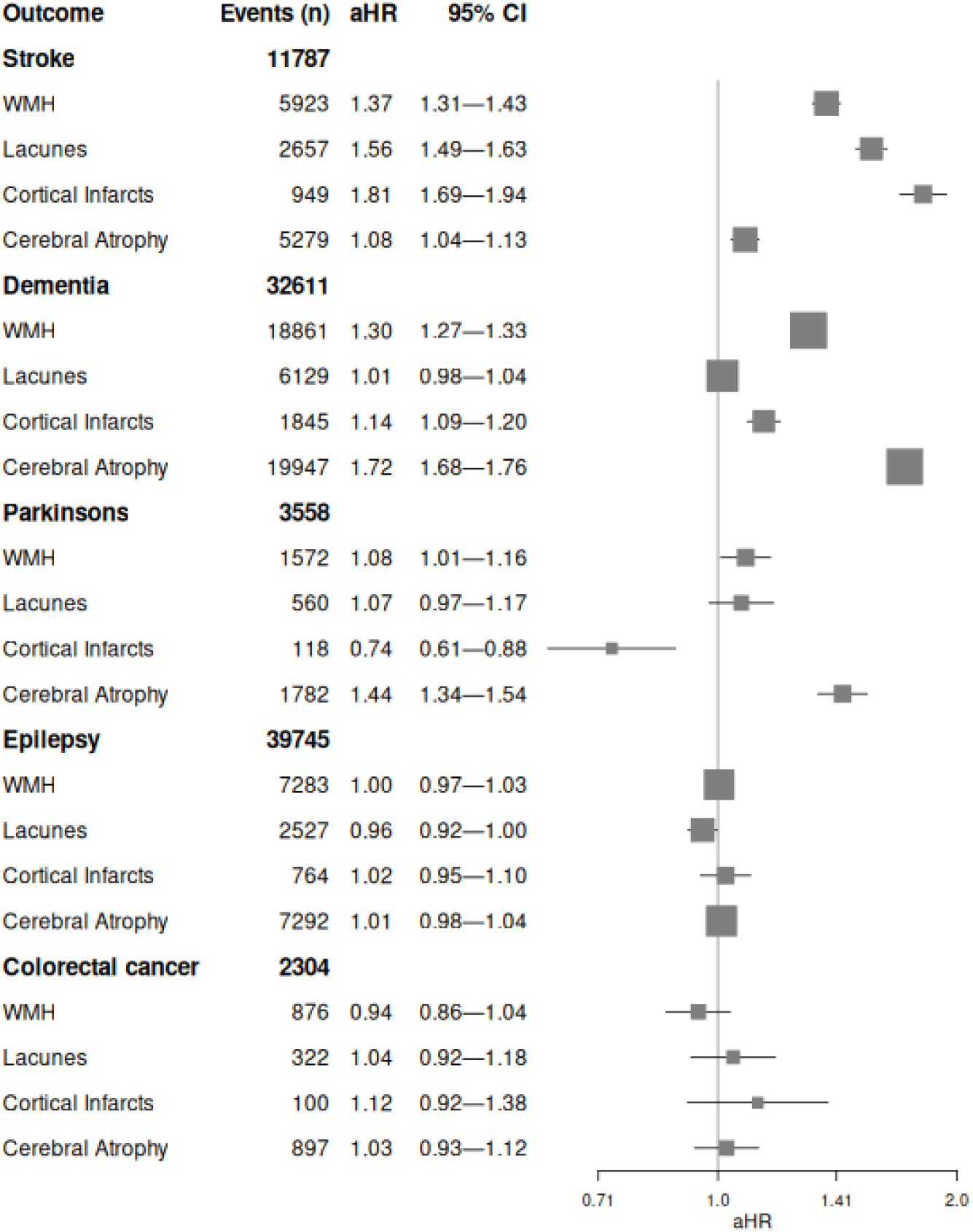
Forest plot showing the event rate and adjusted hazard ratio (aHR) and 95% confidence intervals for each exposure (white matter hyper (MRI) or hypointensity (CT) (WMH), lacunes, cortical infarcts and cerebral atrophy) and their association with stroke risk, dementia risk, Parkinson’s risk, epilepsy risk and colorectal cancer risk across follow-up.

**Figure 4.**
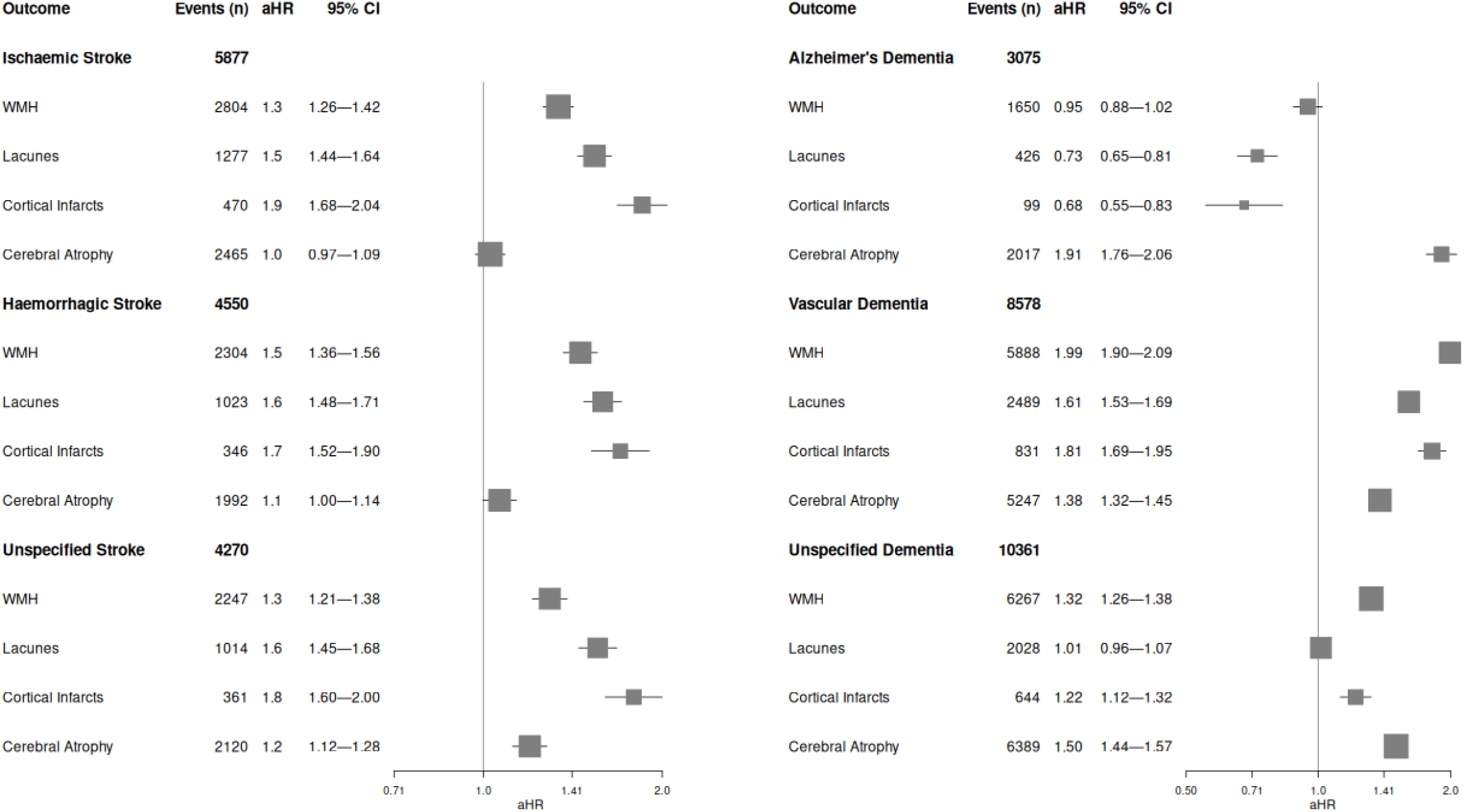
Forest plot showing the event rate and adjusted hazard ratio (aHR) and 95% confidence intervals for each exposure (white matter hyper (MRI) or hypointensity (CT) (WMH), lacunes, cortical infarcts, and cerebral atrophy) and their association with risk of a) stroke subtypes and b) dementia subtypes over follow-up.

Compared with people with no evidence of CCD or atrophy, the aHR for dementia was higher in people with cerebral atrophy (aHR 1·7, 95%CI: 1·7–1·8), WMH (1·3, 1·3–1·3), and cortical infarct at scan (1·1, 1·1–1·2), although not lacunes (1·0, 1·0–1·0) (Figure 3). Of the CCD or atrophy phenotypes, only people with cerebral atrophy at scan had an increased risk of each dementia type: Alzheimer’s disease (1·9, 1·8–2·1), vascular dementia (1·4, 1·3–1·4), and unspecified dementia (1·5, 1·4–1·6). Compared to those without CCD or atrophy phenotypes, people with cortical infarct or lacunes had a higher risk of vascular dementia (cortical infarct: 1·8, 1·7–2·0; lacunes: 1·6, 1·5–1·7) but lower incidence of Alzheimer’s disease (cortical infarct: 0·7, 0·6–0·8; lacunes: 0·7, 0·7–0·8). Compared to those without WMH, people with WMH had a higher risk of vascular dementia (2·0, 1·9–2·1) and unspecified dementia (1·3, 1·3–1·4), but not clearly Alzheimer’s dementia (0·9, 0·9–1·0) (Figure 4). People with more CCD phenotypes had a higher dementia aHR (1·3, 1·3–1·3), with the highest aHR among those exhibiting all four phenotypes (2·6, 2·3–2·9).

Compared with people with no evidence of CCD or atrophy, the aHR of Parkinson’s disease was lower in people with cortical infarct (aHR 0·7, 95%CI: 0·6–0·9) and higher in people with cerebral atrophy (1·4, 1·3–1·5) and WMH (1·1, 1·0–1·2), but not clearly higher in people with lacunes (1·1, 1·0–1·2) (Figure 3). There was a linear association between more scan phenotypes and Parkinson’s disease risk (1·2, 1·1–1·2), but risk in those with all four phenotypes did not significantly differ to those with no phenotypes (1·0, 0·6–1·7). There was a significant association between the presence of lacunes at scan and lower risk of epilepsy, though this association was notable weak (1·0, 0·9–1·0). None of the other CCD or atrophy phenotypes had a large aHR with risk of epilepsy (aHRs 0·9–1·0) or colorectal cancer (aHRs 0·9–1·1), and all had a p-value of >0.05.

### Sensitivity analyses

After excluding the first year and five years of follow up, most estimates were of a similar magnitude and direction to the primary analyses. The exceptions were, after excluding five years of follow up, the aHR for cerebral atrophy with stroke risk was around the null and aHRs between lacunes and cortical infarcts with dementia were <1 (Supplementary Material), indicating that stroke and dementia are diagnosed earlier among these individuals.

### Subgroup analyses

We tested subgroups by sex, age group, and scan modality. The aHR between stroke and WMH was higher in males than females (p_interaction_ < 0·001) and the aHRs between stroke and both cortical infarcts and cerebral atrophy were higher in females than males (p_interaction_ <0·01 and p_interaction_ < 0·001). The aHRs for WMH, lacunes, atrophy (all p_interaction_ < 0·001) and cortical infarcts (p_interaction_ < 0·05) were stronger in younger than older people, and aHRs for lacunes (p_interaction_ < 0·01) and cerebral atrophy (p_interaction_ < 0·001) were stronger with MRI than CT imaging. For dementia, aHRs for cerebral atrophy (p_interaction_ < 0·01) and WMH (p_interaction_ < 0·05) were stronger in males than females, aHRs for WMH, cerebral atrophy (p_interaction_ < 0·001) and cortical infarcts (p_interaction_ < 0·01) were stronger in younger than older people, and aHRs for cerebral atrophy were stronger for MRI than CT (p_interaction_ < 0·001) (Supplementary Material).

## Discussion

This cohort study of 367,988 people demonstrates that those with clinically reported CCD or cerebral atrophy have a higher incidence of future stroke and dementia diagnosis. Compared with people with normal scans, those with cortical infarct had the highest aHR of ischaemic stroke, and those with lacunes had the highest aHR of haemorrhagic stroke. Those with cerebral atrophy had the highest aHR of Alzheimer’s disease, and those with WMH the highest aHR of vascular dementia. Compared with the general population (and those with normal scans), the absolute incidence of dementia and stroke was substantially elevated in people with these scan reports.

As expected, the prevalence of CCD and atrophy phenotypes in this clinical cohort was higher than in research cohorts, where MRI-discovered WMH was present in ∼10% and silent brain infarcts in around ∼2% of 50–69 year-olds (though prevalence increases with age),^1^ compared with 22% and 10% in our study. The aHRs of WMH and deep brain infarct with stroke and dementia in research cohort studies were also stronger (aHR>2) than the maximally adjusted estimates from the present study.^4^ However, the aHRS reported here are similar to clinical studies for both stroke (aHRs 1·07–1·45)^16^ and dementia (aHRs 1·28– 1·93).^17^ Lower aHRs in clinical than research cohort studies could be due to lack of precision in measurement of brain imaging phenotypes in clinical compared with research studies, or higher risk of death in people in clinical studies hence lower measured disease incidence. Overall, the observed aHRs between cortical infarcts and all stroke risk (1·8) and between cerebral atrophy and all dementia risk (1·7) are concordant with previous studies in non-stroke and post-stroke populations, consistent with the view that stroke involves more discrete vascular lesions while dementia involves more brain tissue loss.

Our study had several strengths. First, a validated rules-based NLP pipeline was used to detect markers of CCD and atrophy in clinical radiology reports. Text radiology reports are ideally suited for high-performance NLP, due to their replicable structure, limited phenotypes and short length. Previous studies have not measured multiple phenotypes, have used smaller cohorts (Ns of 261,960 and 241,050), and only examined older people.^16,17^ Second, we took a whole clinical population approach, including both academic centres, where almost all research is currently done, and district hospitals, which do not usually contribute to research. Very few studies of imaging phenotypes have sample sizes of more than a thousand, and one of the largest cohorts, UK Biobank, is aiming for imaging one hundred thousand participants.^18^ The large sample in the present study had sufficient statistical power to study subgroups and rare conditions such as Parkinson’s disease. Third, linking participants to health systems data allowed ascertainment of multiple outcomes at low cost.^19–21^ Fourth, we included an ageing-related non-brain condition (colorectal cancer) that was not associated with CCD or atrophy, increasing confidence that the findings were not entirely due to residual confounding.

Our study had several limitations. First, rules-based NLP models are not state-of-the-art in the field relative to large language models (LLM). However, rules-based models tend to perform well,^22^ and the time taken for data governance approvals and limited compute resource meant that an LLM approach was not feasible when the project started. Second, our population was clinical, hence imaging was only performed in people at higher risk; this selection bias could limit generalisability to research cohorts or asymptomatic screening. Furthermore, imaging request information was too brief to meaningfully classify reports by indication, therefore stroke or dementia as a reason for imaging request could not be entirely excluded. However, we excluded people with a diagnosis of stroke or dementia prior to, or within 6 months of scanning and in sensitivity analysis found that extending this exclusion to one and five years made little qualitative difference to the pattern of aHRs. Third, we were only able to link to hospital, prescription, and death data, and did not have access to data from primary care, so some chronic conditions will have been under ascertained. This also meant that important confounders, such as obesity or smoking, were not available, and residual confounding could explain some of our findings. Fourth, there may have been diagnosis inclusion bias for dementia subtypes, because evidence of CCD could have increased the chance of a clinical diagnosis of vascular rather than other dementia subtypes. Lastly, our imaging phenotypes were relatively crude and did not include quantification or location. However, UK clinical brain imaging reports generally do not have sufficient detail to estimate the severity of WMH or atrophy, or precise location of deep or cortical infarcts. The appearance of old deep infarcts (‘lacunes’) is similar to perivascular spaces, recent small subcortical infarcts, small subcortical haemorrhage, or end-stage cavitation in a white matter hyperintensity, and is often difficult to distinguish in a clinical image. Ongoing work analysing the images in this dataset will provide quantitative phenotyping.

We have demonstrated that NLP can uncover previously difficult to discover phenotypes in electronic health records at a scale and speed not previously possible. However, care is needed as NLP can reflect systematic biases in the clinical text, for example incompleteness in reports in acute illness, and trends in language used in reports over time.^23–27^ NLP phenotypes could be used to identify non-coded outcomes in large cohort studies that rely on the electronic health record for outcome ascertainment, or to identify populations eligible for clinical trials.^28^ This may be true particularly for asymptomatic, incidentally discovered findings such as CCD.

This study emphasises that CCD or atrophy discovered during the process of routine healthcare are risk markers for stroke and dementia. In a clinical population, a report of atrophy indicates a modestly increased risk of subsequent dementia but does not make a clinical diagnosis, hence further assessment with cognitive testing may be needed. Apparently asymptomatic cortical infarcts or lacunes indicate a modestly increased risk of subsequent stroke, and in the absence of evidence to support specific interventions, careful application of primary vascular prevention – chiefly management of hypertension and high LDL cholesterol – should be the mainstay of treatment. Recent guidelines from the European Stroke Organisation (ESO) state that the best practice to prevent stroke or dementia in people with CCD remains uncertain.^29^

In conclusion, we implemented an NLP algorithm across a whole population and found that CCD and atrophy were associated with a higher risk of both dementia and stroke up to 12 years later, with important differences between CCD exposures. Clinical trials to reduce risk in people with CCD are warranted.

## Supporting information

RECORD checklist

## Data Availability

Data is restricted as per Public Health Scotland guidelines, as it relates to individual health records. Data can be made available within the National Safe Haven upon application and approval by Public Health Scotland and the Public Benefit and Privacy panel.

## Contributors

WNW gained funding for the study. MHI conducted the statistical analysis. WNW, BA, CG, RT and ED implemented the natural language processing algorithm and extracted phenotype data. WNW, HW, MM, ED, HZ, LS and AH provided methodological and statistical advice. MHI and WNW drafted and redrafted the manuscript. All authors reviewed the manuscript and provided feedback.

## Acknowledgements

We would like to thank NHS Scotland patients for providing the data involved in this study. We would like to acknowledge the eDRIS team (Public Health Scotland) - particularly Rebecca Fairnie – for their support in obtaining approvals, the provisioning and linking of data and facilitating access to the National Safe Haven. We would also like to thank Arlene Casey for assistance setting up the NLP algorithm within the National Safe Haven.

