## Supplementary Material for "Clinically reported covert cerebrovascular disease and risk of neurological disease: a whole-population cohort of 367,988 people using natural language processing"

*Index*

Table S1. Clinical codes and relevant clinical terminology used to define each outcome

Table S2. Clinical codes and relevant clinical terminology used to define each condition

Table S3. Clinical codes and relevant clinical terminology used to define morbidity history covariates

Table S4. Number of individuals excluded during sample selection for prior morbidities. Note that morbidities are not mutually exclusive

Table S5. Prevalence of each brain imaging phenotype by age band

Table S6. Risk table showing incidence of each outcome for the analytic cohort (N = 367,988)

Table S7. Risk table showing incidence of each outcome for the wider Scottish population (the analytic sample plus the unscanned population alive at 01/07/2010) (N = 4, 637,231)

Table S8. Median follow-up time for each outcome in the analytic cohort and wider Scottish population (the analytic sample plus the unscanned population alive at 01/07/2010)

Table S9. 1-year and 5-year unadjusted absolute risks (AR) for each scan phenotype and outcome, and the absolute risk increase (ARI) and relative risk ratios (RR) compared both to the analytic cohort without the phenotype and to the wider Scottish population

Table S10. 1- and 5-year adjusted absolute risks (AR), split by age group, for each outcome, and the absolute risk increase (ARI) and relative risk ratios (RR) compared to the wider Scottish population

Figure S1. Kaplan-Meier curves showing stroke survival for those with and without WMH, lacunes, cortical infarcts, cerebral atrophy, and any/no CCD phenotype

Figure S2. Kaplan-Meier curves showing dementia survival for those with and without WMH, lacunes, cortical infarcts, cerebral atrophy, and any/no CCD phenotype

Figure S3. Kaplan-Meier curves showing Parkinson’s disease survival for those with and without WMH, lacunes, cortical infarcts, cerebral atrophy, and any/no CCD phenotype

Figure S4. Kaplan-Meier curves showing epilepsy survival for those with and without WMH, lacunes, cortical infarcts, cerebral atrophy, and any/no CCD phenotype

Figure S5. Kaplan-Meier curves showing colorectal cancer survival for those with and without WMH, lacunes, cortical infarcts, cerebral atrophy, and any/no CCD phenotype

Table S11. Univariate hazard ratios (HR) and fully-adjusted hazard ratios (aHR), 95% CIs and p-values for each scan phenotype associated with each outcome in the primary analyses

Table S12. Univariate hazard ratios (HR) and fully-adjusted hazard ratios (aHR), 95% CIs and p-values for each scan phenotype associated with stroke and dementia subtypes in the primary analyses

Table S13. Adjusted hazard ratios (aHR), 95% CIs and p-values for each scan phenotype associated with each outcome after removing the first year of follow-up

Table S14. Adjusted hazard ratios (aHR), 95% CIs and p-values for each scan phenotype associated with each outcome after removing the first 5 years of follow-up

Table S15. Adjusted hazard Ratios (aHR), 95% CIs and p-values for each scan phenotype associated with each outcome, broken down by sex, age group and scan modality

Figure S6. Forest plot summarising event rates, hazard ratios and 95% CIs for each scan phenotype in the main analyses (Baseline), sex-specific models, age-specific models, and modality-specific models, split by outcome: stroke (A), dementia (B), Parkinson’s disease (C), epilepsy (D), and colorectal cancer (E)

Figure S7. Forest plot showing the event rate and adjusted hazard ratio (aHR) and 95% CIs per number of exposures (WMH, lacunes, cortical infarcts, and cerebral atrophy) and their association with risk of each outcome across follow-up.

Figure S8. Forest plot showing the event rate and adjusted hazard ratio (aHR) and 95% CIs per number of exposures (WMH, lacunes, cortical infarcts, and cerebral atrophy) and their association with risk of a) stroke subtypes and b) dementia subtypes across follow-up.

**Table S1. Clinical codes and relevant clinical terminology used to define each outcome**

| **Outcome** | **Subgroup** | **Code** | **Terminology** | **Description** |
| --- | --- | --- | --- | --- |
| Stroke | Intracerebral haemorrhage | I61 | ICD10 | Intracerebral haemorrhage |
| Stroke | Intracerebral haemorrhage | I610 | ICD10 | Intracerebral haemorrhage in hemisphere |
| Stroke | Intracerebral haemorrhage | I611 | ICD10 | Intracerebral haemorrhage in hemisphere |
| Stroke | Intracerebral haemorrhage | I612 | ICD10 | Intracerebral haemorrhage in hemisphere |
| Stroke | Intracerebral haemorrhage | I613 | ICD10 | Intracerebral haemorrhage in brain stem |
| Stroke | Intracerebral haemorrhage | I614 | ICD10 | Intracerebral haemorrhage in cerebellum |
| Stroke | Intracerebral haemorrhage | I615 | ICD10 | Intracerebral haemorrhage |
| Stroke | Intracerebral haemorrhage | I616 | ICD10 | Intracerebral haemorrhage |
| Stroke | Intracerebral haemorrhage | I618 | ICD10 | Other intracerebral haemorrhage |
| Stroke | Intracerebral haemorrhage | I619 | ICD10 | Intracerebral haemorrhage |
| Stroke | Ischaemic stroke | I63 | ICD10 | Cerebral infarction |
| Stroke | Ischaemic stroke | I630 | ICD10 | Cerebral infarction due to thrombosis of precerebral arteries |
| Stroke | Ischaemic stroke | I631 | ICD10 | Cerebral infarction due to embolism of precerebral arteries |
| Stroke | Ischaemic stroke | I632 | ICD10 | Cerebral infarction due to unspecified occlusion or stenosis of precerebral arteries |
| Stroke | Ischaemic stroke | I633 | ICD10 | Cerebral infarction due to thrombosis of cerebral arteries |
| Stroke | Ischaemic stroke | I634 | ICD10 | Cerebral infarction due to embolism of cerebral arteries |
| Stroke | Ischaemic stroke | I635 | ICD10 | Cerebral infarction due to unspecified occlusion or stenosis of cerebral arteries |
| Stroke | Ischaemic stroke | I636 | ICD10 | Cerebral infarction due to cerebral venous thrombosis |
| Stroke | Ischaemic stroke | I638 | ICD10 | Other cerebral infarction |
| Stroke | Ischaemic stroke | I639 | ICD10 | Cerebral infarction, unspecified |
| Stroke | Unspecified stroke | I64 | ICD10 | Stroke, not specified as haemorrhage or infarction |
| Stroke | Unspecified stroke | I64X | ICD10 | Stroke, not specified as haemorrhage or infarction |
| Dementia | Alzheimer's dementia | F00 | ICD10 | Dementia in Alzheimer disease |
| Dementia | Alzheimer's dementia | F000 | ICD10 | Dementia in Alzheimer disease with early onset |
| Dementia | Alzheimer's dementia | F001 | ICD10 | Dementia in Alzheimer disease with late onset |
| Dementia | Alzheimer's dementia | F002 | ICD10 | Dementia in Alzheimer disease, atypical or mixed type |
| Dementia | Alzheimer's dementia | F009 | ICD10 | Dementia in Alzheimer disease, unspecified |
| Dementia | Alzheimer's dementia | G30 | ICD10 | Alzheimer disease |
| Dementia | Alzheimer's dementia | G300 | ICD10 | Alzheimer disease with early onset |
| Dementia | Alzheimer's dementia | G301 | ICD10 | Alzheimer disease with late onset |
| Dementia | Alzheimer's dementia | G308 | ICD10 | Other Alzheimer disease |
| Dementia | Alzheimer's dementia | G309 | ICD10 | Alzheimer disease, unspecified |
| Dementia | Alzheimer's disease | 0411000D0 | BNF | Donepezil hydrochloride |
| Dementia | Alzheimer's disease | 0411000F0 | BNF | Galantamine |
| Dementia | Alzheimer's disease | 411000 | BNF | Rivastigmine (alzheimers and parkinson’s dementia and lewy body) |
| Dementia | Alzheimer's disease | 0411000G0 | BNF | Memantine hydrochloride |
| Dementia | Vascular dementia | F01 | ICD10 | Vascular dementia |
| Dementia | Vascular dementia | F010 | ICD10 | Vascular dementia of acute onset |
| Dementia | Vascular dementia | F011 | ICD10 | Multi-infarct dementia |
| Dementia | Vascular dementia | F012 | ICD10 | Subcortical vascular dementia |
| Dementia | Vascular dementia | F013 | ICD10 | Mixed cortical and subcortical vascular dementia |
| Dementia | Vascular dementia | F018 | ICD10 | Other vascular dementia |
| Dementia | Vascular dementia | F019 | ICD10 | Vascular dementia, unspecified |
| Dementia | Rare and Unspecified dementia | F02 | ICD10 | Dementia in other diseases classified elsewhere |
| Dementia | Rare and Unspecified dementia | F020 | ICD10 | Dementia in Pick disease |
| Dementia | Rare and Unspecified dementia | F021 | ICD10 | Dementia in Creutzfeldt-Jakob disease |
| Dementia | Rare and Unspecified dementia | F022 | ICD10 | Dementia in Huntington disease |
| Dementia | Rare and Unspecified dementia | F023 | ICD10 | Dementia in Parkinson disease |
| Dementia | Rare and Unspecified dementia | F024 | ICD10 | Dementia in human immunodeficiency virus [HIV] disease |
| Dementia | Rare and Unspecified dementia | F028 | ICD10 | Dementia in other specified diseases classified elsewhere |
| Dementia | Rare and Unspecified dementia | F03 | ICD10 | Unspecified dementia |
| Dementia | Rare and Unspecified dementia | F051 | ICD10 | Delirium superimposed on dementia |
| Parkinson's |  | F023 | ICD10 | Dementia in Parkinson's disease |
| Parkinson's |  | G20 | ICD10 | Parkinson's disease |
| Parkinson's |  | G21 | ICD10 | Secondary Parkinsonism |
| Parkinson's |  | G210 | ICD10 | Malignant neuroleptic syndrome |
| Parkinson's |  | G211 | ICD10 | Other drug-induced secondary parkinsonism |
| Parkinson's |  | G212 | ICD10 | Secondary parkinsonism due to other external agents |
| Parkinson's |  | G213 | ICD10 | Postencephalitic parkinsonism |
| Parkinson's |  | G214 | ICD10 | Vascular parkinsonism |
| Parkinson's |  | G218 | ICD10 | Other secondary parkinsonism |
| Parkinson's |  | G219 | ICD10 | Secondary parkinsonism, unspecified |
| Parkinson's |  | G22 | ICD10 | Parkinson's due to other diseases |
| Parkinson's |  | G231 | ICD10 | Progressive supranuclear palsy |
| Parkinson's |  | G232 | ICD10 | Multiple system atrophy, parkinsonian |
| Parkinson's |  | G233 | ICD10 | Multiple system atrophy, cerebellar |
| Parkinson's |  | G903 | ICD10 | Multiple system degeneration |
| Parkinson's |  | 0409010N0 | BNF | Co-careldopa |
| Parkinson's |  | 0409010K0 | BNF | Co-beneldopa (Madopar) |
| Parkinson's |  | 0409010Y0 | BNF | Rasagiline (monoamine-oxidase-B inhibitors) |
| Parkinson's |  | 0409010T0 | BNF | Selegiline hydrochloride (monoamine-oxidase-B inhibitors) |
| Parkinson's |  | 0409010W0 | BNF | Pramipexole (Non-ergot-derived dopamine-receptor agonists) |
| Parkinson's |  | 0409010H0 | BNF | Ropinirole (Non-ergot-derived dopamine-receptor agonists) |
| Parkinson's |  | 0409010Z0 | BNF | Rotigotine (Non-ergot-derived dopamine-receptor agonists) |
| Parkinson's |  | 0409010X0 | BNF | Levodopa/carbidopa/entacapone |
| Parkinson's |  | 0409010V0 | BNF | Entacapone (COMT inhib) |
| Parkinson's |  | 0409010S0 | BNF | Tolcapone (COMT inhib) |
| Epilepsy |  | G40 | ICD10 | Epilepsy |
| Epilepsy |  | G400 | ICD10 | Localization-related (focal)(partial) idiopathic epilepsy and epileptic syndromes with seizures of localized onset |
| Epilepsy |  | G401 | ICD10 | Localization-related (focal)(partial) symptomatic epilepsy and epileptic syndromes with simple partial seizures |
| Epilepsy |  | G402 | ICD10 | Localization-related (focal)(partial) symptomatic epilepsy and epileptic syndromes with complex partial seizures |
| Epilepsy |  | G403 | ICD10 | Generalized idiopathic epilepsy and epileptic syndromes |
| Epilepsy |  | G404 | ICD10 | Other generalized epilepsy and epileptic syndromes |
| Epilepsy |  | G405 | ICD10 | Special epileptic syndromes |
| Epilepsy |  | G406 | ICD10 | Grand mal seizures, unspecified (with or without petit mal) |
| Epilepsy |  | G407 | ICD10 | Petit mal, unspecified, without grand mal seizures |
| Epilepsy |  | G408 | ICD10 | Other epilepsy |
| Epilepsy |  | G409 | ICD10 | Epilepsy, unspecified |
| Epilepsy |  | G41 | ICD10 | Status epilepticus |
| Epilepsy |  | G410 | ICD10 | Grand mal status epilepticus |
| Epilepsy |  | G411 | ICD10 | Petit mal status epilepticus |
| Epilepsy |  | G412 | ICD10 | Complex partial status epilepticus |
| Epilepsy |  | G418 | ICD10 | Other status epilepticus |
| Epilepsy |  | G419 | ICD10 | Status epilepticus, unspecified |
| Epilepsy |  | R568 | ICD10 | Seizures |
| Epilepsy |  | 0408010C0 | BNF | Carbamazepine |
| Epilepsy |  | 040801060 | BNF | Clobazam |
| Epilepsy |  | 0408010AI | BNF | Eslicarbazepine |
| Epilepsy |  | 0408010I0 | BNF | Ethosuximide |
| Epilepsy |  | 0408010G0 | BNF | Gabapentin |
| Epilepsy |  | 0408010AH | BNF | Lacosamide |
| Epilepsy |  | 0408010H0 | BNF | Lamotrigine |
| Epilepsy |  | 0408010A0 | BNF | Levetiracetam |
| Epilepsy |  | 0408010D0 | BNF | Oxcarbazepine |
| Epilepsy |  | 0408010AK | BNF | Perampanel |
| Epilepsy |  | 0408010N0 | BNF | Phenobarbital |
| Epilepsy |  | 0408010Z0 | BNF | Phenytoin |
| Epilepsy |  | 0408020T0 | BNF | Phenytoin Sodium |
| Epilepsy |  | 0408010T0 | BNF | Phenytoin Sodium |
| Epilepsy |  | 0409030P0 | BNF | Piracetam |
| Epilepsy |  | 0408010U0 | BNF | Primidone |
| Epilepsy |  | 0408010AJ | BNF | Retigabine |
| Epilepsy |  | 0408010AF | BNF | Rufinamide |
| Epilepsy |  | 0408010W0 | BNF | Sodium valproate |
| Epilepsy |  | 0408010AB | BNF | Tiagabine |
| Epilepsy |  | 040801050 | BNF | Topiramate |
| Epilepsy |  | 0408010X0 | BNF | Vigabatrin |
| Epilepsy |  | 0408010AD | BNF | Zonisamide |
| Colorectal cancer |  | C18 | ICD10 | Malignant neoplasm of colon |
| Colorectal cancer |  | C180 | ICD10 | Malignant neoplasm of colon |
| Colorectal cancer |  | C1809 | ICD10 | Malignant neoplasm of colon |
| Colorectal cancer |  | C181 | ICD10 | Malignant neoplasm of colon |
| Colorectal cancer |  | C182 | ICD10 | Malignant neoplasm of colon |
| Colorectal cancer |  | C183 | ICD10 | Malignant neoplasm of colon |
| Colorectal cancer |  | C184 | ICD10 | Malignant neoplasm of colon |
| Colorectal cancer |  | C185 | ICD10 | Malignant neoplasm of colon |
| Colorectal cancer |  | C186 | ICD10 | Malignant neoplasm of colon |
| Colorectal cancer |  | C187 | ICD10 | Malignant neoplasm of colon |
| Colorectal cancer |  | C188 | ICD10 | Malignant neoplasm of colon |
| Colorectal cancer |  | C189 | ICD10 | Malignant neoplasm of colon |
| Colorectal cancer |  | C19 | ICD10 | Malignant neoplasm of rectosigmoid junction |
| Colorectal cancer |  | C19X | ICD10 | Malignant neoplasm of rectosigmoid junction |
| Colorectal cancer |  | C20 | ICD10 | Malignant neoplasm of rectum |
| Colorectal cancer |  | C20X | ICD10 | Malignant neoplasm of rectum |
| Colorectal cancer |  | C20X1 | ICD10 | Malignant neoplasm of rectum |
| Colorectal cancer |  | C21 | ICD10 | Malignant neoplasm of anus and anal canal |
| Colorectal cancer |  | C210 | ICD10 | Malignant neoplasm of anus and anal canal |
| Colorectal cancer |  | C211 | ICD10 | Malignant neoplasm of anus and anal canal |
| Colorectal cancer |  | C212 | ICD10 | Malignant neoplasm of anus and anal canal |
| Colorectal cancer |  | C218 | ICD10 | Malignant neoplasm of anus and anal canal |

**Table S2. Clinical codes and relevant clinical terminology used to define each condition**

| **Condition** | **Code** | **Terminology** | **Description** |
| --- | --- | --- | --- |
| Transient Ischaemic Attack | G450 | ICD10 | Vertebro-basilar artery syndrome |
| Transient Ischaemic Attack | G451 | ICD10 | Carotid artery syndrome (hemispheric) |
| Transient Ischaemic Attack | G452 | ICD10 | Multiple and bilateral precerebral artery syndromes |
| Transient Ischaemic Attack | G453 | ICD10 | Amaurosis fugax |
| Transient Ischaemic Attack | G458 | ICD10 | Other transient cerebral ischaemic attacks and related syndromes |
| Transient Ischaemic Attack | G459 | ICD10 | Transient cerebral ischaemic attack, unspecified |
| Transient Ischaemic Attack | G460 | ICD10 | Middle cerebral artery syndrome |
| Transient Ischaemic Attack | G461 | ICD10 | Anterior cerebral artery syndrome |
| Transient Ischaemic Attack | G462 | ICD10 | Posterior cerebral artery syndrome |
| Transient Ischaemic Attack | I65 | ICD10 | Occlusion and stenosis of precerebral arteries, not resulting in cerebral infarction |
| Transient Ischaemic Attack | I650 | ICD10 | Occlusion and stenosis of vertebral artery, not resulting in cerebral infarction |
| Transient Ischaemic Attack | I651 | ICD10 | Occlusion and stenosis of basilar artery, not resulting in cerebral infarction |
| Transient Ischaemic Attack | I652 | ICD10 | Occlusion and stenosis of carotid artery, not resulting in cerebral infarction |
| Transient Ischaemic Attack | I653 | ICD10 | Occlusion and stenosis of multiple and bilateral precerebral arteries, not resulting in cerebral infarction |
| Transient Ischaemic Attack | I658 | ICD10 | Occlusion and stenosis of other precerebral artery, not resulting in cerebral infarction |
| Transient Ischaemic Attack | I659 | ICD10 | Occlusion and stenosis of unspecified precerebral artery, not resulting in cerebral infarction |
| Transient Ischaemic Attack | I66 | ICD10 | Occlusion and stenosis of cerebral arteries, not resulting in cerebral infarction |
| Transient Ischaemic Attack | I660 | ICD10 | Occlusion and stenosis of middle cerebral artery, not resulting in cerebral infarction |
| Transient Ischaemic Attack | I661 | ICD10 | Occlusion and stenosis of anterior cerebral artery, not resulting in cerebral infarction |
| Transient Ischaemic Attack | I662 | ICD10 | Occlusion and stenosis of posterior cerebral artery, not resulting in cerebral infarction |
| Transient Ischaemic Attack | I663 | ICD10 | Occlusion and stenosis of cerebellar arteries, not resulting in cerebral infarction |
| Transient Ischaemic Attack | I664 | ICD10 | Occlusion and stenosis of multiple and bilateral cerebral arteries, not resulting in cerebral infarction |
| Transient Ischaemic Attack | I668 | ICD10 | Occlusion and stenosis of other cerebral artery, not resulting in cerebral infarction |
| Transient Ischaemic Attack | I669 | ICD10 | Occlusion and stenosis of unspecified cerebral artery, not resulting in cerebral infarction |
| Transient Ischaemic Attack | H34 | ICD10 | Retinal vascular occlusions |
| Transient Ischaemic Attack | H340 | ICD10 | Transient retinal artery occlusion |
| Transient Ischaemic Attack | H341 | ICD10 | Central retinal artery occlusion |
| Transient Ischaemic Attack | H342 | ICD10 | Other retinal artery occlusions |
| Transient Ischaemic Attack | H348 | ICD10 | Other retinal vascular occlusions |
| Transient Ischaemic Attack | H349 | ICD10 | Retinal vascular occlusion, unspecified |
| External Haematoma | I621 | ICD10 | Non-traumatic extradural haemorrhage (Non-traumatic epidural haemorrhage) |
| Intracranial Haemorrhage | I629 | ICD10 | Intracranial haemorrhage (nontraumatic), unspecified |
| Intracranial Haemorrhage | I692 | ICD10 | Sequelae of other nontraumatic intracranial haemorrhage |
| Subarachnoid haemorrhage | I60 | ICD10 | Subarachnoid haemorrhage |
| Subarachnoid haemorrhage | I600 | ICD10 | Subarachnoid haemorrhage from carotid siphon and bifurcation |
| Subarachnoid haemorrhage | I601 | ICD10 | Subarachnoid haemorrhage from middle cerebral artery |
| Subarachnoid haemorrhage | I602 | ICD10 | Subarachnoid haemorrhage from anterior communicating artery |
| Subarachnoid haemorrhage | I603 | ICD10 | Subarachnoid haemorrhage from posterior communicating artery |
| Subarachnoid haemorrhage | I604 | ICD10 | Subarachnoid haemorrhage from basilar artery |
| Subarachnoid haemorrhage | I605 | ICD10 | Subarachnoid haemorrhage from vertebral artery |
| Subarachnoid haemorrhage | I606 | ICD10 | Subarachnoid haemorrhage from other intracranial arteries |
| Subarachnoid haemorrhage | I607 | ICD10 | Subarachnoid haemorrhage from intracranial artery |
| Subarachnoid haemorrhage | I608 | ICD10 | Other subarachnoid haemorrhage |
| Subarachnoid haemorrhage | I609 | ICD10 | Subarachnoid haemorrhage |
| Subarachnoid haemorrhage | I690 | ICD10 | Sequelae of subarachnoid haemorrhage |
| Subdural Haemorrhage | I620 | ICD10 | Subdural haemorrhage (acute)(nontraumatic) |

**Table S3. Clinical codes and relevant clinical terminology used to define morbidity history covariates**

| **Condition** | **Code** | **Terminology** | **Description** |
| --- | --- | --- | --- |
| Large Vessel Disease | I21 | ICD10 | Acute myocardial infarction |
| Large Vessel Disease | I210 | ICD10 | Acute transmural myocardial infarction of anterior wall |
| Large Vessel Disease | I211 | ICD10 | Acute transmural myocardial infarction of inferior wall |
| Large Vessel Disease | I212 | ICD10 | Acute transmural myocardial infarction of other sites |
| Large Vessel Disease | I213 | ICD10 | Acute transmural myocardial infarction of unspecified site |
| Large Vessel Disease | I214 | ICD10 | Acute subendocardial myocardial infarction |
| Large Vessel Disease | I219 | ICD10 | Acute myocardial infarction, unspecified |
| Large Vessel Disease | I22 | ICD10 | Subsequent myocardial infarction |
| Large Vessel Disease | I220 | ICD10 | Subsequent myocardial infarction of anterior wall |
| Large Vessel Disease | I221 | ICD10 | Subsequent myocardial infarction of inferior wall |
| Large Vessel Disease | I228 | ICD10 | Subsequent myocardial infarction of other sites |
| Large Vessel Disease | I229 | ICD10 | Subsequent myocardial infarction of unspecified site |
| Large Vessel Disease | I23 | ICD10 | Certain current complications following acute myocardial infarction |
| Large Vessel Disease | I230 | ICD10 | Haemopericardium as current complication following acute myocardial infarction |
| Large Vessel Disease | I231 | ICD10 | Atrial septal defect as current complication following acute myocardial infarction |
| Large Vessel Disease | I232 | ICD10 | Ventricular septal defect as current complication following acute myocardial infarction |
| Large Vessel Disease | I233 | ICD10 | Rupture of cardiac wall without haemopericardium as current complication following acute myocardial |
| Large Vessel Disease | I234 | ICD10 | Rupture of chordae tendineae as current complication following acute myocardial infarction |
| Large Vessel Disease | I235 | ICD10 | Rupture of papillary muscle as current complication following acute myocardial infarction |
| Large Vessel Disease | I236 | ICD10 | Thrombosis of atrium, auricular appendage, and ventricle as current complications following acute my |
| Large Vessel Disease | I238 | ICD10 | Other current complications following acute myocardial infarction |
| Large Vessel Disease | I241 | ICD10 | Dressler's syndrome |
| Large Vessel Disease | I252 | ICD10 | Old myocardial infarction |
| Large Vessel Disease | I20 | ICD10 | Angina pectoris |
| Large Vessel Disease | I200 | ICD10 | Unstable angina |
| Large Vessel Disease | I201 | ICD10 | Angina pectoris with documented spasm |
| Large Vessel Disease | I208 | ICD10 | Other forms of angina pectoris |
| Large Vessel Disease | I209 | ICD10 | Angina pectoris, unspecified |
| Large Vessel Disease | I731 | ICD10 | Thromboangiitis obliterans [Buerger] |
| Large Vessel Disease | I738 | ICD10 | Other specified peripheral vascular diseases |
| Large Vessel Disease | E105 | ICD10 | Insulin-dependent diabetes mellitus with peripheral circulatory complications |
| Large Vessel Disease | T87 | ICD10 | Complications peculiar to reattachment and amputation |
| Large Vessel Disease | I702 | ICD10 | Atherosclerosis of arteries of extremities |
| Large Vessel Disease | E115 | ICD10 | Non-insulin-dependent diabetes mellitus with peripheral circulatory complications |
| Large Vessel Disease | E145 | ICD10 | Unspecified diabetes mellitus with peripheral circulatory complications |
| Large Vessel Disease | I739 | ICD10 | Peripheral vascular disease, unspecified |
| Large Vessel Disease | I74 | ICD10 | Arterial embolism and thrombosis |
| Large Vessel Disease | I740 | ICD10 | Embolism and thrombosis of abdominal aorta |
| Large Vessel Disease | I741 | ICD10 | Embolism and thrombosis of other and unspecified parts of aorta |
| Large Vessel Disease | I742 | ICD10 | Embolism and thrombosis of arteries of upper extremities |
| Large Vessel Disease | I743 | ICD10 | Embolism and thrombosis of arteries of lower extremities |
| Large Vessel Disease | I744 | ICD10 | Embolism and thrombosis of arteries of extremities, unspecified |
| Large Vessel Disease | I745 | ICD10 | Embolism and thrombosis of iliac artery |
| Large Vessel Disease | I748 | ICD10 | Embolism and thrombosis of other arteries |
| Large Vessel Disease | I749 | ICD10 | Embolism and thrombosis of unspecified artery |
| Small Vessel Disease | N00 | ICD10 | Acute nephritic syndrome |
| Small Vessel Disease | N000 | ICD10 | Acute nephritic syndrome: Minor glomerular abnormality |
| Small Vessel Disease | N001 | ICD10 | Acute nephritic syndrome: Focal and segmental glomerular lesions |
| Small Vessel Disease | N002 | ICD10 | Acute nephritic syndrome: Diffuse membranous glomerulonephritis |
| Small Vessel Disease | N003 | ICD10 | Acute nephritic syndrome: Diffuse mesangial proliferative glomerulonephritis |
| Small Vessel Disease | N004 | ICD10 | Acute nephritic syndrome: Diffuse endocapillary proliferative glomerulonephritis |
| Small Vessel Disease | N005 | ICD10 | Acute nephritic syndrome: Diffuse mesangiocapillary glomerulonephritis |
| Small Vessel Disease | N006 | ICD10 | Acute nephritic syndrome: Dense deposit disease |
| Small Vessel Disease | N007 | ICD10 | Acute nephritic syndrome: Diffuse crescentic glomerulonephritis |
| Small Vessel Disease | N008 | ICD10 | Acute nephritic syndrome: Other |
| Small Vessel Disease | N009 | ICD10 | Acute nephritic syndrome: Unspecified |
| Small Vessel Disease | N01 | ICD10 | Rapidly progressive nephritic syndrome |
| Small Vessel Disease | N010 | ICD10 | Rapidly progressive nephritic syndrome: Minor glomerular abnormality |
| Small Vessel Disease | N011 | ICD10 | Rapidly progressive nephritic syndrome: Focal and segmental glomerular lesions |
| Small Vessel Disease | N012 | ICD10 | Rapidly progressive nephritic syndrome: Diffuse membranous glomerulonephritis |
| Small Vessel Disease | N013 | ICD10 | Rapidly progressive nephritic syndrome: Diffuse mesangial proliferative glomerulonephritis |
| Small Vessel Disease | N014 | ICD10 | Rapidly progressive nephritic syndrome: Diffuse endocapillary proliferative glomerulonephritis |
| Small Vessel Disease | N015 | ICD10 | Rapidly progressive nephritic syndrome: Diffuse mesangiocapillary glomerulonephritis |
| Small Vessel Disease | N016 | ICD10 | Rapidly progressive nephritic syndrome: Dense deposit disease |
| Small Vessel Disease | N017 | ICD10 | Rapidly progressive nephritic syndrome: Diffuse crescentic glomerulonephritis |
| Small Vessel Disease | N018 | ICD10 | Rapidly progressive nephritic syndrome: Other |
| Small Vessel Disease | N019 | ICD10 | Rapidly progressive nephritic syndrome: Unspecified |
| Small Vessel Disease | N03 | ICD10 | Chronic nephritic syndrome |
| Small Vessel Disease | N030 | ICD10 | Chronic nephritic syndrome: Minor glomerular abnormality |
| Small Vessel Disease | N031 | ICD10 | Chronic nephritic syndrome: Focal and segmental glomerular lesions |
| Small Vessel Disease | N032 | ICD10 | Chronic nephritic syndrome: Diffuse membranous glomerulonephritis |
| Small Vessel Disease | N033 | ICD10 | Chronic nephritic syndrome: Diffuse mesangial proliferative glomerulonephritis |
| Small Vessel Disease | N034 | ICD10 | Chronic nephritic syndrome: Diffuse endocapillary proliferative glomerulonephritis |
| Small Vessel Disease | N035 | ICD10 | Chronic nephritic syndrome: Diffuse mesangiocapillary glomerulonephritis |
| Small Vessel Disease | N036 | ICD10 | Chronic nephritic syndrome: Dense deposit disease |
| Small Vessel Disease | N037 | ICD10 | Chronic nephritic syndrome: Diffuse crescentic glomerulonephritis |
| Small Vessel Disease | N038 | ICD10 | Chronic nephritic syndrome: Other |
| Small Vessel Disease | N039 | ICD10 | Chronic nephritic syndrome: Unspecified |
| Small Vessel Disease | N052 | ICD10 | Unspecified nephritic syndrome |
| Small Vessel Disease | N053 | ICD10 | Unspecified nephritic syndrome |
| Small Vessel Disease | N054 | ICD10 | Unspecified nephritic syndrome |
| Small Vessel Disease | N055 | ICD10 | Unspecified nephritic syndrome |
| Small Vessel Disease | N056 | ICD10 | Unspecified nephritic syndrome |
| Small Vessel Disease | N072 | ICD10 | Hereditary nephropathy, not elsewhere classified ; Diffuse membranous glomerulonephritis |
| Small Vessel Disease | N073 | ICD10 | Hereditary nephropathy, not elsewhere classified ; Diffuse mesangial proliferative glomerulonephritis |
| Small Vessel Disease | N074 | ICD10 | Hereditary nephropathy, not elsewhere classified ; Diffuse endocapillary proliferative glomerulonephritis |
| Small Vessel Disease | N10 | ICD10 | Acute tubulo-interstitial nephritis |
| Small Vessel Disease | N17 | ICD10 | Acute renal failure |
| Small Vessel Disease | N170 | ICD10 | Acute renal failure with tubular necrosis |
| Small Vessel Disease | N171 | ICD10 | Acute renal failure with acute cortical necrosis |
| Small Vessel Disease | N172 | ICD10 | Acute renal failure with medullary necrosis |
| Small Vessel Disease | N178 | ICD10 | Other acute renal failure |
| Small Vessel Disease | N179 | ICD10 | Acute renal failure, unspecified |
| Small Vessel Disease | N181 | ICD10 | Chronic kidney  disease, stage 1 |
| Small Vessel Disease | N182 | ICD10 | Chronic kidney  disease, stage 2 |
| Small Vessel Disease | N183 | ICD10 | Chronic kidney disease, stage 3 |
| Small Vessel Disease | N184 | ICD10 | Chronic kidney disease, stage 4 |
| Small Vessel Disease | N185 | ICD10 | Chronic kidney disease, stage 5 |
| Small Vessel Disease | N189 | ICD10 | Chronic kidney disease, unspecified |
| Small Vessel Disease | N19 | ICD10 | Unspecified kidney failure |
| Small Vessel Disease | N25 | ICD10 | Disorders resulting from impaired renal tubular function |
| Small Vessel Disease | N250 | ICD10 | Renal osteodystrophy |
| Small Vessel Disease | N251 | ICD10 | Nephrogenic diabetes insipidus |
| Small Vessel Disease | N258 | ICD10 | Other disorders resulting from impaired renal tubular function |
| Small Vessel Disease | N259 | ICD10 | Disorder resulting from impaired renal tubular function, unspecified |
| Small Vessel Disease | T861 | ICD10 | Kidney transplant failure and rejection |
| Small Vessel Disease | Y841 | ICD10 | Kidney dialysis |
| Small Vessel Disease | Z490 | ICD10 | Preparatory care for dialysis |
| Small Vessel Disease | Z491 | ICD10 | Extracorporeal dialysis |
| Small Vessel Disease | Z492 | ICD10 | Other dialysis |
| Small Vessel Disease | Z940 | ICD10 | Kidney transplant status |
| Small Vessel Disease | Z992 | ICD10 | Dependence on renal dialysis |
| Small Vessel Disease | N165 | ICD10 | Renal tubulo-interstitial disorders in transplant rejection |
| Small Vessel Disease | T824 | ICD10 | Mechanical complication of vascular dialysis catheter |
| Small Vessel Disease | Y602 | ICD10 | Unintentional cut, puncture, perforation or haemorrhage during surgical and medical care: During kidney dialysis or other perfusion |
| Small Vessel Disease | Y612 | ICD10 | Foreign object accidentally left in body during surgical and medical care: During kidney dialysis or other perfusion |
| Small Vessel Disease | Y622 | ICD10 | Failure of sterile precautions during surgical and medical care: During kidney dialysis or other perfusion |
| Small Vessel Disease | I770 | ICD10 | arteriovenous fistula, acquired |
| Heart Failure | I110 | ICD10 | Hypertensive heart disease with (congestive) heart failure |
| Heart Failure | I130 | ICD10 | Hypertensive heart and renal disease with (congestive) heart failure |
| Heart Failure | I132 | ICD10 | Hypertensive heart and renal disease with both (congestive) heart failure and renal failure |
| Heart Failure | I50 | ICD10 | Heart failure |
| Heart Failure | I500 | ICD10 | Congestive heart failure |
| Heart Failure | I501 | ICD10 | Left ventricular failure |
| Heart Failure | I509 | ICD10 | Heart failure, unspecified |
| Any cancer | C00 | ICD10 | Malignant neoplasm of lip |
| Any cancer | C01 | ICD10 | Malignant neoplasm of base of tongue |
| Any cancer | C02 | ICD10 | Malignant neoplasm of other and unspecified parts of tongue |
| Any cancer | C03 | ICD10 | Malignant neoplasm of gum |
| Any cancer | C04 | ICD10 | Malignant neoplasm of floor of mouth |
| Any cancer | C05 | ICD10 | Malignant neoplasm of palate |
| Any cancer | C06 | ICD10 | Malignant neoplasm of other and unspecified parts of mouth |
| Any cancer | C07 | ICD10 | Malignant neoplasm of parotid gland |
| Any cancer | C08 | ICD10 | Malignant neoplasm of other and unspecified major salivary glands |
| Any cancer | C09 | ICD10 | Malignant neoplasm of tonsil |
| Any cancer | C10 | ICD10 | Malignant neoplasm of oropharynx |
| Any cancer | C11 | ICD10 | Malignant neoplasm of nasopharynx |
| Any cancer | C12 | ICD10 | Malignant neoplasm of pyriform sinus |
| Any cancer | C13 | ICD10 | Malignant neoplasm of hypopharynx |
| Any cancer | C14 | ICD10 | Malignant neoplasm of other and ill-defined sites in the lip, oral cavity and pharynx |
| Any cancer | C15 | ICD10 | Malignant neoplasm of oesophagus |
| Any cancer | C16 | ICD10 | Malignant neoplasm of stomach |
| Any cancer | C17 | ICD10 | Malignant neoplasm of small intestine |
| Any cancer | C18 | ICD10 | Malignant neoplasm of colon |
| Any cancer | C180 | ICD10 | Malignant neoplasm of colon |
| Any cancer | C1809 | ICD10 | Malignant neoplasm of colon |
| Any cancer | C181 | ICD10 | Malignant neoplasm of colon |
| Any cancer | C182 | ICD10 | Malignant neoplasm of colon |
| Any cancer | C183 | ICD10 | Malignant neoplasm of colon |
| Any cancer | C184 | ICD10 | Malignant neoplasm of colon |
| Any cancer | C185 | ICD10 | Malignant neoplasm of colon |
| Any cancer | C186 | ICD10 | Malignant neoplasm of colon |
| Any cancer | C187 | ICD10 | Malignant neoplasm of colon |
| Any cancer | C188 | ICD10 | Malignant neoplasm of colon |
| Any cancer | C189 | ICD10 | Malignant neoplasm of colon |
| Any cancer | C19 | ICD10 | Malignant neoplasm of rectosigmoid junction |
| Any cancer | C19X | ICD10 | Malignant neoplasm of rectosigmoid junction |
| Any cancer | C20 | ICD10 | Malignant neoplasm of rectum |
| Any cancer | C20X | ICD10 | Malignant neoplasm of rectum |
| Any cancer | C20X1 | ICD10 | Malignant neoplasm of rectum |
| Any cancer | C21 | ICD10 | Malignant neoplasm of anus and anal canal |
| Any cancer | C210 | ICD10 | Malignant neoplasm of anus and anal canal |
| Any cancer | C211 | ICD10 | Malignant neoplasm of anus and anal canal |
| Any cancer | C212 | ICD10 | Malignant neoplasm of anus and anal canal |
| Any cancer | C218 | ICD10 | Malignant neoplasm of anus and anal canal |
| Any cancer | C22 | ICD10 | Malignant neoplasm of liver and intrahepatic bile ducts |
| Any cancer | C23 | ICD10 | Malignant neoplasm of gallbladder |
| Any cancer | C24 | ICD10 | Malignant neoplasm of other and unspecified parts of biliary tract |
| Any cancer | C25 | ICD10 | Malignant neoplasm of pancreas |
| Any cancer | C26 | ICD10 | Malignant neoplasm of other and ill-defined digestive organs |
| Any cancer | C30 | ICD10 | Malignant neoplasm of nasal cavity and middle ear |
| Any cancer | C31 | ICD10 | Malignant neoplasm of accessory sinuses |
| Any cancer | C32 | ICD10 | Malignant neoplasm of larynx |
| Any cancer | C33 | ICD10 | Malignant neoplasm of trachea |
| Any cancer | C34 | ICD10 | Malignant neoplasm of bronchus and lung |
| Any cancer | C37 | ICD10 | Malignant neoplasm of thymus |
| Any cancer | C38 | ICD10 | Malignant neoplasm of heart, mediastinum and pleura |
| Any cancer | C39 | ICD10 | Malignant neoplasm of other and ill-defined sites in the respiratory system and intrathoracic organs |
| Any cancer | C40 | ICD10 | Malignant neoplasm of bone and articular cartilage of limbs |
| Any cancer | C41 | ICD10 | Malignant neoplasm of bone and articular cartilage of other and unspecified sites |
| Any cancer | C42 | ICD10 | hematopoietic and reticuloendothelial systems (ICD-O-3 specific) |
| Any cancer | C43 | ICD10 | Malignant melanoma of skin |
| Any cancer | C44 | ICD10 | Other malignant neoplasms of skin |
| Any cancer | C45 | ICD10 | Mesothelioma |
| Any cancer | C46 | ICD10 | Kaposi's sarcoma |
| Any cancer | C47 | ICD10 | Malignant neoplasm of peripheral nerves and autonomic nervous system |
| Any cancer | C48 | ICD10 | Malignant neoplasm of retroperitoneum and peritoneum |
| Any cancer | C49 | ICD10 | Malignant neoplasm of other connective and soft tissue |
| Any cancer | C50 | ICD10 | Malignant neoplasm of breast |
| Any cancer | C51 | ICD10 | Malignant neoplasm of vulva |
| Any cancer | C52 | ICD10 | Malignant neoplasm of vagina |
| Any cancer | C53 | ICD10 | Malignant neoplasm of cervix uteri |
| Any cancer | C54 | ICD10 | Malignant neoplasm of corpus uteri |
| Any cancer | C55 | ICD10 | Malignant neoplasm of uterus, part unspecified |
| Any cancer | C56 | ICD10 | Malignant neoplasm of ovary |
| Any cancer | C57 | ICD10 | Malignant neoplasm of other and unspecified female genital organs |
| Any cancer | C58 | ICD10 | Malignant neoplasm of placenta |
| Any cancer | C60 | ICD10 | Malignant neoplasm of penis |
| Any cancer | C61 | ICD10 | Malignant neoplasm of prostate |
| Any cancer | C62 | ICD10 | Malignant neoplasm of testis |
| Any cancer | C63 | ICD10 | Malignant neoplasm of other and unspecified male genital organs |
| Any cancer | C64 | ICD10 | Malignant neoplasm of kidney, except renal pelvis |
| Any cancer | C65 | ICD10 | Malignant neoplasm of renal pelvis |
| Any cancer | C66 | ICD10 | Malignant neoplasm of ureter |
| Any cancer | C67 | ICD10 | Malignant neoplasm of bladder |
| Any cancer | C68 | ICD10 | Malignant neoplasm of other and unspecified urinary organs |
| Any cancer | C69 | ICD10 | Malignant neoplasm of eye and adnexa |
| Any cancer | C70 | ICD10 | Malignant neoplasm of meninges |
| Any cancer | C71 | ICD10 | Malignant neoplasm of brain |
| Any cancer | C72 | ICD10 | Malignant neoplasm of spinal cord, cranial nerves and other parts of central nervous system |
| Any cancer | C73 | ICD10 | Malignant neoplasm of thyroid gland |
| Any cancer | C74 | ICD10 | Malignant neoplasm of adrenal gland |
| Any cancer | C75 | ICD10 | Malignant neoplasm of other endocrine glands and related structures |
| Any cancer | C79 | ICD10 | Secondary malignant neoplasm of other and unspecified sites |
| Diabetes | E10 | ICD10 | Insulin-dependent diabetes mellitus |
| Diabetes | E100 | ICD10 | Insulin-dependent diabetes mellitus: with coma |
| Diabetes | E101 | ICD10 | Insulin-dependent diabetes mellitus: with ketoacidosis |
| Diabetes | E102 | ICD10 | Insulin-dependent diabetes mellitus: with renal complications |
| Diabetes | E103 | ICD10 | Insulin-dependent diabetes mellitus: with ophthalmic complications |
| Diabetes | E104 | ICD10 | Insulin-dependent diabetes mellitus: with neurological complications |
| Diabetes | E105 | ICD10 | Insulin-dependent diabetes mellitus: with peripheral circulatory complications |
| Diabetes | E106 | ICD10 | Insulin-dependent diabetes mellitus: with other specified complications |
| Diabetes | E107 | ICD10 | Insulin-dependent diabetes mellitus: with multiple complications |
| Diabetes | E108 | ICD10 | Insulin-dependent diabetes mellitus: with unspecified complications |
| Diabetes | E109 | ICD10 | Insulin-dependent diabetes mellitus: without complications |
| Diabetes | E11 | ICD10 | Non-insulin-dependent diabetes mellitus |
| Diabetes | E110 | ICD10 | Non-insulin-dependent diabetes mellitus: with coma |
| Diabetes | E111 | ICD10 | Non-insulin-dependent diabetes mellitus: with ketoacidosis |
| Diabetes | E112 | ICD10 | Non-insulin-dependent diabetes mellitus: with renal complications |
| Diabetes | E113 | ICD10 | Non-insulin-dependent diabetes mellitus: with ophthalmic complications |
| Diabetes | E114 | ICD10 | Non-insulin-dependent diabetes mellitus: with neurological complications |
| Diabetes | E115 | ICD10 | Non-insulin-dependent diabetes mellitus: with peripheral circulatory complications |
| Diabetes | E116 | ICD10 | Non-insulin-dependent diabetes mellitus: with other specified complications |
| Diabetes | E117 | ICD10 | Non-insulin-dependent diabetes mellitus: with multiple complications |
| Diabetes | E118 | ICD10 | Non-insulin-dependent diabetes mellitus: with unspecified complications |
| Diabetes | E119 | ICD10 | Non-insulin-dependent diabetes mellitus: without complications |
| Diabetes | E12 | ICD10 | Malnutrition-related diabetes mellitus |
| Diabetes | E120 | ICD10 | Malnutrition-related diabetes mellitus: with coma |
| Diabetes | E121 | ICD10 | Malnutrition-related diabetes mellitus: with ketoacidosis |
| Diabetes | E122 | ICD10 | Malnutrition-related diabetes mellitus: with renal complications |
| Diabetes | E123 | ICD10 | Malnutrition-related diabetes mellitus: with ophthalmic complications |
| Diabetes | E124 | ICD10 | Malnutrition-related diabetes mellitus: with neurological complications |
| Diabetes | E125 | ICD10 | Malnutrition-related diabetes mellitus: with peripheral circulatory complications |
| Diabetes | E126 | ICD10 | Malnutrition-related diabetes mellitus: with other specified complications |
| Diabetes | E127 | ICD10 | Malnutrition-related diabetes mellitus: with multiple complications |
| Diabetes | E128 | ICD10 | Malnutrition-related diabetes mellitus: with unspecified complications |
| Diabetes | E129 | ICD10 | Malnutrition-related diabetes mellitus: without complications |
| Diabetes | E13 | ICD10 | Other specified diabetes mellitus |
| Diabetes | E130 | ICD10 | Other specified diabetes mellitus: with coma |
| Diabetes | E131 | ICD10 | Other specified diabetes mellitus: with ketoacidosis |
| Diabetes | E132 | ICD10 | Other specified diabetes mellitus: with renal complications |
| Diabetes | E133 | ICD10 | Other specified diabetes mellitus: with ophthalmic complications |
| Diabetes | E134 | ICD10 | Other specified diabetes mellitus: with neurological complications |
| Diabetes | E135 | ICD10 | Other specified diabetes mellitus: with peripheral circulatory complications |
| Diabetes | E136 | ICD10 | Other specified diabetes mellitus: with other specified complications |
| Diabetes | E137 | ICD10 | Other specified diabetes mellitus: with multiple complications |
| Diabetes | E138 | ICD10 | Other specified diabetes mellitus: with unspecified complications |
| Diabetes | E139 | ICD10 | Other specified diabetes mellitus: without complications |
| Diabetes | E14 | ICD10 | Unspecified diabetes mellitus |
| Diabetes | E140 | ICD10 | Unspecified diabetes mellitus: with coma |
| Diabetes | E141 | ICD10 | Unspecified diabetes mellitus: with ketoacidosis |
| Diabetes | E142 | ICD10 | Unspecified diabetes mellitus: with renal complications |
| Diabetes | E143 | ICD10 | Unspecified diabetes mellitus: with ophthalmic complications |
| Diabetes | E144 | ICD10 | Unspecified diabetes mellitus: with neurological complications |
| Diabetes | E145 | ICD10 | Unspecified diabetes mellitus: with peripheral circulatory complications |
| Diabetes | E146 | ICD10 | Unspecified diabetes mellitus: with other specified complications |
| Diabetes | E147 | ICD10 | Unspecified diabetes mellitus: with multiple complications |
| Diabetes | E148 | ICD10 | Unspecified diabetes mellitus: with unspecified complications |
| Diabetes | E149 | ICD10 | Unspecified diabetes mellitus: without complications |
| Diabetes | G590 | ICD10 | Diabetic mononeuropathy |
| Diabetes | G632 | ICD10 | Diabetic polyneuropathy |
| Diabetes | H280 | ICD10 | Diabetic cataract |
| Diabetes | H360 | ICD10 | Diabetic retinopathy |
| Diabetes | M142 | ICD10 | Diabetic arthropathy |
| Diabetes | N083 | ICD10 | Glomerular disorders in diabetes mellitus |
| Diabetes | O240 | ICD10 | Diabetes mellitus in pregnancy: Pre-existing diabetes mellitus, insulin-dependent |
| Diabetes | O241 | ICD10 | Diabetes mellitus in pregnancy: Pre-existing diabetes mellitus, non-insulin-dependent |
| Diabetes | O242 | ICD10 | Diabetes mellitus in pregnancy: Pre-existing malnutrition-related diabetes mellitus |
| Diabetes | O243 | ICD10 | Diabetes mellitus in pregnancy: Pre-existing diabetes mellitus, unspecified |
| Antihypertensives | 0202010B0AAABAB | BNF | Bendroflumethiazide 25mg tablets |
| Antihypertensives | 0202010B0AAACAC | BNF | Bendroflumethiazide 5mg tablets |
| Antihypertensives | 0202010B0AAAQAQ | BNF | Bendroflumethiazide 5mg/5ml oral suspension |
| Antihypertensives | 0202010B0AAAUAU | BNF | Bendroflumethiazide 125mg/5ml oral suspension |
| Antihypertensives | 0202010B0AAAXAX | BNF | Bendroflumethiazide 25mg/5ml oral suspension |
| Antihypertensives | 0202010B0BBAAAB | BNF | Aprinox 25mg tablets |
| Antihypertensives | 0202010B0BEABAB | BNF | Neo-Naclex 25mg tablets |
| Antihypertensives | 0202010D0AAAFAF | BNF | Chlorothiazide 500mg tablets |
| Antihypertensives | 0202010D0AAAUAU | BNF | Chlorothiazide 250mg/5ml oral suspension |
| Antihypertensives | 0202010D0AABCBC | BNF | Chlorothiazide 250mg/5ml oral solution |
| Antihypertensives | 0202010D0AABHBH | BNF | Chlorothiazide 40mg/5ml oral liquid |
| Antihypertensives | 0202010D0AABIBI | BNF | Chlorothiazide 200mg/5ml oral solution |
| Antihypertensives | 0202010D0AABJBJ | BNF | Chlorothiazide 30mg/5ml oral liquid |
| Antihypertensives | 0202010D0AABWBW | BNF | Chlorothiazide 125mg/5ml oral liquid |
| Antihypertensives | 0202010D0AACDCD | BNF | Chlorothiazide 225mg/5ml oral liquid |
| Antihypertensives | 0202010D0AACYCY | BNF | Chlorothiazide 250mg tablets |
| Antihypertensives | 0202010D0AADDDD | BNF | Chlorothiazide 150mg/5ml oral suspension |
| Antihypertensives | 0202010D0AADEDE | BNF | Chlorothiazide 25mg/5ml oral suspension |
| Antihypertensives | 0202010D0AADFDF | BNF | Chlorothiazide 300mg capsules |
| Antihypertensives | 0202010D0AADGDG | BNF | Chlorothiazide 200mg/5ml oral suspension |
| Antihypertensives | 0202010D0BCAAAU | BNF | Diuril 250mg/5ml oral suspension |
| Antihypertensives | 0202010F0AAAAAA | BNF | Chlortalidone 50mg tablets |
| Antihypertensives | 0202010F0BBAAAA | BNF | Hygroton 50mg tablets |
| Antihypertensives | 0202010L0AAABAB | BNF | Hydrochlorothiazide 25mg tablets |
| Antihypertensives | 0202010L0AAACAC | BNF | Hydrochlorothiazide 50mg tablets |
| Antihypertensives | 0202010L0AAAJAJ | BNF | Hydrochlorothiazide 125mg/5ml oral liquid |
| Antihypertensives | 0202010L0AAAMAM | BNF | Hydrochlorothiazide 25mg/5ml oral liquid |
| Antihypertensives | 0202010L0AAAPAP | BNF | Hydrochlorothiazide 50mg/5ml oral liquid |
| Antihypertensives | 0202010L0AAASAS | BNF | Hydrochlorothiazide 10mg/5ml oral liquid |
| Antihypertensives | 0202010L0AAATAT | BNF | Hydrochlorothiazide 5mg/5ml oral liquid |
| Antihypertensives | 0202010L0AAAYAY | BNF | Hydrochlorothiazide 125mg tablets |
| Antihypertensives | 0202010L0BCAAAB | BNF | Hydrosaluric 25mg tablets |
| Antihypertensives | 0202010P0AAAAAA | BNF | Indapamide 25mg tablets |
| Antihypertensives | 0202010P0AAABAB | BNF | Indapamide 25mg/5ml oral suspension |
| Antihypertensives | 0202010P0AAADAD | BNF | Indapamide 15mg modified-release tablets |
| Antihypertensives | 0202010P0AAAFAF | BNF | Indapamide 15mg modified-release tablets (old) |
| Antihypertensives | 0202010P0BBAAAA | BNF | Natrilix 25mg tablets |
| Antihypertensives | 0202010P0BBABAD | BNF | Natrilix SR 15mg tablets |
| Antihypertensives | 0202010P0BHAAAD | BNF | Tensaid XL 15mg tablets |
| Antihypertensives | 0202010P0BJAAAD | BNF | Indipam XL 15mg tablets |
| Antihypertensives | 0202010P0BLAAAD | BNF | Rawel XL 15mg tablets |
| Antihypertensives | 0202010P0BMAAAD | BNF | Cardide SR 15mg tablets |
| Antihypertensives | 0202010P0BNAAAD | BNF | Alkapamid XL 15mg tablets (HBS Healthcare) |
| Antihypertensives | 0202010P0BPAAAD | BNF | Alkapamid XL 15mg tablets (Rivopharm) |
| Antihypertensives | 0202010P0BQAAAD | BNF | Lorvacs XL 15mg tablets |
| Antihypertensives | 0202010V0AAAAAA | BNF | Metolazone 5mg tablets |
| Antihypertensives | 0202010V0AAAKAK | BNF | Metolazone 25mg/5ml oral liquid |
| Antihypertensives | 0202010V0AAAMAM | BNF | Metolazone 125mg/5ml oral liquid |
| Antihypertensives | 0202010V0AAANAN | BNF | Metolazone 25mg tablets |
| Antihypertensives | 0202010V0BBAAAA | BNF | Metenix 5mg tablets |
| Antihypertensives | 0202010V0BEAAAN | BNF | Zaroxolyn 25mg tablets |
| Antihypertensives | 0202010V0BEABAA | BNF | Zaroxolyn 5mg tablets |
| Antihypertensives | 0202010Y0AAAAAA | BNF | Xipamide 20mg tablets |
| Antihypertensives | 0202010Y0BBAAAA | BNF | Diurexan 20mg tablets |
| Antihypertensives | 0202030C0AAACAC | BNF | Amiloride 5mg tablets |
| Antihypertensives | 0202030C0AAASAS | BNF | Amiloride 5mg/5ml oral solution sugar free |
| Antihypertensives | 0202030C0AAAXAX | BNF | Amiloride 5mg/5ml oral suspension |
| Antihypertensives | 0202030C0AAAZAZ | BNF | Amiloride 10mg/5ml oral liquid |
| Antihypertensives | 0202030C0BFAAAS | BNF | Amilamont 5mg/5ml oral solution sugar free |
| Antihypertensives | 0202030S0AAATAT | BNF | Spironolactone 25mg tablets |
| Antihypertensives | 0202030S0AAAUAU | BNF | Spironolactone 50mg tablets |
| Antihypertensives | 0202030S0AAAVAV | BNF | Spironolactone 100mg tablets |
| Antihypertensives | 0202030S0AACMCM | BNF | Spironolactone 5mg/5ml oral solution |
| Antihypertensives | 0202030S0AACPCP | BNF | Spironolactone 50mg/5ml oral solution |
| Antihypertensives | 0202030S0AACQCQ | BNF | Spironolactone 10mg/5ml oral solution |
| Antihypertensives | 0202030S0AACRCR | BNF | Spironolactone 100mg/5ml oral solution |
| Antihypertensives | 0202030S0AACTCT | BNF | Spironolactone 625mg/5ml oral liquid |
| Antihypertensives | 0202030S0AACWCW | BNF | Spironolactone 4mg/5ml oral liquid |
| Antihypertensives | 0202030S0AACXCX | BNF | Spironolactone 125mg/5ml oral liquid |
| Antihypertensives | 0202030S0AACYCY | BNF | Spironolactone 7mg/5ml oral liquid |
| Antihypertensives | 0202030S0AACZCZ | BNF | Spironolactone 250mg/5ml oral liquid |
| Antihypertensives | 0202030S0AADADA | BNF | Spironolactone 25mg/5ml oral liquid |
| Antihypertensives | 0202030S0AADCDC | BNF | Spironolactone 15mg/5ml oral suspension |
| Antihypertensives | 0202030S0AADDDD | BNF | Spironolactone 20mg/5ml oral liquid |
| Antihypertensives | 0202030S0AADSDS | BNF | Spironolactone 12mg/5ml oral liquid |
| Antihypertensives | 0202030S0AADUDU | BNF | Spironolactone 2mg/5ml oral liquid |
| Antihypertensives | 0202030S0AAEAEA | BNF | Spironolactone 25mg/5ml oral suspension |
| Antihypertensives | 0202030S0AAEBEB | BNF | Spironolactone 50mg/5ml oral suspension |
| Antihypertensives | 0202030S0AAECEC | BNF | Spironolactone 5mg/5ml oral suspension |
| Antihypertensives | 0202030S0AAEDED | BNF | Spironolactone 10mg/5ml oral suspension |
| Antihypertensives | 0202030S0AAEEEE | BNF | Spironolactone 100mg/5ml oral suspension |
| Antihypertensives | 0202030S0AAEFEF | BNF | Spironolactone 25mg/5ml oral solution |
| Antihypertensives | 0202030S0AAEGEG | BNF | Spironolactone 125mg tablets |
| Antihypertensives | 0202030S0BBAAAT | BNF | Aldactone 25mg tablets |
| Antihypertensives | 0202030S0BBABAU | BNF | Aldactone 50mg tablets |
| Antihypertensives | 0202030S0BBACAV | BNF | Aldactone 100mg tablets |
| Antihypertensives | 0202030W0AAAAAA | BNF | Triamterene 50mg capsules |
| Antihypertensives | 0202030X0AAAAAA | BNF | Eplerenone 25mg tablets |
| Antihypertensives | 0202030X0AAABAB | BNF | Eplerenone 50mg tablets |
| Antihypertensives | 0202030X0BBAAAA | BNF | Inspra 25mg tablets |
| Antihypertensives | 0202030X0BBABAB | BNF | Inspra 50mg tablets |
| Antihypertensives | 020400010AAAAAA | BNF | Pindolol 10mg / Clopamide 5mg tablets |
| Antihypertensives | 020400010BBAAAA | BNF | Viskaldix tablets |
| Antihypertensives | 020400030AAAEAE | BNF | Timolol 10mg / Bendroflumethiazide 25mg tablets |
| Antihypertensives | 020400040AAAAAA | BNF | Co-tenidone 50mg/125mg tablets |
| Antihypertensives | 020400040AAABAB | BNF | Co-tenidone 100mg/25mg tablets |
| Antihypertensives | 020400040BBAAAA | BNF | Tenoret 50mg/125mg tablets |
| Antihypertensives | 020400040BCAAAB | BNF | Tenoretic 100mg/25mg tablets |
| Antihypertensives | 020400060AAAAAA | BNF | Celiprolol 200mg tablets |
| Antihypertensives | 020400060AAABAB | BNF | Celiprolol 400mg tablets |
| Antihypertensives | 020400060BBAAAA | BNF | Celectol 200mg tablets |
| Antihypertensives | 020400060BBABAB | BNF | Celectol 400mg tablets |
| Antihypertensives | 020400080AAABAB | BNF | Carvedilol 125mg tablets |
| Antihypertensives | 020400080AAACAC | BNF | Carvedilol 25mg tablets |
| Antihypertensives | 020400080AAAEAE | BNF | Carvedilol 3125mg tablets |
| Antihypertensives | 020400080AAAFAF | BNF | Carvedilol 625mg tablets |
| Antihypertensives | 020400080AAAPAP | BNF | Carvedilol 5mg/5ml oral suspension |
| Antihypertensives | 0204000ABAAAAAA | BNF | Nebivolol 5mg tablets |
| Antihypertensives | 0204000ABAAABAB | BNF | Nebivolol 25mg tablets |
| Antihypertensives | 0204000ABAAACAC | BNF | Nebivolol 10mg tablets |
| Antihypertensives | 0204000ABAAADAD | BNF | Nebivolol 125mg tablets |
| Antihypertensives | 0204000ABBBAAAA | BNF | Nebilet 5mg tablets |
| Antihypertensives | 0204000ACAAAAAA | BNF | Bisoprolol 5mg / Aspirin 75mg capsules |
| Antihypertensives | 0204000ACAAABAB | BNF | Bisoprolol 10mg / Aspirin 75mg capsules |
| Antihypertensives | 0204000ACAAADAD | BNF | Bisoprolol 5mg / Aspirin 100mg capsules |
| Antihypertensives | 0204000C0AAAAAA | BNF | Acebutolol 100mg capsules |
| Antihypertensives | 0204000C0AAABAB | BNF | Acebutolol 200mg capsules |
| Antihypertensives | 0204000C0AAADAD | BNF | Acebutolol 400mg tablets |
| Antihypertensives | 0204000C0BBAAAA | BNF | Sectral 100mg capsules |
| Antihypertensives | 0204000C0BBABAB | BNF | Sectral 200mg capsules |
| Antihypertensives | 0204000C0BBADAD | BNF | Sectral 400mg tablets |
| Antihypertensives | 0204000E0AAAAAA | BNF | Atenolol 25mg/5ml oral solution sugar free |
| Antihypertensives | 0204000E0AAABAB | BNF | Atenolol 50mg tablets |
| Antihypertensives | 0204000E0AAACAC | BNF | Atenolol 100mg tablets |
| Antihypertensives | 0204000E0AAAGAG | BNF | Atenolol 25mg tablets |
| Antihypertensives | 0204000E0AAANAN | BNF | Atenolol 25mg/5ml oral liquid |
| Antihypertensives | 0204000E0AAASAS | BNF | Atenolol 10mg/5ml oral liquid |
| Antihypertensives | 0204000E0BBABAA | BNF | Tenormin 25mg/5ml syrup |
| Antihypertensives | 0204000E0BBACAC | BNF | Tenormin 100mg tablets |
| Antihypertensives | 0204000E0BBAFAB | BNF | Tenormin LS 50mg tablets |
| Antihypertensives | 0204000H0AAAAAA | BNF | Bisoprolol 5mg tablets |
| Antihypertensives | 0204000H0AAABAB | BNF | Bisoprolol 10mg tablets |
| Antihypertensives | 0204000H0AAAJAJ | BNF | Bisoprolol 25mg tablets |
| Antihypertensives | 0204000H0AAAKAK | BNF | Bisoprolol 375mg tablets |
| Antihypertensives | 0204000H0AAALAL | BNF | Bisoprolol 75mg tablets |
| Antihypertensives | 0204000H0AAAMAM | BNF | Bisoprolol 125mg tablets |
| Antihypertensives | 0204000H0AAARAR | BNF | Bisoprolol 10mg/5ml oral liquid |
| Antihypertensives | 0204000H0AABBBB | BNF | Bisoprolol 75mg/5ml oral liquid |
| Antihypertensives | 0204000H0AABDBD | BNF | Bisoprolol 625micrograms/5ml oral liquid |
| Antihypertensives | 0204000H0AABEBE | BNF | Bisoprolol 25mg/5ml oral solution |
| Antihypertensives | 0204000H0AABFBF | BNF | Bisoprolol 25mg/5ml oral suspension |
| Antihypertensives | 0204000H0AABGBG | BNF | Bisoprolol 5mg/5ml oral solution |
| Antihypertensives | 0204000H0AABHBH | BNF | Bisoprolol 5mg/5ml oral suspension |
| Antihypertensives | 0204000H0AABIBI | BNF | Bisoprolol 125mg/5ml oral suspension |
| Antihypertensives | 0204000H0AABJBJ | BNF | Bisoprolol 125mg/5ml oral solution |
| Antihypertensives | 0204000H0BFAAAM | BNF | Cardicor 125mg tablets |
| Antihypertensives | 0204000H0BFABAJ | BNF | Cardicor 25mg tablets |
| Antihypertensives | 0204000H0BFACAK | BNF | Cardicor 375mg tablets |
| Antihypertensives | 0204000H0BFADAA | BNF | Cardicor 5mg tablets |
| Antihypertensives | 0204000H0BFAEAL | BNF | Cardicor 75mg tablets |
| Antihypertensives | 0204000H0BFAFAB | BNF | Cardicor 10mg tablets |
| Antihypertensives | 0204000H0BKAAAM | BNF | Congescor 125mg tablets (Tillomed) |
| Antihypertensives | 0204000H0BKABAJ | BNF | Congescor 25mg tablets (Tillomed) |
| Antihypertensives | 0204000I0AAAAAA | BNF | Labetalol 100mg/20ml solution for injection ampoules |
| Antihypertensives | 0204000I0AAABAB | BNF | Labetalol 50mg tablets |
| Antihypertensives | 0204000I0AAACAC | BNF | Labetalol 100mg tablets |
| Antihypertensives | 0204000I0AAADAD | BNF | Labetalol 200mg tablets |
| Antihypertensives | 0204000I0AAAEAE | BNF | Labetalol 400mg tablets |
| Antihypertensives | 0204000I0AAAIAI | BNF | Labetalol 50mg/5ml oral liquid |
| Antihypertensives | 0204000I0AAAJAJ | BNF | Labetalol 200mg/5ml oral liquid |
| Antihypertensives | 0204000I0BCAAAB | BNF | Trandate 50mg tablets |
| Antihypertensives | 0204000I0BCABAC | BNF | Trandate 100mg tablets |
| Antihypertensives | 0204000I0BCACAD | BNF | Trandate 200mg tablets |
| Antihypertensives | 0204000I0BCADAE | BNF | Trandate 400mg tablets |
| Antihypertensives | 0204000K0AAAAAA | BNF | Metoprolol 5mg/5ml solution for injection ampoules |
| Antihypertensives | 0204000K0AAABAB | BNF | Metoprolol 50mg tablets |
| Antihypertensives | 0204000K0AAACAC | BNF | Metoprolol 100mg tablets |
| Antihypertensives | 0204000K0AAADAD | BNF | Metoprolol 200mg modified-release tablets |
| Antihypertensives | 0204000K0AAAFAF | BNF | Metoprolol 125mg capsules |
| Antihypertensives | 0204000K0AAAMAM | BNF | Metoprolol 10mg/5ml oral liquid |
| Antihypertensives | 0204000K0AAANAN | BNF | Metoprolol 5mg/5ml oral liquid |
| Antihypertensives | 0204000K0AAASAS | BNF | Metoprolol 25mg/5ml oral liquid |
| Antihypertensives | 0204000K0AAATAT | BNF | Metoprolol 50mg/5ml oral suspension |
| Antihypertensives | 0204000K0AABKBK | BNF | Metoprolol 125mg/5ml oral solution |
| Antihypertensives | 0204000K0AABLBL | BNF | Metoprolol 125mg/5ml oral suspension |
| Antihypertensives | 0204000K0AABMBM | BNF | Metoprolol 50mg/5ml oral solution |
| Antihypertensives | 0204000K0BBAAAB | BNF | Betaloc 50mg tablets |
| Antihypertensives | 0204000K0BBACAA | BNF | Betaloc IV 5mg/5ml solution for injection ampoules |
| Antihypertensives | 0204000K0BDAAAB | BNF | Lopresor 50mg tablets |
| Antihypertensives | 0204000K0BDABAC | BNF | Lopresor 100mg tablets |
| Antihypertensives | 0204000K0BDACAD | BNF | Lopresor SR 200mg tablets |
| Antihypertensives | 0204000M0AAAAAA | BNF | Nadolol 40mg tablets |
| Antihypertensives | 0204000M0AAABAB | BNF | Nadolol 80mg tablets |
| Antihypertensives | 0204000M0AAADAD | BNF | Nadolol 10mg/5ml oral liquid |
| Antihypertensives | 0204000M0AAAFAF | BNF | Nadolol 30mg/5ml oral solution |
| Antihypertensives | 0204000M0AAAGAG | BNF | Nadolol 20mg/5ml oral solution |
| Antihypertensives | 0204000M0AAAHAH | BNF | Nadolol 20mg/5ml oral suspension |
| Antihypertensives | 0204000M0AAAIAI | BNF | Nadolol 30mg/5ml oral suspension |
| Antihypertensives | 0204000M0AAAJAJ | BNF | Nadolol 40mg/5ml oral suspension |
| Antihypertensives | 0204000M0BBABAB | BNF | Corgard 80mg tablets |
| Antihypertensives | 0204000N0AAABAB | BNF | Oxprenolol 20mg tablets |
| Antihypertensives | 0204000N0AAACAC | BNF | Oxprenolol 40mg tablets |
| Antihypertensives | 0204000N0AAAFAF | BNF | Oxprenolol 160mg modified-release tablets |
| Antihypertensives | 0204000N0BEAAAF | BNF | Slow-Trasicor 160mg tablets |
| Antihypertensives | 0204000P0AAAAAA | BNF | Pindolol 5mg tablets |
| Antihypertensives | 0204000P0BBAAAA | BNF | Visken 5mg tablets |
| Antihypertensives | 0204000T0AAABAB | BNF | Sotalol 40mg tablets |
| Antihypertensives | 0204000T0AAACAC | BNF | Sotalol 80mg tablets |
| Antihypertensives | 0204000T0AAAEAE | BNF | Sotalol 160mg tablets |
| Antihypertensives | 0204000T0AAAFAF | BNF | Sotalol 200mg tablets |
| Antihypertensives | 0204000T0AAAMAM | BNF | Sotalol 40mg/5ml oral liquid |
| Antihypertensives | 0204000T0AAARAR | BNF | Sotalol 30mg/5ml oral solution |
| Antihypertensives | 0204000T0AAATAT | BNF | Sotalol 25mg/5ml oral solution |
| Antihypertensives | 0204000T0AABCBC | BNF | Sotalol 25mg/5ml oral suspension |
| Antihypertensives | 0204000T0BBAAAB | BNF | Beta-Cardone 40mg tablets |
| Antihypertensives | 0204000T0BBABAC | BNF | Beta-Cardone 80mg tablets |
| Antihypertensives | 0204000T0BBACAF | BNF | Beta-Cardone 200mg tablets |
| Antihypertensives | 0204000T0BCAAAC | BNF | Sotacor 80mg tablets |
| Antihypertensives | 0204000T0BCABAE | BNF | Sotacor 160mg tablets |
| Antihypertensives | 0204000U0AAAAAA | BNF | Atenolol 50mg / Nifedipine 20mg modified-release capsules |
| Antihypertensives | 0204000U0BBAAAA | BNF | Beta-Adalat modified-release capsules |
| Antihypertensives | 0204000U0BCAAAA | BNF | Tenif 50mg/20mg modified-release capsules |
| Antihypertensives | 0204000V0AAAAAA | BNF | Timolol 10mg tablets |
| Antihypertensives | 0205010AAAAAAAA | BNF | Macitentan 10mg tablets |
| Antihypertensives | 0205010ABAAADAD | BNF | Riociguat 2mg tablets |
| Antihypertensives | 0205010ABAAAEAE | BNF | Riociguat 25mg tablets |
| Antihypertensives | 0205010ABBBAEAE | BNF | Adempas 25mg tablets |
| Antihypertensives | 0205010J0AAA3A3 | BNF | Hydralazine 10mg/5ml oral solution |
| Antihypertensives | 0205010J0AAA4A4 | BNF | Hydralazine 50mg/5ml oral liquid |
| Antihypertensives | 0205010J0AAA8A8 | BNF | Hydralazine 25mg/5ml oral liquid |
| Antihypertensives | 0205010J0AAAGAG | BNF | Hydralazine 25mg tablets |
| Antihypertensives | 0205010J0AAAHAH | BNF | Hydralazine 50mg tablets |
| Antihypertensives | 0205010J0AABKBK | BNF | Hydralazine 10mg tablets |
| Antihypertensives | 0205010J0AABQBQ | BNF | Hydralazine 25mg/5ml oral liquid |
| Antihypertensives | 0205010J0AABTBT | BNF | Hydralazine 10mg/5ml oral suspension |
| Antihypertensives | 0205010J0BBAAAG | BNF | Apresoline 25mg tablets |
| Antihypertensives | 0205010N0AAAAAA | BNF | Minoxidil 25mg tablets |
| Antihypertensives | 0205010N0AAABAB | BNF | Minoxidil 5mg tablets |
| Antihypertensives | 0205010N0AAACAC | BNF | Minoxidil 10mg tablets |
| Antihypertensives | 0205010N0BBAAAA | BNF | Loniten 25mg tablets |
| Antihypertensives | 0205010N0BBABAB | BNF | Loniten 5mg tablets |
| Antihypertensives | 0205010N0BBACAC | BNF | Loniten 10mg tablets |
| Antihypertensives | 0205010U0AAAAAA | BNF | Bosentan 625mg tablets |
| Antihypertensives | 0205010U0AAABAB | BNF | Bosentan 125mg tablets |
| Antihypertensives | 0205010U0BBABAB | BNF | Tracleer 125mg tablets |
| Antihypertensives | 0205010U0BCABAB | BNF | Stayveer 125mg tablets |
| Antihypertensives | 0205010V0AAACAC | BNF | Iloprost 10micrograms/1ml nebuliser liquid ampoules |
| Antihypertensives | 0205010X0AAAAAA | BNF | Ambrisentan 5mg tablets |
| Antihypertensives | 0205010X0AAABAB | BNF | Ambrisentan 10mg tablets |
| Antihypertensives | 0205010X0BBAAAA | BNF | Volibris 5mg tablets |
| Antihypertensives | 0205010X0BBABAB | BNF | Volibris 10mg tablets |
| Antihypertensives | 0205010Y0AAAAAA | BNF | Sildenafil 20mg tablets |
| Antihypertensives | 0205010Y0AAABAB | BNF | Sildenafil 10mg/ml oral suspension sugar free |
| Antihypertensives | 0205010Y0AAACAC | BNF | Sildenafil 10mg/125ml solution for injection vials |
| Antihypertensives | 0205010Y0BBAAAA | BNF | Revatio 20mg tablets |
| Antihypertensives | 0205010Y0BBABAB | BNF | Revatio 10mg/ml oral suspension |
| Antihypertensives | 0205010Y0BBACAC | BNF | Revatio 10mg/125ml solution for injection vials |
| Antihypertensives | 0205010Z0BBAAAA | BNF | Adcirca 20mg tablets |
| Antihypertensives | 0205020E0AAABAB | BNF | Clonidine 150micrograms/1ml solution for injection ampoules |
| Antihypertensives | 0205020E0AAACAC | BNF | Clonidine 100microgram tablets |
| Antihypertensives | 0205020E0AAAFAF | BNF | Clonidine 100micrograms/24hours transdermal patches |
| Antihypertensives | 0205020E0AAAGAG | BNF | Clonidine 200micrograms/24hours transdermal patches |
| Antihypertensives | 0205020E0AAAHAH | BNF | Clonidine 300micrograms/24hours transdermal patches |
| Antihypertensives | 0205020E0BBAAAC | BNF | Catapres 100microgram tablets |
| Antihypertensives | 0205020E0BBADAB | BNF | Catapres 150micrograms/1ml solution for injection ampoules |
| Antihypertensives | 0205020E0BBAEAF | BNF | Catapres TTS 1 patches |
| Antihypertensives | 0205020E0BBAFAG | BNF | Catapres TTS 2 patches |
| Antihypertensives | 0205020E0BBAGAH | BNF | Catapres TTS 3 patches |
| Antihypertensives | 0205020H0AAACAC | BNF | Methyldopa 125mg tablets |
| Antihypertensives | 0205020H0AAADAD | BNF | Methyldopa 250mg tablets |
| Antihypertensives | 0205020H0AAAEAE | BNF | Methyldopa 500mg tablets |
| Antihypertensives | 0205020H0AAAIAI | BNF | Methyldopa 250mg/5ml oral suspension |
| Antihypertensives | 0205020H0BBABAD | BNF | Aldomet 250mg tablets |
| Antihypertensives | 0205020H0BBACAE | BNF | Aldomet 500mg tablets |
| Antihypertensives | 0205020M0AAAAAA | BNF | Moxonidine 200microgram tablets |
| Antihypertensives | 0205020M0AAABAB | BNF | Moxonidine 400microgram tablets |
| Antihypertensives | 0205020M0AAACAC | BNF | Moxonidine 300microgram tablets |
| Antihypertensives | 0205020M0BBAAAA | BNF | Physiotens 200microgram tablets (Mylan) |
| Antihypertensives | 0205020M0BBABAB | BNF | Physiotens 400microgram tablets (Mylan) |
| Antihypertensives | 0205020M0BBACAC | BNF | Physiotens 300microgram tablets (Mylan) |
| Antihypertensives | 0205040D0AAAAAA | BNF | Doxazosin 1mg tablets |
| Antihypertensives | 0205040D0AAABAB | BNF | Doxazosin 2mg tablets |
| Antihypertensives | 0205040D0AAACAC | BNF | Doxazosin 4mg tablets |
| Antihypertensives | 0205040D0AAAHAH | BNF | Doxazosin 8mg/5ml oral liquid |
| Antihypertensives | 0205040D0AAAIAI | BNF | Doxazosin 2mg/5ml oral liquid |
| Antihypertensives | 0205040D0AAAKAK | BNF | Doxazosin 5mg/5ml oral liquid |
| Antihypertensives | 0205040D0AAAQAQ | BNF | Doxazosin 4mg modified-release tablets |
| Antihypertensives | 0205040D0AAARAR | BNF | Doxazosin 8mg modified-release tablets |
| Antihypertensives | 0205040D0AAAXAX | BNF | Doxazosin 4mg/5ml oral solution |
| Antihypertensives | 0205040D0AAAYAY | BNF | Doxazosin 4mg/5ml oral suspension |
| Antihypertensives | 0205040D0AAAZAZ | BNF | Doxazosin 1mg/5ml oral solution |
| Antihypertensives | 0205040D0AABABA | BNF | Doxazosin 1mg/5ml oral suspension |
| Antihypertensives | 0205040D0AABBBB | BNF | Doxazosin 8mg tablets |
| Antihypertensives | 0205040D0BBAAAA | BNF | Cardura 1mg tablets |
| Antihypertensives | 0205040D0BBABAB | BNF | Cardura 2mg tablets |
| Antihypertensives | 0205040D0BBADAQ | BNF | Cardura XL 4mg tablets |
| Antihypertensives | 0205040D0BBAEAR | BNF | Cardura XL 8mg tablets |
| Antihypertensives | 0205040D0BDAAAA | BNF | Doxadura 1mg tablets |
| Antihypertensives | 0205040D0BDABAB | BNF | Doxadura 2mg tablets |
| Antihypertensives | 0205040D0BDACAC | BNF | Doxadura 4mg tablets |
| Antihypertensives | 0205040D0BDADAQ | BNF | Doxadura XL 4mg tablets |
| Antihypertensives | 0205040D0BEAAAQ | BNF | Slocinx XL 4mg tablets |
| Antihypertensives | 0205040D0BGABAQ | BNF | Cardozin XL 4mg tablets (Almus) |
| Antihypertensives | 0205040D0BIAAAQ | BNF | Doxzogen XL 4mg tablets |
| Antihypertensives | 0205040D0BJAAAQ | BNF | Larbex XL 4mg tablets |
| Antihypertensives | 0205040D0BKAAAQ | BNF | Raporsin XL 4mg tablets (Actavis) |
| Antihypertensives | 0205040I0AAAAAA | BNF | Indoramin 25mg tablets |
| Antihypertensives | 0205040M0AAACAC | BNF | Phenoxybenzamine 10mg capsules |
| Antihypertensives | 0205040S0AAABAB | BNF | Prazosin 500microgram tablets |
| Antihypertensives | 0205040S0AAACAC | BNF | Prazosin 1mg tablets |
| Antihypertensives | 0205040S0AAADAD | BNF | Prazosin 2mg tablets |
| Antihypertensives | 0205040S0AAAEAE | BNF | Prazosin 5mg tablets |
| Antihypertensives | 0205040S0AAAIAI | BNF | Prazosin 500micrograms/5ml oral liquid |
| Antihypertensives | 0205040S0AAAKAK | BNF | Prazosin 1mg/5ml oral liquid |
| Antihypertensives | 0205040S0BBAAAB | BNF | Hypovase 500microgram tablets |
| Antihypertensives | 0205040S0BBABAC | BNF | Hypovase 1mg tablets |
| Antihypertensives | 0205040S0BCABAD | BNF | Minipress 2mg tablets |
| Antihypertensives | 0205040V0AAAAAA | BNF | Terazosin 2mg tablets and Terazosin 1mg tablets |
| Antihypertensives | 0205040V0AAABAB | BNF | Terazosin 2mg tablets |
| Antihypertensives | 0205040V0AAACAC | BNF | Terazosin 5mg tablets |
| Antihypertensives | 0205040V0AAADAD | BNF | Terazosin 10mg tablets |
| Antihypertensives | 0205040V0BBABAB | BNF | Hytrin 2mg tablets |
| Antihypertensives | 0205040V0BBACAC | BNF | Hytrin 5mg tablets |
| Antihypertensives | 0205040V0BBADAD | BNF | Hytrin 10mg tablets |
| Antihypertensives | 0205040V0BBAEAA | BNF | Hytrin tablets starter pack |
| Antihypertensives | 0205040V0BCAAAB | BNF | Benph 2mg tablets |
| Antihypertensives | 0205051AAAAABAB | BNF | Perindopril tosilate 5mg tablets |
| Antihypertensives | 0205051AAAAACAC | BNF | Perindopril tosilate 10mg tablets |
| Antihypertensives | 0205051ABAAAAAA | BNF | Perindopril tosilate 5mg / Indapamide 125mg tablets |
| Antihypertensives | 0205051ACAAAAAA | BNF | Perindopril erbumine 4mg / Amlodipine 5mg tablets |
| Antihypertensives | 0205051ACAAABAB | BNF | Perindopril erbumine 4mg / Amlodipine 10mg tablets |
| Antihypertensives | 0205051ACAAACAC | BNF | Perindopril erbumine 8mg / Amlodipine 10mg tablets |
| Antihypertensives | 0205051ACAAADAD | BNF | Perindopril erbumine 8mg / Amlodipine 5mg tablets |
| Antihypertensives | 0205051E0AAAEAE | BNF | Cilazapril 5mg tablets |
| Antihypertensives | 0205051F0AAADAD | BNF | Captopril 125mg tablets |
| Antihypertensives | 0205051F0AAAEAE | BNF | Captopril 25mg tablets |
| Antihypertensives | 0205051F0AAAFAF | BNF | Captopril 50mg tablets |
| Antihypertensives | 0205051F0AABNBN | BNF | Captopril 5mg/5ml oral liquid |
| Antihypertensives | 0205051F0AABPBP | BNF | Captopril 20mg/5ml oral liquid |
| Antihypertensives | 0205051F0AABRBR | BNF | Captopril 10mg/5ml oral liquid |
| Antihypertensives | 0205051F0AABTBT | BNF | Captopril 125mg/5ml oral liquid |
| Antihypertensives | 0205051F0AABWBW | BNF | Captopril 25mg/5ml oral liquid |
| Antihypertensives | 0205051F0AABXBX | BNF | Captopril 625mg/5ml oral liquid |
| Antihypertensives | 0205051F0AABYBY | BNF | Captopril 25mg/5ml oral liquid |
| Antihypertensives | 0205051F0AACACA | BNF | Captopril 50mg/5ml oral liquid |
| Antihypertensives | 0205051F0AACDCD | BNF | Captopril 3mg/5ml oral liquid |
| Antihypertensives | 0205051F0AACICI | BNF | Captopril 3125mg/5ml oral liquid |
| Antihypertensives | 0205051F0AACTCT | BNF | Captopril 5mg/ml oral solution sugar free |
| Antihypertensives | 0205051F0AADFDF | BNF | Captopril 12mg/5ml oral solution |
| Antihypertensives | 0205051F0AADUDU | BNF | Captopril 50mg capsules |
| Antihypertensives | 0205051F0AADZDZ | BNF | Captopril 25mg/5ml oral solution sugar free |
| Antihypertensives | 0205051F0AAEAEA | BNF | Captopril 5mg/5ml oral solution sugar free |
| Antihypertensives | 0205051F0BCABAE | BNF | Capoten 25mg tablets |
| Antihypertensives | 0205051F0BCACAF | BNF | Capoten 50mg tablets |
| Antihypertensives | 0205051F0BDABAE | BNF | Ecopace 25mg tablets |
| Antihypertensives | 0205051F0BIAADZ | BNF | Noyada 25mg/5ml oral solution |
| Antihypertensives | 0205051F0BIABEA | BNF | Noyada 5mg/5ml oral solution |
| Antihypertensives | 0205051G0AAAAAA | BNF | Co-zidocapt 25mg/50mg tablets |
| Antihypertensives | 0205051G0AAABAB | BNF | Co-zidocapt 125mg/25mg tablets |
| Antihypertensives | 0205051G0BCAAAA | BNF | Capozide 25mg/50mg tablets |
| Antihypertensives | 0205051H0AAAAAA | BNF | Enalapril 20mg / Hydrochlorothiazide 125mg tablets |
| Antihypertensives | 0205051H0BBAAAA | BNF | Innozide 20mg/125mg tablets |
| Antihypertensives | 0205051I0AAAAAA | BNF | Enalapril 25mg tablets |
| Antihypertensives | 0205051I0AAABAB | BNF | Enalapril 5mg tablets |
| Antihypertensives | 0205051I0AAACAC | BNF | Enalapril 10mg tablets |
| Antihypertensives | 0205051I0AAADAD | BNF | Enalapril 20mg tablets |
| Antihypertensives | 0205051I0AAAJAJ | BNF | Enalapril 25mg/5ml oral liquid |
| Antihypertensives | 0205051I0AAAUAU | BNF | Enalapril 10mg/5ml oral solution |
| Antihypertensives | 0205051I0AABFBF | BNF | Enalapril 125mg/5ml oral solution |
| Antihypertensives | 0205051I0AABSBS | BNF | Enalapril 15mg/5ml oral liquid |
| Antihypertensives | 0205051I0AABTBT | BNF | Enalapril 12mg/5ml oral liquid |
| Antihypertensives | 0205051I0AABYBY | BNF | Enalapril 5mg/5ml oral solution |
| Antihypertensives | 0205051I0AABZBZ | BNF | Enalapril 5mg/5ml oral suspension |
| Antihypertensives | 0205051I0AACACA | BNF | Enalapril 10mg/5ml oral suspension |
| Antihypertensives | 0205051I0AACBCB | BNF | Enalapril 125mg/5ml oral suspension |
| Antihypertensives | 0205051I0BBAAAA | BNF | Innovace 25mg tablets |
| Antihypertensives | 0205051I0BBABAB | BNF | Innovace 5mg tablets |
| Antihypertensives | 0205051I0BBACAC | BNF | Innovace 10mg tablets |
| Antihypertensives | 0205051I0BBADAD | BNF | Innovace 20mg tablets |
| Antihypertensives | 0205051J0AAAAAA | BNF | Fosinopril 10mg tablets |
| Antihypertensives | 0205051J0AAABAB | BNF | Fosinopril 20mg tablets |
| Antihypertensives | 0205051J0BBABAB | BNF | Staril 20mg tablets |
| Antihypertensives | 0205051K0AAAAAA | BNF | Lisinopril 20mg / Hydrochlorothiazide 125mg tablets |
| Antihypertensives | 0205051K0AAABAB | BNF | Lisinopril 10mg / Hydrochlorothiazide 125mg tablets |
| Antihypertensives | 0205051K0BBAAAA | BNF | Zestoretic 20 tablets |
| Antihypertensives | 0205051K0BBABAB | BNF | Zestoretic 10 tablets |
| Antihypertensives | 0205051K0BCAAAA | BNF | Carace 20 Plus tablets |
| Antihypertensives | 0205051K0BDABAA | BNF | Caralpha 20mg/125mg tablets |
| Antihypertensives | 0205051K0BFAAAA | BNF | Lisoretic 20mg/125mg tablets |
| Antihypertensives | 0205051K0BFABAB | BNF | Lisoretic 10mg/125mg tablets |
| Antihypertensives | 0205051L0AAAAAA | BNF | Lisinopril 25mg tablets |
| Antihypertensives | 0205051L0AAABAB | BNF | Lisinopril 5mg tablets |
| Antihypertensives | 0205051L0AAACAC | BNF | Lisinopril 10mg tablets |
| Antihypertensives | 0205051L0AAADAD | BNF | Lisinopril 20mg tablets |
| Antihypertensives | 0205051L0AAAGAG | BNF | Lisinopril 5mg/5ml oral liquid |
| Antihypertensives | 0205051L0AAAHAH | BNF | Lisinopril 10mg/5ml oral liquid |
| Antihypertensives | 0205051L0AAAIAI | BNF | Lisinopril 25mg/5ml oral liquid |
| Antihypertensives | 0205051L0AAAKAK | BNF | Lisinopril 15mg/5ml oral liquid |
| Antihypertensives | 0205051L0AAANAN | BNF | Lisinopril 75mg/5ml oral liquid |
| Antihypertensives | 0205051L0AAAQAQ | BNF | Lisinopril 5mg/5ml oral solution sugar free |
| Antihypertensives | 0205051L0AAAYAY | BNF | Lisinopril 20mg/5ml oral solution |
| Antihypertensives | 0205051L0AAAZAZ | BNF | Lisinopril 20mg/5ml oral suspension |
| Antihypertensives | 0205051L0BBAAAA | BNF | Zestril 25mg tablets |
| Antihypertensives | 0205051L0BBABAB | BNF | Zestril 5mg tablets |
| Antihypertensives | 0205051L0BBACAC | BNF | Zestril 10mg tablets |
| Antihypertensives | 0205051L0BBADAD | BNF | Zestril 20mg tablets |
| Antihypertensives | 0205051L0BCACAC | BNF | Carace 10mg tablets |
| Antihypertensives | 0205051L0BEAAAA | BNF | Lisopress 25mg tablets |
| Antihypertensives | 0205051L0BEACAC | BNF | Lisopress 10mg tablets |
| Antihypertensives | 0205051M0AAAAAA | BNF | Perindopril erbumine 2mg tablets |
| Antihypertensives | 0205051M0AAABAB | BNF | Perindopril erbumine 4mg tablets |
| Antihypertensives | 0205051M0AAACAC | BNF | Perindopril erbumine 1mg/5ml oral liquid |
| Antihypertensives | 0205051M0AAAFAF | BNF | Perindopril erbumine 8mg tablets |
| Antihypertensives | 0205051M0AAAHAH | BNF | Perindopril erbumine 8mg/5ml oral liquid |
| Antihypertensives | 0205051M0AAAKAK | BNF | Perindopril erbumine 4mg/5ml oral solution |
| Antihypertensives | 0205051M0AAALAL | BNF | Perindopril erbumine 4mg/5ml oral suspension |
| Antihypertensives | 0205051M0BBAAAA | BNF | Coversyl 2mg tablets |
| Antihypertensives | 0205051M0BBABAB | BNF | Coversyl 4mg tablets |
| Antihypertensives | 0205051M0BBACAF | BNF | Coversyl 8mg tablets |
| Antihypertensives | 0205051N0AAAAAA | BNF | Perindopril erbumine 4mg / Indapamide 125mg tablets |
| Antihypertensives | 0205051N0BBAAAA | BNF | Coversyl Plus tablets |
| Antihypertensives | 0205051P0AAAAAA | BNF | Quinapril 10mg / Hydrochlorothiazide 125mg tablets |
| Antihypertensives | 0205051P0BBAAAA | BNF | Accuretic 10mg/125mg tablets |
| Antihypertensives | 0205051Q0AAAAAA | BNF | Quinapril 5mg tablets |
| Antihypertensives | 0205051Q0AAABAB | BNF | Quinapril 10mg tablets |
| Antihypertensives | 0205051Q0AAACAC | BNF | Quinapril 20mg tablets |
| Antihypertensives | 0205051Q0AAADAD | BNF | Quinapril 40mg tablets |
| Antihypertensives | 0205051Q0BBAAAA | BNF | Accupro 5mg tablets |
| Antihypertensives | 0205051Q0BBABAB | BNF | Accupro 10mg tablets |
| Antihypertensives | 0205051Q0BBACAC | BNF | Accupro 20mg tablets |
| Antihypertensives | 0205051Q0BBADAD | BNF | Accupro 40mg tablets |
| Antihypertensives | 0205051R0AAAAAA | BNF | Ramipril 125mg capsules |
| Antihypertensives | 0205051R0AAABAB | BNF | Ramipril 25mg capsules |
| Antihypertensives | 0205051R0AAACAC | BNF | Ramipril 5mg capsules |
| Antihypertensives | 0205051R0AAADAD | BNF | Ramipril 10mg capsules |
| Antihypertensives | 0205051R0AAAEAE | BNF | Ramipril 5mg/5ml oral liquid |
| Antihypertensives | 0205051R0AAAFAF | BNF | Ramipril 25mg/5ml oral liquid |
| Antihypertensives | 0205051R0AAAGAG | BNF | Ramipril 125mg/5ml oral liquid |
| Antihypertensives | 0205051R0AAAHAH | BNF | Ramipril 10mg/5ml oral liquid |
| Antihypertensives | 0205051R0AAAKAK | BNF | Ramipril 125mg tablets |
| Antihypertensives | 0205051R0AAALAL | BNF | Ramipril 25mg tablets |
| Antihypertensives | 0205051R0AAAMAM | BNF | Ramipril 5mg tablets |
| Antihypertensives | 0205051R0AAANAN | BNF | Ramipril 10mg tablets |
| Antihypertensives | 0205051R0AAATAT | BNF | Ramipril 25mg/5ml oral solution sugar free |
| Antihypertensives | 0205051R0AAAUAU | BNF | Generic Tritace titration pack tablets |
| Antihypertensives | 0205051R0BBADAD | BNF | Tritace 10mg capsules |
| Antihypertensives | 0205051R0BBAFAK | BNF | Tritace 125mg tablets |
| Antihypertensives | 0205051R0BBAGAL | BNF | Tritace 25mg tablets |
| Antihypertensives | 0205051R0BBAHAM | BNF | Tritace 5mg tablets |
| Antihypertensives | 0205051R0BBAIAN | BNF | Tritace 10mg tablets |
| Antihypertensives | 0205051R0BBAJAU | BNF | Tritace titration pack tablets |
| Antihypertensives | 0205051S0AAAAAA | BNF | Felodipine 25mg modified-release / Ramipril 25mg tablets |
| Antihypertensives | 0205051S0AAABAB | BNF | Felodipine 5mg modified-release / Ramipril 5mg tablets |
| Antihypertensives | 0205051S0BBAAAA | BNF | Triapin 25mg/25mg modified-release tablets |
| Antihypertensives | 0205051S0BBABAB | BNF | Triapin 5mg/5mg modified-release tablets |
| Antihypertensives | 0205051U0AAAAAA | BNF | Trandolapril 500microgram capsules |
| Antihypertensives | 0205051U0AAABAB | BNF | Trandolapril 1mg capsules |
| Antihypertensives | 0205051U0AAACAC | BNF | Trandolapril 2mg capsules |
| Antihypertensives | 0205051U0AAAFAF | BNF | Trandolapril 4mg capsules |
| Antihypertensives | 0205051W0AAAAAA | BNF | Imidapril 10mg tablets |
| Antihypertensives | 0205051W0AAABAB | BNF | Imidapril 5mg tablets |
| Antihypertensives | 0205051W0AAACAC | BNF | Imidapril 20mg tablets |
| Antihypertensives | 0205051W0BBAAAA | BNF | Tanatril 10mg tablets |
| Antihypertensives | 0205051W0BBABAB | BNF | Tanatril 5mg tablets |
| Antihypertensives | 0205051W0BBACAC | BNF | Tanatril 20mg tablets |
| Antihypertensives | 0205051Y0AAAAAA | BNF | Perindopril arginine 25mg tablets |
| Antihypertensives | 0205051Y0AAABAB | BNF | Perindopril arginine 5mg tablets |
| Antihypertensives | 0205051Y0AAACAC | BNF | Perindopril arginine 10mg tablets |
| Antihypertensives | 0205051Y0BBAAAA | BNF | Coversyl Arginine 25mg tablets |
| Antihypertensives | 0205051Y0BBABAB | BNF | Coversyl Arginine 5mg tablets |
| Antihypertensives | 0205051Y0BBACAC | BNF | Coversyl Arginine 10mg tablets |
| Antihypertensives | 0205051Z0AAAAAA | BNF | Perindopril arginine 5mg / Indapamide 125mg tablets |
| Antihypertensives | 0205051Z0BBAAAA | BNF | Coversyl Arginine Plus 5mg/125mg tablets |
| Antihypertensives | 0205052A0AAAAAA | BNF | Irbesartan 150mg / Hydrochlorothiazide 125mg tablets |
| Antihypertensives | 0205052A0AAABAB | BNF | Irbesartan 300mg / Hydrochlorothiazide 125mg tablets |
| Antihypertensives | 0205052A0AAACAC | BNF | Irbesartan 300mg / Hydrochlorothiazide 25mg tablets |
| Antihypertensives | 0205052A0BBAAAA | BNF | CoAprovel 150mg/125mg tablets |
| Antihypertensives | 0205052A0BBABAB | BNF | CoAprovel 300mg/125mg tablets |
| Antihypertensives | 0205052A0BBACAC | BNF | CoAprovel 300mg/25mg tablets |
| Antihypertensives | 0205052ABAAAAAA | BNF | Olmesartan medoxomil 20mg / Amlodipine 5mg tablets |
| Antihypertensives | 0205052ABAAABAB | BNF | Olmesartan medoxomil 40mg / Amlodipine 5mg tablets |
| Antihypertensives | 0205052ABAAACAC | BNF | Olmesartan medoxomil 40mg / Amlodipine 10mg tablets |
| Antihypertensives | 0205052ABBBAAAA | BNF | Sevikar 20mg/5mg tablets |
| Antihypertensives | 0205052ABBBABAB | BNF | Sevikar 40mg/5mg tablets |
| Antihypertensives | 0205052ABBBACAC | BNF | Sevikar 40mg/10mg tablets |
| Antihypertensives | 0205052ACAAAAAA | BNF | Generic Sevikar HCT 20mg/5mg/125mg tablets |
| Antihypertensives | 0205052ACAAABAB | BNF | Generic Sevikar HCT 40mg/5mg/125mg tablets |
| Antihypertensives | 0205052ACAAACAC | BNF | Generic Sevikar HCT 40mg/10mg/125mg tablets |
| Antihypertensives | 0205052ACAAADAD | BNF | Generic Sevikar HCT 40mg/5mg/25mg tablets |
| Antihypertensives | 0205052ACAAAEAE | BNF | Generic Sevikar HCT 40mg/10mg/25mg tablets |
| Antihypertensives | 0205052ACBBAAAA | BNF | Sevikar HCT 20mg/5mg/125mg tablets |
| Antihypertensives | 0205052ACBBABAB | BNF | Sevikar HCT 40mg/5mg/125mg tablets |
| Antihypertensives | 0205052ACBBACAC | BNF | Sevikar HCT 40mg/10mg/125mg tablets |
| Antihypertensives | 0205052ACBBADAD | BNF | Sevikar HCT 40mg/5mg/25mg tablets |
| Antihypertensives | 0205052ACBBAEAE | BNF | Sevikar HCT 40mg/10mg/25mg tablets |
| Antihypertensives | 0205052ADAAAAAA | BNF | Azilsartan medoxomil 20mg tablets |
| Antihypertensives | 0205052ADAAABAB | BNF | Azilsartan medoxomil 40mg tablets |
| Antihypertensives | 0205052ADAAACAC | BNF | Azilsartan medoxomil 80mg tablets |
| Antihypertensives | 0205052ADBBAAAA | BNF | Edarbi 20mg tablets |
| Antihypertensives | 0205052ADBBABAB | BNF | Edarbi 40mg tablets |
| Antihypertensives | 0205052ADBBACAC | BNF | Edarbi 80mg tablets |
| Antihypertensives | 0205052AEAAAAAA | BNF | Sacubitril 49mg / Valsartan 51mg tablets |
| Antihypertensives | 0205052AEAAABAB | BNF | Sacubitril 97mg / Valsartan 103mg tablets |
| Antihypertensives | 0205052AEAAACAC | BNF | Sacubitril 24mg / Valsartan 26mg tablets |
| Antihypertensives | 0205052AEBBAAAA | BNF | Entresto 49mg/51mg tablets |
| Antihypertensives | 0205052AEBBABAB | BNF | Entresto 97mg/103mg tablets |
| Antihypertensives | 0205052AEBBACAC | BNF | Entresto 24mg/26mg tablets |
| Antihypertensives | 0205052B0AAAAAA | BNF | Olmesartan medoxomil 10mg tablets |
| Antihypertensives | 0205052B0AAABAB | BNF | Olmesartan medoxomil 20mg tablets |
| Antihypertensives | 0205052B0AAACAC | BNF | Olmesartan medoxomil 40mg tablets |
| Antihypertensives | 0205052B0AAADAD | BNF | Olmesartan medoxomil 10mg/5ml oral liquid |
| Antihypertensives | 0205052B0BBAAAA | BNF | Olmetec 10mg tablets |
| Antihypertensives | 0205052B0BBABAB | BNF | Olmetec 20mg tablets |
| Antihypertensives | 0205052B0BBACAC | BNF | Olmetec 40mg tablets |
| Antihypertensives | 0205052C0AAAAAA | BNF | Candesartan 2mg tablets |
| Antihypertensives | 0205052C0AAABAB | BNF | Candesartan 4mg tablets |
| Antihypertensives | 0205052C0AAACAC | BNF | Candesartan 8mg tablets |
| Antihypertensives | 0205052C0AAADAD | BNF | Candesartan 16mg tablets |
| Antihypertensives | 0205052C0AAAHAH | BNF | Candesartan 32mg tablets |
| Antihypertensives | 0205052C0AAAJAJ | BNF | Candesartan 8mg/5ml oral liquid |
| Antihypertensives | 0205052C0BBAAAA | BNF | Amias 2mg tablets |
| Antihypertensives | 0205052C0BBABAB | BNF | Amias 4mg tablets |
| Antihypertensives | 0205052C0BBACAC | BNF | Amias 8mg tablets |
| Antihypertensives | 0205052C0BBADAD | BNF | Amias 16mg tablets |
| Antihypertensives | 0205052C0BBAEAH | BNF | Amias 32mg tablets |
| Antihypertensives | 0205052I0AAAAAA | BNF | Irbesartan 75mg tablets |
| Antihypertensives | 0205052I0AAABAB | BNF | Irbesartan 150mg tablets |
| Antihypertensives | 0205052I0AAACAC | BNF | Irbesartan 300mg tablets |
| Antihypertensives | 0205052I0AAAFAF | BNF | Irbesartan 150mg/5ml oral suspension |
| Antihypertensives | 0205052I0AAAGAG | BNF | Irbesartan 300mg/5ml oral suspension |
| Antihypertensives | 0205052I0AAAMAM | BNF | Irbesartan 100mg/5ml oral suspension |
| Antihypertensives | 0205052I0BBAAAA | BNF | Aprovel 75mg tablets |
| Antihypertensives | 0205052I0BBABAB | BNF | Aprovel 150mg tablets |
| Antihypertensives | 0205052I0BBACAC | BNF | Aprovel 300mg tablets |
| Antihypertensives | 0205052I0BEAAAA | BNF | Ifirmasta 75mg tablets |
| Antihypertensives | 0205052I0BEABAB | BNF | Ifirmasta 150mg tablets |
| Antihypertensives | 0205052I0BEACAC | BNF | Ifirmasta 300mg tablets |
| Antihypertensives | 0205052N0AAAAAA | BNF | Losartan 25mg tablets |
| Antihypertensives | 0205052N0AAABAB | BNF | Losartan 50mg tablets |
| Antihypertensives | 0205052N0AAADAD | BNF | Losartan 100mg tablets |
| Antihypertensives | 0205052N0AAAEAE | BNF | Losartan 50mg/5ml oral solution |
| Antihypertensives | 0205052N0AAAFAF | BNF | Losartan 100mg/5ml oral liquid |
| Antihypertensives | 0205052N0AAAGAG | BNF | Losartan 25mg/5ml oral liquid |
| Antihypertensives | 0205052N0AAAHAH | BNF | Losartan 125mg tablets |
| Antihypertensives | 0205052N0AAAIAI | BNF | Losartan 25mg/ml oral suspension sugar free |
| Antihypertensives | 0205052N0AAAJAJ | BNF | Losartan 50mg/5ml oral suspension |
| Antihypertensives | 0205052N0AAALAL | BNF | Losartan 125mg/5ml oral suspension |
| Antihypertensives | 0205052N0BBAAAA | BNF | Cozaar 25mg tablets |
| Antihypertensives | 0205052N0BBABAB | BNF | Cozaar 50mg tablets |
| Antihypertensives | 0205052N0BBACAD | BNF | Cozaar 100mg tablets |
| Antihypertensives | 0205052N0BBADAH | BNF | Cozaar 125mg tablets |
| Antihypertensives | 0205052N0BBAEAI | BNF | Cozaar 25mg/ml oral suspension |
| Antihypertensives | 0205052P0AAAAAA | BNF | Losartan 50mg / Hydrochlorothiazide 125mg tablets |
| Antihypertensives | 0205052P0AAABAB | BNF | Losartan 100mg / Hydrochlorothiazide 25mg tablets |
| Antihypertensives | 0205052P0AAACAC | BNF | Losartan 100mg / Hydrochlorothiazide 125mg tablets |
| Antihypertensives | 0205052P0BBAAAA | BNF | Cozaar-Comp 50mg/125mg tablets |
| Antihypertensives | 0205052P0BBABAB | BNF | Cozaar-Comp 100mg/25mg tablets |
| Antihypertensives | 0205052P0BBACAC | BNF | Cozaar-Comp 100mg/125mg tablets |
| Antihypertensives | 0205052Q0AAAAAA | BNF | Telmisartan 40mg tablets |
| Antihypertensives | 0205052Q0AAABAB | BNF | Telmisartan 80mg tablets |
| Antihypertensives | 0205052Q0AAACAC | BNF | Telmisartan 20mg tablets |
| Antihypertensives | 0205052Q0BBAAAA | BNF | Micardis 40mg tablets |
| Antihypertensives | 0205052Q0BBABAB | BNF | Micardis 80mg tablets |
| Antihypertensives | 0205052Q0BBACAC | BNF | Micardis 20mg tablets |
| Antihypertensives | 0205052Q0BCAAAA | BNF | Tolura 40mg tablets |
| Antihypertensives | 0205052Q0BCABAB | BNF | Tolura 80mg tablets |
| Antihypertensives | 0205052Q0BCACAC | BNF | Tolura 20mg tablets |
| Antihypertensives | 0205052R0AAAAAA | BNF | Telmisartan 40mg / Hydrochlorothiazide 125mg tablets |
| Antihypertensives | 0205052R0AAABAB | BNF | Telmisartan 80mg / Hydrochlorothiazide 125mg tablets |
| Antihypertensives | 0205052R0AAACAC | BNF | Telmisartan 80mg / Hydrochlorothiazide 25mg tablets |
| Antihypertensives | 0205052R0BBAAAA | BNF | MicardisPlus 40mg/125mg tablets |
| Antihypertensives | 0205052R0BBABAB | BNF | MicardisPlus 80mg/125mg tablets |
| Antihypertensives | 0205052R0BBACAC | BNF | MicardisPlus 80mg/25mg tablets |
| Antihypertensives | 0205052R0BCAAAA | BNF | Actelsar HCT 40mg/125mg tablets |
| Antihypertensives | 0205052R0BCABAB | BNF | Actelsar HCT 80mg/125mg tablets |
| Antihypertensives | 0205052R0BCACAC | BNF | Actelsar HCT 80mg/25mg tablets |
| Antihypertensives | 0205052R0BDAAAA | BNF | Tolucombi 40mg/125mg tablets |
| Antihypertensives | 0205052R0BDABAB | BNF | Tolucombi 80mg/125mg tablets |
| Antihypertensives | 0205052R0BDACAC | BNF | Tolucombi 80mg/25mg tablets |
| Antihypertensives | 0205052V0AAAAAA | BNF | Valsartan 40mg capsules |
| Antihypertensives | 0205052V0AAABAB | BNF | Valsartan 80mg capsules |
| Antihypertensives | 0205052V0AAACAC | BNF | Valsartan 160mg capsules |
| Antihypertensives | 0205052V0AAADAD | BNF | Valsartan 40mg tablets |
| Antihypertensives | 0205052V0AAAEAE | BNF | Valsartan 40mg/5ml oral liquid |
| Antihypertensives | 0205052V0AAAFAF | BNF | Valsartan 320mg tablets |
| Antihypertensives | 0205052V0AAAHAH | BNF | Valsartan 160mg tablets |
| Antihypertensives | 0205052V0AAAIAI | BNF | Valsartan 80mg tablets |
| Antihypertensives | 0205052V0AAAJAJ | BNF | Valsartan 3mg/ml oral solution |
| Antihypertensives | 0205052V0BBAAAA | BNF | Diovan 40mg capsules |
| Antihypertensives | 0205052V0BBABAB | BNF | Diovan 80mg capsules |
| Antihypertensives | 0205052V0BBACAC | BNF | Diovan 160mg capsules |
| Antihypertensives | 0205052V0BBADAD | BNF | Diovan 40mg tablets |
| Antihypertensives | 0205052V0BBAEAF | BNF | Diovan 320mg tablets |
| Antihypertensives | 0205052V0BBAFAJ | BNF | Diovan 3mg/1ml oral solution |
| Antihypertensives | 0205052W0AAAAAA | BNF | Eprosartan 300mg tablets |
| Antihypertensives | 0205052W0AAABAB | BNF | Eprosartan 400mg tablets |
| Antihypertensives | 0205052W0AAACAC | BNF | Eprosartan 600mg tablets |
| Antihypertensives | 0205052W0BBAAAA | BNF | Teveten 300mg tablets |
| Antihypertensives | 0205052W0BBABAB | BNF | Teveten 400mg tablets |
| Antihypertensives | 0205052W0BBACAC | BNF | Teveten 600mg tablets |
| Antihypertensives | 0205052X0AAAAAA | BNF | Valsartan 160mg / Hydrochlorothiazide 125mg tablets |
| Antihypertensives | 0205052X0AAABAB | BNF | Valsartan 160mg / Hydrochlorothiazide 25mg tablets |
| Antihypertensives | 0205052X0AAACAC | BNF | Valsartan 80mg / Hydrochlorothiazide 125mg tablets |
| Antihypertensives | 0205052X0BBAAAA | BNF | Co-Diovan 160mg/125mg tablets |
| Antihypertensives | 0205052X0BBABAB | BNF | Co-Diovan 160mg/25mg tablets |
| Antihypertensives | 0205052X0BBACAC | BNF | Co-Diovan 80mg/125mg tablets |
| Antihypertensives | 0205052Y0AAAAAA | BNF | Olmesartan medoxomil 20mg / Hydrochlorothiazide 125mg tab |
| Antihypertensives | 0205052Y0AAABAB | BNF | Olmesartan medoxomil 20mg / Hydrochlorothiazide 25mg tablets |
| Antihypertensives | 0205052Y0AAACAC | BNF | Olmesartan medoxomil 40mg / Hydrochlorothiazide 125mg tab |
| Antihypertensives | 0205052Y0BBAAAA | BNF | Olmetec Plus 20mg/125mg tablets |
| Antihypertensives | 0205052Y0BBABAB | BNF | Olmetec Plus 20mg/25mg tablets |
| Antihypertensives | 0205052Y0BBACAC | BNF | Olmetec Plus 40mg/125mg tablets |
| Antihypertensives | 0205053A0AAAAAA | BNF | Aliskiren 150mg tablets |
| Antihypertensives | 0205053A0AAABAB | BNF | Aliskiren 300mg tablets |
| Antihypertensives | 0205053A0BBAAAA | BNF | Rasilez 150mg tablets |
| Antihypertensives | 0205053A0BBABAB | BNF | Rasilez 300mg tablets |
| Antihypertensives | 0206020A0AAAAAA | BNF | Amlodipine 5mg tablets |
| Antihypertensives | 0206020A0AAABAB | BNF | Amlodipine 10mg tablets |
| Antihypertensives | 0206020A0AAACAC | BNF | Amlodipine 5mg/5ml oral liquid |
| Antihypertensives | 0206020A0AAADAD | BNF | Amlodipine 10mg/5ml oral liquid |
| Antihypertensives | 0206020A0AAAFAF | BNF | Amlodipine 50mg/5ml oral liquid |
| Antihypertensives | 0206020A0AAAJAJ | BNF | Amlodipine 25mg/5ml oral suspension |
| Antihypertensives | 0206020A0AAALAL | BNF | Amlodipine 5mg/5ml oral solution sugar free |
| Antihypertensives | 0206020A0AAAMAM | BNF | Amlodipine 15mg/5ml oral liquid |
| Antihypertensives | 0206020A0AAANAN | BNF | Amlodipine 2mg/5ml oral liquid |
| Antihypertensives | 0206020A0AAATAT | BNF | Amlodipine 10mg/5ml oral solution sugar free |
| Antihypertensives | 0206020A0AAAUAU | BNF | Amlodipine 5mg/5ml oral suspension sugar free |
| Antihypertensives | 0206020A0AAAVAV | BNF | Amlodipine 25mg tablets |
| Antihypertensives | 0206020A0BBAAAA | BNF | Istin 5mg tablets |
| Antihypertensives | 0206020A0BBABAB | BNF | Istin 10mg tablets |
| Antihypertensives | 0206020A0BFAAAA | BNF | Amlostin 5mg tablets |
| Antihypertensives | 0206020A0BFABAB | BNF | Amlostin 10mg tablets |
| Antihypertensives | 0206020B0AAAAAA | BNF | Trimetazidine 20mg tablets |
| Antihypertensives | 0206020B0AAABAB | BNF | Trimetazidine 35mg modified-release tablets |
| Antihypertensives | 0206020B0BBAAAA | BNF | Vastarel 20mg tablets |
| Antihypertensives | 0206020B0BBABAB | BNF | Vastarel MR 35mg tablets |
| Antihypertensives | 0206020C0AAAAAA | BNF | Diltiazem 60mg modified-release tablets |
| Antihypertensives | 0206020C0AAACAC | BNF | Diltiazem 90mg modified-release tablets |
| Antihypertensives | 0206020C0AAAEAE | BNF | Diltiazem 300mg modified-release capsules |
| Antihypertensives | 0206020C0AAAJAJ | BNF | Diltiazem 60mg modified-release capsules |
| Antihypertensives | 0206020C0AAARAR | BNF | Diltiazem 60mg/5ml oral solution |
| Antihypertensives | 0206020C0AAASAS | BNF | Diltiazem 120mg modified-release tablets |
| Antihypertensives | 0206020C0AAATAT | BNF | Diltiazem 90mg modified-release capsules |
| Antihypertensives | 0206020C0AAAUAU | BNF | Diltiazem 120mg modified-release capsules |
| Antihypertensives | 0206020C0AAAVAV | BNF | Diltiazem 180mg modified-release capsules |
| Antihypertensives | 0206020C0AAAWAW | BNF | Diltiazem 240mg modified-release capsules |
| Antihypertensives | 0206020C0AAAXAX | BNF | Diltiazem 200mg modified-release capsules |
| Antihypertensives | 0206020C0AABABA | BNF | Diltiazem 360mg modified-release capsules |
| Antihypertensives | 0206020C0AABHBH | BNF | Diltiazem 10mg/5ml oral solution |
| Antihypertensives | 0206020C0AABIBI | BNF | Diltiazem 60mg/5ml oral suspension |
| Antihypertensives | 0206020C0BBAAAA | BNF | Tildiem 60mg modified-release tablets |
| Antihypertensives | 0206020C0BBABAC | BNF | Tildiem Retard 90mg tablets |
| Antihypertensives | 0206020C0BBACAS | BNF | Tildiem Retard 120mg tablets |
| Antihypertensives | 0206020C0BBADAE | BNF | Tildiem LA 300 capsules |
| Antihypertensives | 0206020C0BBAEAX | BNF | Tildiem LA 200 capsules |
| Antihypertensives | 0206020C0BCACAC | BNF | Calcicard CR 90mg tablets |
| Antihypertensives | 0206020C0BFAAAS | BNF | Adizem-SR 120mg tablets |
| Antihypertensives | 0206020C0BFACAT | BNF | Adizem-SR 90mg capsules |
| Antihypertensives | 0206020C0BFADAU | BNF | Adizem-SR 120mg capsules |
| Antihypertensives | 0206020C0BFAEAV | BNF | Adizem-SR 180mg capsules |
| Antihypertensives | 0206020C0BFAFAE | BNF | Adizem-XL 300mg capsules |
| Antihypertensives | 0206020C0BFAGAU | BNF | Adizem-XL 120mg capsules |
| Antihypertensives | 0206020C0BFAHAV | BNF | Adizem-XL 180mg capsules |
| Antihypertensives | 0206020C0BFAIAW | BNF | Adizem-XL 240mg capsules |
| Antihypertensives | 0206020C0BFAJAX | BNF | Adizem-XL 200mg capsules |
| Antihypertensives | 0206020C0BHAAAJ | BNF | Dilzem SR 60 capsules |
| Antihypertensives | 0206020C0BHABAT | BNF | Dilzem SR 90 capsules |
| Antihypertensives | 0206020C0BHACAU | BNF | Dilzem SR 120 capsules |
| Antihypertensives | 0206020C0BHADAU | BNF | Dilzem XL 120 capsules |
| Antihypertensives | 0206020C0BHAEAV | BNF | Dilzem XL 180 capsules |
| Antihypertensives | 0206020C0BHAFAW | BNF | Dilzem XL 240 capsules |
| Antihypertensives | 0206020C0BIAAAU | BNF | Slozem 120mg capsules |
| Antihypertensives | 0206020C0BIABAV | BNF | Slozem 180mg capsules |
| Antihypertensives | 0206020C0BIACAW | BNF | Slozem 240mg capsules |
| Antihypertensives | 0206020C0BIADAE | BNF | Slozem 300mg capsules |
| Antihypertensives | 0206020C0BJAAAT | BNF | Angitil SR 90 capsules |
| Antihypertensives | 0206020C0BJABAU | BNF | Angitil SR 120 capsules |
| Antihypertensives | 0206020C0BJACAV | BNF | Angitil SR 180 capsules |
| Antihypertensives | 0206020C0BJADAW | BNF | Angitil XL 240 capsules |
| Antihypertensives | 0206020C0BJAEAE | BNF | Angitil XL 300 capsules |
| Antihypertensives | 0206020C0BMAAAE | BNF | Zemtard 300 XL capsules |
| Antihypertensives | 0206020C0BMABAU | BNF | Zemtard 120 XL capsules |
| Antihypertensives | 0206020C0BMACAV | BNF | Zemtard 180 XL capsules |
| Antihypertensives | 0206020C0BMADAW | BNF | Zemtard 240 XL capsules |
| Antihypertensives | 0206020C0BNAAAU | BNF | Viazem XL 120mg capsules |
| Antihypertensives | 0206020C0BNABAV | BNF | Viazem XL 180mg capsules |
| Antihypertensives | 0206020C0BNACAW | BNF | Viazem XL 240mg capsules |
| Antihypertensives | 0206020C0BNADAE | BNF | Viazem XL 300mg capsules |
| Antihypertensives | 0206020C0BNAEBA | BNF | Viazem XL 360mg capsules |
| Antihypertensives | 0206020C0BPAAAT | BNF | Dilcardia SR 90mg capsules |
| Antihypertensives | 0206020C0BPABAU | BNF | Dilcardia SR 120mg capsules |
| Antihypertensives | 0206020C0BPACAJ | BNF | Dilcardia SR 60mg capsules |
| Antihypertensives | 0206020C0BRAAAJ | BNF | Bi-Carzem SR 60mg capsules |
| Antihypertensives | 0206020C0BRABAT | BNF | Bi-Carzem SR 90mg capsules |
| Antihypertensives | 0206020C0BRACAU | BNF | Bi-Carzem SR 120mg capsules |
| Antihypertensives | 0206020C0BRAEAE | BNF | Bi-Carzem XL 300mg capsules |
| Antihypertensives | 0206020C0BTAAAA | BNF | Retalzem 60 modified-release tablets |
| Antihypertensives | 0206020C0BVAAAV | BNF | Zemret 180 XL capsules |
| Antihypertensives | 0206020C0BVABAW | BNF | Zemret 240 XL capsules |
| Antihypertensives | 0206020C0BVACAE | BNF | Zemret 300 XL capsules |
| Antihypertensives | 0206020C0BWAEAJ | BNF | Disogram SR 60mg capsules |
| Antihypertensives | 0206020C0BYAAAJ | BNF | Kenzem SR 60mg capsules |
| Antihypertensives | 0206020C0BYABAT | BNF | Kenzem SR 90mg capsules |
| Antihypertensives | 0206020C0BYACAU | BNF | Kenzem SR 120mg capsules |
| Antihypertensives | 0206020C0BZABAU | BNF | Uard 120XL capsules |
| Antihypertensives | 0206020C0BZACAV | BNF | Uard 180XL capsules |
| Antihypertensives | 0206020F0AAABAB | BNF | Felodipine 5mg modified-release tablets |
| Antihypertensives | 0206020F0AAACAC | BNF | Felodipine 10mg modified-release tablets |
| Antihypertensives | 0206020F0AAADAD | BNF | Felodipine 25mg modified-release tablets |
| Antihypertensives | 0206020F0AAAEAE | BNF | Felodipine 25mg modified-release tablets (old) |
| Antihypertensives | 0206020F0AAAGAG | BNF | Felodipine 25mg/5ml oral solution |
| Antihypertensives | 0206020F0AAAHAH | BNF | Felodipine 5mg/5ml oral solution |
| Antihypertensives | 0206020F0BBAAAB | BNF | Plendil 5mg modified-release tablets |
| Antihypertensives | 0206020F0BBABAC | BNF | Plendil 10mg modified-release tablets |
| Antihypertensives | 0206020F0BBACAD | BNF | Plendil 25mg modified-release tablets |
| Antihypertensives | 0206020F0BDABAD | BNF | Cabren 25mg modified-release tablets |
| Antihypertensives | 0206020F0BDACAC | BNF | Cabren 10mg modified-release tablets |
| Antihypertensives | 0206020F0BEAAAB | BNF | Felotens XL 5mg tablets |
| Antihypertensives | 0206020F0BEABAC | BNF | Felotens XL 10mg tablets |
| Antihypertensives | 0206020F0BEACAD | BNF | Felotens XL 25mg tablets |
| Antihypertensives | 0206020F0BFAAAB | BNF | Felogen XL 5mg tablets |
| Antihypertensives | 0206020F0BFABAC | BNF | Felogen XL 10mg tablets |
| Antihypertensives | 0206020F0BGAAAB | BNF | Folpik XL 5mg tablets |
| Antihypertensives | 0206020F0BGABAC | BNF | Folpik XL 10mg tablets |
| Antihypertensives | 0206020F0BGACAD | BNF | Folpik XL 25mg tablets |
| Antihypertensives | 0206020F0BIAAAB | BNF | Vascalpha 5mg modified-release tablets (Accord) |
| Antihypertensives | 0206020F0BIABAC | BNF | Vascalpha 10mg modified-release tablets (Accord) |
| Antihypertensives | 0206020F0BIACAB | BNF | Vascalpha 5mg modified-release tablets (Almus) |
| Antihypertensives | 0206020F0BIADAC | BNF | Vascalpha 10mg modified-release tablets (Almus) |
| Antihypertensives | 0206020F0BIAEAB | BNF | Vascalpha 5mg modified-release tablets (NorthStar) |
| Antihypertensives | 0206020F0BIAFAC | BNF | Vascalpha 10mg modified-release tablets (NorthStar) |
| Antihypertensives | 0206020F0BJAAAB | BNF | Parmid XL 5mg tablets |
| Antihypertensives | 0206020F0BJABAC | BNF | Parmid XL 10mg tablets |
| Antihypertensives | 0206020F0BJACAD | BNF | Parmid XL 25mg tablets |
| Antihypertensives | 0206020F0BKAAAB | BNF | Cardioplen XL 5mg tablets |
| Antihypertensives | 0206020F0BKABAC | BNF | Cardioplen XL 10mg tablets |
| Antihypertensives | 0206020F0BKACAD | BNF | Cardioplen XL 25mg tablets |
| Antihypertensives | 0206020F0BLAAAB | BNF | Neofel XL 5mg tablets |
| Antihypertensives | 0206020F0BLABAC | BNF | Neofel XL 10mg tablets |
| Antihypertensives | 0206020F0BLACAD | BNF | Neofel XL 25mg tablets (Kent Pharm) |
| Antihypertensives | 0206020F0BLADAD | BNF | Neofel XL 25mg tablets (Actavis) |
| Antihypertensives | 0206020F0BLAEAD | BNF | Neofel XL 25mg tablets (Almus) |
| Antihypertensives | 0206020F0BMAAAC | BNF | Pinefeld XL 10mg tablets |
| Antihypertensives | 0206020I0AAAAAA | BNF | Isradipine 25mg tablets |
| Antihypertensives | 0206020K0AAAAAA | BNF | Lacidipine 2mg tablets |
| Antihypertensives | 0206020K0AAABAB | BNF | Lacidipine 4mg tablets |
| Antihypertensives | 0206020K0AAAEAE | BNF | Lacidipine 6mg tablets |
| Antihypertensives | 0206020K0BBAAAA | BNF | Motens 2mg tablets |
| Antihypertensives | 0206020K0BBABAB | BNF | Motens 4mg tablets |
| Antihypertensives | 0206020K0BDAAAB | BNF | Molap 4mg tablets |
| Antihypertensives | 0206020L0AAAAAA | BNF | Lercanidipine 10mg tablets |
| Antihypertensives | 0206020L0AAABAB | BNF | Lercanidipine 20mg tablets |
| Antihypertensives | 0206020L0BBAAAA | BNF | Zanidip 10mg tablets |
| Antihypertensives | 0206020L0BBABAB | BNF | Zanidip 20mg tablets |
| Antihypertensives | 0206020M0AAABAB | BNF | Nimodipine 30mg tablets |
| Antihypertensives | 0206020M0BBABAB | BNF | Nimotop 30mg tablets |
| Antihypertensives | 0206020Q0AAAAAA | BNF | Nicardipine 20mg capsules |
| Antihypertensives | 0206020Q0AAABAB | BNF | Nicardipine 30mg capsules |
| Antihypertensives | 0206020Q0AAACAC | BNF | Nicardipine 30mg modified-release capsules |
| Antihypertensives | 0206020Q0AAADAD | BNF | Nicardipine 45mg modified-release capsules |
| Antihypertensives | 0206020Q0BBAAAA | BNF | Cardene 20mg capsules |
| Antihypertensives | 0206020Q0BBABAB | BNF | Cardene 30mg capsules |
| Antihypertensives | 0206020Q0BBACAC | BNF | Cardene SR 30mg capsules |
| Antihypertensives | 0206020Q0BBADAD | BNF | Cardene SR 45mg capsules |
| Antihypertensives | 0206020R0AAAAAA | BNF | Nifedipine 5mg capsules |
| Antihypertensives | 0206020R0AAABAB | BNF | Nifedipine 10mg capsules |
| Antihypertensives | 0206020R0AAAEAE | BNF | Nifedipine 10mg modified-release tablets |
| Antihypertensives | 0206020R0AAAHAH | BNF | Nifedipine 20mg modified-release capsules |
| Antihypertensives | 0206020R0AAAMAM | BNF | Nifedipine 10mg modified-release capsules |
| Antihypertensives | 0206020R0AAANAN | BNF | Nifedipine 30mg modified-release tablets |
| Antihypertensives | 0206020R0AAAPAP | BNF | Nifedipine 60mg modified-release tablets |
| Antihypertensives | 0206020R0AAARAR | BNF | Nifedipine 20mg modified-release tablets |
| Antihypertensives | 0206020R0AAASAS | BNF | Nifedipine 20mg/ml oral drops |
| Antihypertensives | 0206020R0AAAUAU | BNF | Nifedipine 100mg/5ml oral suspension |
| Antihypertensives | 0206020R0AAAXAX | BNF | Nifedipine 40mg modified-release tablets |
| Antihypertensives | 0206020R0AABCBC | BNF | Nifedipine 25mg/5ml oral suspension |
| Antihypertensives | 0206020R0AABEBE | BNF | Nifedipine 30mg modified-release capsules |
| Antihypertensives | 0206020R0AABFBF | BNF | Nifedipine 60mg modified-release capsules |
| Antihypertensives | 0206020R0AABKBK | BNF | Nifedipine 25mg/5ml oral suspension |
| Antihypertensives | 0206020R0AABQBQ | BNF | Nifedipine 10mg/5ml oral suspension |
| Antihypertensives | 0206020R0AABRBR | BNF | Nifedipine 5mg/5ml oral suspension |
| Antihypertensives | 0206020R0BBAAAA | BNF | Adalat 5mg capsules |
| Antihypertensives | 0206020R0BBABAB | BNF | Adalat 10mg capsules |
| Antihypertensives | 0206020R0BBAFAR | BNF | Adalat retard 20mg tablets |
| Antihypertensives | 0206020R0BBAGAE | BNF | Adalat retard 10mg tablets |
| Antihypertensives | 0206020R0BBAHAN | BNF | Adalat LA 30mg tablets |
| Antihypertensives | 0206020R0BBAIAP | BNF | Adalat LA 60mg tablets |
| Antihypertensives | 0206020R0BBAJAR | BNF | Adalat LA 20mg tablets |
| Antihypertensives | 0206020R0BGAAAH | BNF | Coracten SR 20mg capsules |
| Antihypertensives | 0206020R0BGABAM | BNF | Coracten SR 10mg capsules |
| Antihypertensives | 0206020R0BGACBE | BNF | Coracten XL 30mg capsules |
| Antihypertensives | 0206020R0BGADBF | BNF | Coracten XL 60mg capsules |
| Antihypertensives | 0206020R0BHAEAE | BNF | Angiopine MR 10mg tablets |
| Antihypertensives | 0206020R0BKADAE | BNF | Kentipine MR 10 tablets |
| Antihypertensives | 0206020R0BLAAAR | BNF | Cardilate MR 20mg tablets |
| Antihypertensives | 0206020R0BLABAE | BNF | Cardilate MR 10mg tablets |
| Antihypertensives | 0206020R0BRAAAR | BNF | Adipine MR 20 tablets |
| Antihypertensives | 0206020R0BRABAE | BNF | Adipine MR 10 tablets |
| Antihypertensives | 0206020R0BRACAN | BNF | Adipine XL 30mg tablets |
| Antihypertensives | 0206020R0BRADAP | BNF | Adipine XL 60mg tablets |
| Antihypertensives | 0206020R0BTAAAS | BNF | Nifedipin-ratiopharm 20mg/ml oral drops |
| Antihypertensives | 0206020R0BWAAAE | BNF | Tensipine MR 10 tablets |
| Antihypertensives | 0206020R0BWABAR | BNF | Tensipine MR 20 tablets |
| Antihypertensives | 0206020R0BXAAAX | BNF | Fortipine LA 40 tablets |
| Antihypertensives | 0206020R0CBAAAE | BNF | Nifedipress MR 10 tablets (Dexcel-Pharma) |
| Antihypertensives | 0206020R0CBABAR | BNF | Nifedipress MR 20 tablets (Dexcel-Pharma) |
| Antihypertensives | 0206020R0CBACAE | BNF | Nifedipress MR 10 tablets (Teva) |
| Antihypertensives | 0206020R0CBADAR | BNF | Nifedipress MR 20 tablets (Actavis) |
| Antihypertensives | 0206020R0CBAEAR | BNF | Nifedipress MR 20 tablets (Teva) |
| Antihypertensives | 0206020R0CEAAAR | BNF | Nifopress Retard 20mg tablets |
| Antihypertensives | 0206020R0CEABAR | BNF | Nifopress MR 20mg tablets |
| Antihypertensives | 0206020R0CFAAAR | BNF | Valni 20 Retard tablets |
| Antihypertensives | 0206020R0CFABAN | BNF | Valni XL 30mg tablets |
| Antihypertensives | 0206020R0CFACAP | BNF | Valni XL 60mg tablets |
| Antihypertensives | 0206020R0CGAAAE | BNF | Calchan MR 10 tablets |
| Antihypertensives | 0206020R0CGABAR | BNF | Calchan MR 20 tablets |
| Antihypertensives | 0206020R0CHAAAN | BNF | Neozipine XL 30mg tablets |
| Antihypertensives | 0206020R0CHABAP | BNF | Neozipine XL 60mg tablets |
| Antihypertensives | 0206020R0CIAAAN | BNF | Adanif XL 30mg tablets |
| Antihypertensives | 0206020R0CIABAP | BNF | Adanif XL 60mg tablets |
| Antihypertensives | 0206020R0CJAAAN | BNF | Nidef 30mg modified-release tablets |
| Antihypertensives | 0206020R0CJABAP | BNF | Nidef 60mg modified-release tablets |
| Antihypertensives | 0206020T0AAAAAA | BNF | Verapamil 5mg/2ml solution for injection ampoules |
| Antihypertensives | 0206020T0AAACAC | BNF | Verapamil 40mg tablets |
| Antihypertensives | 0206020T0AAADAD | BNF | Verapamil 80mg tablets |
| Antihypertensives | 0206020T0AAAFAF | BNF | Verapamil 120mg tablets |
| Antihypertensives | 0206020T0AAAGAG | BNF | Verapamil 160mg tablets |
| Antihypertensives | 0206020T0AAAHAH | BNF | Verapamil 240mg modified-release tablets |
| Antihypertensives | 0206020T0AAAIAI | BNF | Verapamil 120mg modified-release capsules |
| Antihypertensives | 0206020T0AAAJAJ | BNF | Verapamil 180mg modified-release capsules |
| Antihypertensives | 0206020T0AAAKAK | BNF | Verapamil 240mg modified-release capsules |
| Antihypertensives | 0206020T0AAAUAU | BNF | Verapamil 120mg modified-release tablets |
| Antihypertensives | 0206020T0AAAVAV | BNF | Verapamil 40mg/5ml oral solution sugar free |
| Antihypertensives | 0206020T0AAAZAZ | BNF | Verapamil 40mg/5ml oral liquid |
| Antihypertensives | 0206020T0AABIBI | BNF | Verapamil 120mg modified-release tablets (old) |
| Antihypertensives | 0206020T0BCAFAH | BNF | Cordilox MR 240mg tablets |
| Antihypertensives | 0206020T0BDACAF | BNF | Securon 120mg tablets |
| Antihypertensives | 0206020T0BDAEAH | BNF | Securon SR 240mg tablets |
| Antihypertensives | 0206020T0BDAGAU | BNF | Half Securon SR 120mg tablets |
| Antihypertensives | 0206020T0BDAIAA | BNF | Securon IV 5mg/2ml solution for injection ampoules |
| Antihypertensives | 0206020T0BFAAAI | BNF | Univer 120mg modified-release capsules |
| Antihypertensives | 0206020T0BFABAJ | BNF | Univer 180mg modified-release capsules |
| Antihypertensives | 0206020T0BFACAK | BNF | Univer 240mg modified-release capsules |
| Antihypertensives | 0206020T0BHAAAH | BNF | Verapress MR 240mg tablets (Dexcel-Pharma) |
| Antihypertensives | 0206020T0BHABAH | BNF | Verapress MR 240mg tablets (Sandoz) |
| Antihypertensives | 0206020T0BHACAH | BNF | Verapress MR 240mg tablets (Actavis) |
| Antihypertensives | 0206020T0BHADAH | BNF | Verapress MR 240mg tablets (Teva) |
| Antihypertensives | 0206020T0BJAAAH | BNF | Vertab SR 240 tablets |
| Antihypertensives | 0206020T0BKAAAV | BNF | Zolvera 40mg/5ml oral solution |
| Antihypertensives | 0206020T0BMAAAU | BNF | Vera-Til SR 120mg tablets (Tillomed) |
| Antihypertensives | 0206020T0BMABAH | BNF | Vera-Til SR 240mg tablets (Tillomed) |
| Antihypertensives | 0206020T0BMACAH | BNF | Vera-Til SR 240mg tablets (Accord Healthcare Ltd) |
| Antihypertensives | 0206020T0BMADAU | BNF | Vera-Til SR 120mg tablets (Accord Healthcare Ltd) |
| Antihypertensives | 0206020Z0AAAAAA | BNF | Amlodipine 5mg / Valsartan 80mg tablets |
| Antihypertensives | 0206020Z0AAABAB | BNF | Amlodipine 5mg / Valsartan 160mg tablets |
| Antihypertensives | 0206020Z0AAACAC | BNF | Amlodipine 10mg / Valsartan 160mg tablets |
| Antihypertensives | 0206020Z0BBAAAA | BNF | Exforge 5mg/80mg tablets |
| Antihypertensives | 0206020Z0BBABAB | BNF | Exforge 5mg/160mg tablets |
| Antihypertensives | 0206020Z0BBACAC | BNF | Exforge 10mg/160mg tablets |
| Statins | 0212000ABBJAAAA | BNF | AceOmeg 1000mg capsules |
| Statins | 0212000A0AAAAAA | BNF | Acipimox 250mg capsules |
| Statins | 0212000AIAAABAB | BNF | Alirocumab 150mg/1ml inj pre-filled disposable devices |
| Statins | 0212000AIAAAAAA | BNF | Alirocumab 75mg/1ml inj pre-filled disposable devices |
| Statins | 0212000B0AAALAL | BNF | Atorvastatin 10mg chewable tablets sugar free |
| Statins | 0212000B0AAAAAA | BNF | Atorvastatin 10mg tablets |
| Statins | 0212000B0AAAHAH | BNF | Atorvastatin 10mg/5ml oral liquid |
| Statins | 0212000B0AAAMAM | BNF | Atorvastatin 20mg chewable tablets sugar free |
| Statins | 0212000B0AAABAB | BNF | Atorvastatin 20mg tablets |
| Statins | 0212000B0AAAFAF | BNF | Atorvastatin 20mg/5ml oral solution |
| Statins | 0212000B0AAAQAQ | BNF | Atorvastatin 20mg/5ml oral suspension |
| Statins | 0212000B0AAANAN | BNF | Atorvastatin 30mg tablets |
| Statins | 0212000B0AAACAC | BNF | Atorvastatin 40mg tablets |
| Statins | 0212000B0AAAGAG | BNF | Atorvastatin 40mg/5ml oral liquid |
| Statins | 0212000B0AAAPAP | BNF | Atorvastatin 60mg tablets |
| Statins | 0212000B0AAADAD | BNF | Atorvastatin 80mg tablets |
| Statins | 0212000B0AAAIAI | BNF | Atorvastatin 80mg/5ml oral suspension |
| Statins | 0212000ALAAAAAA | BNF | Bempedoic acid 180mg / Ezetimibe 10mg tablets |
| Statins | 0212000AKAAAAAA | BNF | Bempedoic acid 180mg tablets |
| Statins | 0212000D0AAAAAA | BNF | Bezafibrate 200mg tablets |
| Statins | 0212000D0AAACAC | BNF | Bezafibrate 200mg/5ml oral suspension |
| Statins | 0212000D0AAABAB | BNF | Bezafibrate 400mg modified-release tablets |
| Statins | 0212000D0BBAAAA | BNF | Bezalip 200mg tablets |
| Statins | 0212000D0BBABAB | BNF | Bezalip Mono 400mg modified-release tablets |
| Statins | 0212000ADBBAAAA | BNF | Cholestagel 625mg tablets |
| Statins | 0212000AJBBAAAA | BNF | Cholib 145mg/20mg tablets |
| Statins | 021200010AAAAAA | BNF | Ciprofibrate 100mg tablets |
| Statins | 0212000ADAAAAAA | BNF | Colesevelam 625mg tablets |
| Statins | 0212000K0BBACAC | BNF | Colestid 1g tablets |
| Statins | 0212000K0BBAAAA | BNF | Colestid 5g granules sachets plain |
| Statins | 0212000K0BBABAA | BNF | Colestid Orange 5g granules sachets |
| Statins | 0212000K0AAACAC | BNF | Colestipol 1g tablets |
| Statins | 0212000K0AAAAAA | BNF | Colestipol 5g granules sachets sugar free |
| Statins | 0212000F0AAABAB | BNF | Colestyramine 4g oral powder sachets |
| Statins | 0212000F0AAAEAE | BNF | Colestyramine 4g oral powder sachets sugar free |
| Statins | 0212000AABBAAAA | BNF | Crestor 10mg tablets |
| Statins | 0212000AABBABAB | BNF | Crestor 20mg tablets |
| Statins | 0212000AABBACAC | BNF | Crestor 40mg tablets |
| Statins | 0212000AABBADAD | BNF | Crestor 5mg tablets |
| Statins | 0212000M0BEAAAD | BNF | Dorisin XL 80mg tablets |
| Statins | 0212000ABBHAAAA | BNF | Dualtis 1000mg capsules |
| Statins | 0212000V0AAABAB | BNF | Eicosapentaenoic acid 170mg/Docosahexaenoic acid 115mg caps |
| Statins | 0212000ABAAAAAA | BNF | Eicosapentaenoic acid 460mg/Docosahexaenoic acid 380mg caps |
| Statins | 0212000AHAAAAAA | BNF | Evolocumab 140mg/1ml inj pre-filled disposable devices |
| Statins | 0212000AHAAABAB | BNF | Evolocumab 140mg/1ml inj pre-filled syringes |
| Statins | 0212000L0AAAAAA | BNF | Ezetimibe 10mg tablets |
| Statins | 0212000L0BBAAAA | BNF | Ezetrol 10mg tablets |
| Statins | 0212000AJAAAAAA | BNF | Fenofibrate 145mg / Simvastatin 20mg tablets |
| Statins | 0212000AJAAABAB | BNF | Fenofibrate 145mg / Simvastatin 40mg tablets |
| Statins | 0212000P0AAAFAF | BNF | Fenofibrate 200mg capsules |
| Statins | 0212000P0AAAEAE | BNF | Fenofibrate micronised 160mg tablets |
| Statins | 0212000P0AAABAB | BNF | Fenofibrate micronised 200mg capsules |
| Statins | 0212000P0AAADAD | BNF | Fenofibrate micronised 267mg capsules |
| Statins | 0212000P0AAACAC | BNF | Fenofibrate micronised 67mg capsules |
| Statins | 0212000P0BDAAAF | BNF | Fenogal 200mg capsules |
| Statins | 0212000D0BFAAAB | BNF | Fibrazate XL 400mg tablets |
| Statins | 0212000M0AAAAAA | BNF | Fluvastatin 20mg capsules |
| Statins | 0212000M0AAABAB | BNF | Fluvastatin 40mg capsules |
| Statins | 0212000M0AAADAD | BNF | Fluvastatin 80mg modified-release tablets |
| Statins | 0212000ABBIAAAA | BNF | G & G Omega 3 1000mg softgels capsules |
| Statins | 0212000Q0AAAAAA | BNF | Gemfibrozil 300mg capsules |
| Statins | 0212000Q0AAABAB | BNF | Gemfibrozil 600mg tablets |
| Statins | 0212000ACBBAAAA | BNF | Inegy 10mg/20mg tablets |
| Statins | 0212000ACBBABAB | BNF | Inegy 10mg/40mg tablets |
| Statins | 0212000ACBBACAC | BNF | Inegy 10mg/80mg tablets |
| Statins | 0212000M0BBAAAA | BNF | Lescol 20mg capsules |
| Statins | 0212000M0BBABAB | BNF | Lescol 40mg capsules |
| Statins | 0212000M0BBACAD | BNF | Lescol XL 80mg tablets |
| Statins | 0212000P0BBABAB | BNF | Lipantil Micro 200 capsules |
| Statins | 0212000P0BBADAD | BNF | Lipantil Micro 267 capsules |
| Statins | 0212000P0BBACAC | BNF | Lipantil Micro 67 capsules |
| Statins | 0212000B0BBAEAL | BNF | Lipitor 10mg chewable tablets |
| Statins | 0212000B0BBAAAA | BNF | Lipitor 10mg tablets |
| Statins | 0212000B0BBAFAM | BNF | Lipitor 20mg chewable tablets |
| Statins | 0212000B0BBABAB | BNF | Lipitor 20mg tablets |
| Statins | 0212000B0BBACAC | BNF | Lipitor 40mg tablets |
| Statins | 0212000B0BBADAD | BNF | Lipitor 80mg tablets |
| Statins | 0212000X0BBABAB | BNF | Lipostat 20mg tablets |
| Statins | 0212000X0BBACAD | BNF | Lipostat 40mg tablets |
| Statins | 0212000Q0BBAAAA | BNF | Lopid 300mg capsules |
| Statins | 0212000Q0BBABAB | BNF | Lopid 600mg tablets |
| Statins | 0212000M0BCAAAD | BNF | Luvinsta XL 80mg tablets |
| Statins | 0212000V0BBABAB | BNF | MaxEPA 1g capsules |
| Statins | 021200010BBAAAA | BNF | Modalim 100mg tablets |
| Statins | 0212000M0BGAAAD | BNF | Nandovar XL 80mg tablets |
| Statins | 0212000ABBFAAAA | BNF | Nebbaro 1000mg capsules |
| Statins | 0212000U0AAAVAV | BNF | Nicotinic acid 500mg modified-release tablets |
| Statins | 0212000U0AAABAB | BNF | Nicotinic acid 50mg tablets |
| Statins | 0212000AKBBAAAA | BNF | Nilemdo 180mg tablets |
| Statins | 0212000ALBBAAAA | BNF | Nustendi 180mg/10mg tablets |
| Statins | 0212000A0BBAAAA | BNF | Olbetam 250mg capsules |
| Statins | 0212000ABBBAAAA | BNF | Omacor capsules |
| Statins | 0212000ABBGAAAA | BNF | Omega 3 1000mg capsules |
| Statins | 0212000ABBEAAAA | BNF | Omega 3-acid-ethyl esters 1000mg capsules |
| Statins | 0212000AGAAAAAA | BNF | Policosanol 10mg capsules |
| Statins | 0212000AIBBABAB | BNF | Praluent 150mg/1ml solution for injection pre-filled pens |
| Statins | 0212000AIBBAAAA | BNF | Praluent 75mg/1ml solution for injection pre-filled pens |
| Statins | 0212000X0AAAAAA | BNF | Pravastatin 10mg tablets |
| Statins | 0212000X0AAABAB | BNF | Pravastatin 20mg tablets |
| Statins | 0212000X0AAADAD | BNF | Pravastatin 40mg tablets |
| Statins | 0212000X0AAAHAH | BNF | Pravastatin 40mg/5ml oral liquid |
| Statins | 0212000X0AAAKAK | BNF | Pravastatin 5mg/5ml oral liquid |
| Statins | 0212000ABBDAAAA | BNF | Prestylon 1g capsules |
| Statins | 0212000F0BBAAAB | BNF | Questran 4g oral powder sachets |
| Statins | 0212000F0BBACAE | BNF | Questran Light 4g oral powder sachets |
| Statins | 0212000AHBBABAB | BNF | Repatha 140mg/1ml solution for injection pre-filled syringes |
| Statins | 0212000AHBBAAAA | BNF | Repatha SureClick 140mg/1ml inj pre-filled pens |
| Statins | 0212000AAAAAAAA | BNF | Rosuvastatin 10mg tablets |
| Statins | 0212000AAAAABAB | BNF | Rosuvastatin 20mg tablets |
| Statins | 0212000AAAAACAC | BNF | Rosuvastatin 40mg tablets |
| Statins | 0212000AAAAADAD | BNF | Rosuvastatin 5mg tablets |
| Statins | 0212000Y0BDAAAA | BNF | Simvador 10mg tablets |
| Statins | 0212000Y0BDABAB | BNF | Simvador 20mg tablets |
| Statins | 0212000Y0BDACAD | BNF | Simvador 40mg tablets |
| Statins | 0212000Y0BDADAH | BNF | Simvador 80mg tablets |
| Statins | 0212000Y0AAAAAA | BNF | Simvastatin 10mg tablets |
| Statins | 0212000ACAAAAAA | BNF | Simvastatin 20mg / Ezetimibe 10mg tablets |
| Statins | 0212000Y0AAABAB | BNF | Simvastatin 20mg tablets |
| Statins | 0212000Y0AAAKAK | BNF | Simvastatin 20mg/5ml oral suspension sugar free |
| Statins | 0212000ACAAABAB | BNF | Simvastatin 40mg / Ezetimibe 10mg tablets |
| Statins | 0212000Y0AAADAD | BNF | Simvastatin 40mg tablets |
| Statins | 0212000Y0AAALAL | BNF | Simvastatin 40mg/5ml oral suspension sugar free |
| Statins | 0212000ACAAACAC | BNF | Simvastatin 80mg / Ezetimibe 10mg tablets |
| Statins | 0212000Y0AAAHAH | BNF | Simvastatin 80mg tablets |
| Statins | 0212000P0BCAAAE | BNF | Supralip 160mg tablets |
| Statins | 0212000ABBCAAAA | BNF | Teromeg 1000mg capsules |
| Statins | 0212000Y0BBAAAA | BNF | Zocor 10mg tablets |
| Statins | 0212000Y0BBABAB | BNF | Zocor 20mg tablets |
| Statins | 0212000Y0BBACAD | BNF | Zocor 40mg tablets |
| Statins | 0212000Y0BBADAH | BNF | Zocor 80mg tablets |
| Statins | 0212000Y0BBAEAA | BNF | Zocor Heart-Pro 10mg tablets |

**Table S4. Number of individuals excluded during sample selection for prior morbidities. Note that morbidities are not mutually exclusive**

| **Excluded condition** | **N** |
| --- | --- |
| Stroke | 66,288 |
| Dementia | 46,816 |
| Parkinson’s disease | 6,274 |
| Epilepsy | 134,028 |
| Colorectal cancer | 8,946 |
| Transient Ischaemic Attack | 26,182 |
| External Haematoma | 69 |
| Intracranial Haemorrhage | 2,302 |
| Subarachnoid Haemorrhage | 5,131 |
| Subdural Haemorrhage | 3,547 |

**Table S5. Prevalence of each brain imaging phenotype by age band**

| **Age band (years)** | **WMH**  **(N = 82,284)** | **Lacunes**  **(N = 28,477)** | **Cortical Infarcts**  **(N = 8,221)** | **Atrophy**  **(N = 80,772)** | **None**  **(N = 238,789)** |
| --- | --- | --- | --- | --- | --- |
| <=30 | 201 (0.2%) | 74 (0.3%) | 22 (0.3%) | 410 (0.5%) | 45,733 (19.2%) |
| 31-50 | 2,186 (2.7%) | 1,045 (3.7%) | 314 (3.8%) | 3,475 (4.3%) | 85,846 (36.0%) |
| 51-70 | 21,015 (25.5%) | 7,639 (26.8%) | 2,134 (26.0%) | 19,772 (24.5%) | 81,234 (34.0%) |
| 71-90 | 54,034 (65.7%) | 18,082 (63.5%) | 5,305 (64.5%) | 52,495 (65.0%) | 25,208 (10.5%) |
| >=91 | 4,848 (5.9%) | 1,637 (5.7%) | 446 (5.4%) | 4,620 (5.7%) | 768 (0.3%) |

**Table S6. Risk table showing incidence of each outcome for the analytic cohort (N = 367,988)**

| **Outcome** |  | **0 Years** | **1 Year** | **2 Years** | **3 Years** | **4 Years** | **5 Years** | **6 Years** | **7 Years** | **8 Years** | **9 Years** | **10 Years** | **11 Years** | **12 Years** |
| --- | --- | --- | --- | --- | --- | --- | --- | --- | --- | --- | --- | --- | --- | --- |
| Stroke | At risk | 367,988 | 348,027 | 331,546 | 317,559 | 287,040 | 239,881 | 196,045 | 155,483 | 118,160 | 84,194 | 54,133 | 27,665 | 3,849 |
|  | Events | 11 | 2,979 | 5,546 | 7,584 | 9,183 | 10,467 | 11,402 | 12,068 | 12,524 | 12,802 | 12,924 | 12,951 | 12,951 |
| Dementia | At risk | 367,988 | 338,939 | 319,930 | 305,665 | 276,298 | 231,401 | 189,792 | 151,072 | 115,343 | 82,544 | 53,361 | 27,386 | 3,840 |
|  | Events | 61 | 13,333 | 20,861 | 26,061 | 29,707 | 32,226 | 33,910 | 35,114 | 35,802 | 36,245 | 36,405 | 36,430 | 36,430 |
| Parkinson’s Disease | At risk | 367,988 | 349,105 | 333,353 | 319,821 | 289,640 | 242,461 | 198,502 | 157,728 | 120,029 | 85,626 | 55,137 | 28,268 | 3,958 |
|  | Events | <10 | 868 | 1,694 | 2,429 | 2,939 | 3,299 | 3,575 | 3,745 | 3,859 | 3,917 | 3,943 | 3,949 | 3,949 |
| Epilepsy | At risk | 367,988 | 336,876 | 313,089 | 293,661 | 260,036 | 212,192 | 169,356 | 131,243 | 97,551 | 68,064 | 42,986 | 21,529 | 2,935 |
|  | Events | 59 | 13,977 | 23,790 | 31,403 | 37,136 | 41,432 | 44,590 | 46,740 | 48,073 | 48,852 | 49,183 | 49,227 | 49,227 |
| Colorectal cancer | At risk | 367,988 | 349,404 | 333,902 | 320,557 | 290,281 | 242,978 | 198,872 | 157,939 | 120,134 | 85,661 | 55,130 | 28,241 | 3,948 |
|  | Events | <10 | 595 | 1061 | 1,486 | 1,834 | 2,100 | 2,310 | 2,474 | 2,571 | 2,617 | 2,648 | 2,650 | 2,650 |

**Table S7. Risk table showing incidence of each outcome for the wider Scottish population (the analytic sample plus the unscanned population alive at 01/07/2010) (N = 4, 637,231)**

| **Outcome** |  | **0 Years** | **1 Year** | **2 Years** | **3 Years** | **4 Years** | **5 Years** | **6 Years** | **7 Years** | **8 Years** | **9 Years** | **10 Years** | **11 Years** | **12 Years** |
| --- | --- | --- | --- | --- | --- | --- | --- | --- | --- | --- | --- | --- | --- | --- |
| Stroke | At risk | 4,637,231 | 4,562,086 | 4,492,778 | 4,424,780 | 4,344,713 | 4,245,602 | 4,155,449 | 4,068,408 | 3,983,912 | 3,895,019 | 3,806,607 | 3,748,093 | 3,724,274 |
|  | Events | 16 | 5,364 | 9,682 | 13,238 | 15,872 | 18,064 | 19,713 | 21,038 | 22,072 | 28,034 | 35,034 | 38,865 | 38,868 |
| Dementia | At risk | 4,637,231 | 4,539,576 | 4,467,470 | 4,400,150 | 4,322,140 | 4,226,787 | 4,139,883 | 4,055,532 | 3,973,441 | 3,888,274 | 3,803,106 | 3,746,095 | 3,722,538 |
|  | Events | 84 | 23,468 | 38,473 | 49,437 | 57,108 | 63,274 | 67,849 | 71,749 | 74,769 | 79,511 | 85,751 | 89,110 | 89,121 |
| Parkinson’s Disease | At risk | 4,637,231 | 4,562,840 | 4,494,217 | 4,426,752 | 4,347,027 | 4,247,903 | 4,157,647 | 4,070,325 | 3,985,413 | 3,899,895 | 3,815,134 | 3,758,414 | 3,734,104 |
|  | Events | 10 | 2,312 | 4,203 | 5,676 | 6,774 | 7,641 | 8,367 | 8,976 | 9,488 | 10,150 | 10,908 | 11,342 | 11,342 |
| Epilepsy | At risk | 4,637,231 | 4,469,912 | 4,369,913 | 4,271,582 | 4,161,194 | 4,033,374 | 3,914,122 | 3,799,854 | 3,692,723 | 3,587,213 | 3,488,298 | 3,428,076 | 3,409,480 |
|  | Events | 66 | 37,413 | 75,098 | 112,225 | 149,647 | 186,701 | 224,809 | 261,611 | 294,350 | 326,369 | 354,149 | 367,583 | 367,585 |
| Colorectal cancer | At risk | 4,637,231 | 4,559,831 | 4,489,898 | 4,421,196 | 4,339,912 | 4,239,365 | 4,147,681 | 4,058,943 | 3,972,344 | 3,885,220 | 3,799,583 | 3,742,618 | 3,718,325 |
|  | Events | 15 | 4,616 | 8,576 | 12,168 | 15,604 | 18,724 | 21,776 | 24,773 | 28,023 | 31,535 | 34,683 | 36,177 | 36,177 |

**Table S8. Median (and Interquartile Range) follow-up time (in years) for each outcome in the analytic cohort and wider Scottish population (the analytic sample plus the unscanned population alive at 01/07/2010)**

| **Outcome** | **Analytic** | | | | | | **Population** |
| --- | --- | --- | --- | --- | --- | --- | --- |
|  | WMH | Lacunes | Cortical Infarcts | Cerebral Atrophy | None | Total | Total |
| Stroke | 4.7 (2.5, 7.0) | 4.6 (2.3, 7.0) | 4.3 (1.9, 6.8) | 4.7 (2.5, 7.1) | 7.0 (4.9, 9.3) | 6.3 (4.2, 8.8) | 12.2 (12.2, 12.2) |
| Ischaemic stroke | 4.7 (2.6, 7.0) | 4.7 (2.4, 7.1) | 4.4 (2.0, 6.9) | 4.8 (2.6, 7.1) | 7.0 (4.9, 9.4) | 6.3 (4.3, 8.8) | 12.2 (12.2, 12.2) |
| Intracerebral haemorrhage | 4.7 (2.6, 7.1) | 4.7 (2.5, 7.1) | 4.4 (2.0, 6.9) | 4.8 (2.6, 7.1) | 7.0 (4.9, 9.4) | 6.4 (4.3, 8.8) | 12.2 (12.2, 12.2) |
| Unspecified stroke | 4.8 (2.6, 7.1) | 4.8 (2.5, 7.2) | 4.5 (2.1, 7.0) | 4.8 (2.6, 7.2) | 7.0 (4.9, 9.4) | 6.4 (4.3, 8.8) | 12.2 (12.2, 12.2) |
| Dementia | 4.2 (1.6, 6.6) | 4.2 (1.7, 6.7) | 4.0 (1.4, 6.6) | 4.1 (1.5, 6.6) | 7.0 (4.9, 9.3) | 6.2 (4.0, 8.7) | 12.2 (12.2, 12.2) |
| Alzheimer’s dementia | 4.8 (2.6, 7.1) | 4.8 (2.6, 7.2) | 4.5 (2.1, 7.0) | 4.8 (2.6, 7.2) | 7.1 (4.9, 9.4) | 6.4 (4.4, 8.8) | 12.2 (12.2, 12.2) |
| Vascular dementia | 4.7 (2.4, 7.0) | 4.7 (2.3, 7.1) | 4.3 (1.8, 6.9) | 4.7 (2.4, 7.1) | 7.1 (4.9, 9.4) | 6.3 (4.3, 8.8) | 12.2 (12.2, 12.2) |
| Unspecified dementia | 4.7 (2.4, 7.0) | 4.7 (2.3, 7.1) | 4.4 (1.9, 6.9) | 4.7 (2.4, 7.1) | 7.0 (4.9, 9.4) | 6.3 (4.3, 8.8) | 12.2 (12.2, 12.2) |
| Parkinson’s Disease | 4.8 (2.6, 7.1) | 4.8 (2.5, 7.2) | 4.5 (2.1, 7.0) | 4.8 (2.6, 7.1) | 7.0 (4.9, 9.4) | 6.4 (4.3, 8.8) | 12.2 (12.2, 12.2) |
| Epilepsy | 4.4 (2.1, 6.6) | 4.4 (2.1, 6.7) | 4.1 (1.8, 6.6) | 4.4 (2.1, 6.7) | 6.3 (4.2, 8.7) | 5.7 (3.7, 8.2) | 12.2 (12.2, 12.2) |
| Colorectal cancer | 4.8 (2.7, 7.1) | 4.8 (2.6, 7.2) | 4.5 (2.1, 7.0) | 4.8 (2.7, 7.2) | 7.0 (4.9, 9.4) | 6.4 (4.3, 8.8) | 12.2 (12.2, 12.2) |

**Table S9. 1-year and 5-year unadjusted absolute risks (AR) for each scan phenotype and outcome, and the absolute risk increase (ARI) and relative risk ratios (RR) compared both to the analytic cohort without the phenotype and to the wider Scottish population**

|  | **WMH** | | | | | **Lacunes** | | | | | **Cortical infarcts** | | | | | **Cerebral atrophy** | | | | |
| --- | --- | --- | --- | --- | --- | --- | --- | --- | --- | --- | --- | --- | --- | --- | --- | --- | --- | --- | --- | --- |
|  |  | Analytic | | Population | |  | Analytic | | Population | |  | Analytic | | Population | |  | Analytic | | Population | |
|  | AR | ARI | RR | ARI | RR | AR | ARI | RR | ARI | RR | AR | ARI | RR | ARI | RR | AR | ARI | RR | ARI | RR |
| **1-Year Survival** |  |  |  |  |  |  |  |  |  |  |  |  |  |  |  |  |  |  |  |  |
| Stroke | 0.02 | 0.01 | 2.70 | 0.02 | 17.23 | 0.03 | 0.02 | 3.42 | 0.03 | 21.89 | 0.04 | 0.03 | 4.78 | 0.04 | 30.54 | 0.02 | 0.01 | 2.40 | 0.02 | 15.34 |
| Dementia | 0.12 | 0.08 | 2.97 | 0.11 | 15.13 | 0.10 | 0.06 | 2.56 | 0.09 | 13.05 | 0.11 | 0.07 | 2.88 | 0.11 | 14.67 | 0.14 | 0.10 | 3.47 | 0.13 | 17.67 |
| Parkinson’s | 0.01 | <0.01 | 2.47 | <0.01 | 8.71 | 0.01 | <0.01 | 2.42 | <0.01 | 8.56 | <0.01 | <0.01 | 1.48 | <0.01 | 5.23 | 0.01 | <0.01 | 2.63 | <0.01 | 9.30 |
| Epilepsy | 0.04 | -0.01 | 0.88 | 0.01 | 1.64 | 0.04 | >-0.01 | 0.89 | 0.01 | 1.64 | 0.04 | >-0.01 | 0.96 | 0.02 | 1.78 | 0.04 | -0.01 | 0.87 | 0.01 | 1.62 |
| Colorectal Cancer | <0.01 | <0.01 | 2.00 | <0.01 | 2.14 | <0.01 | <0.01 | 2.04 | <0.01 | 2.18 | <0.01 | <0.01 | 2.67 | <0.01 | 2.84 | <0.01 | <0.01 | 2.03 | <0.01 | 2.17 |
| **5-Year Survival** |  |  |  |  |  |  |  |  |  |  |  |  |  |  |  |  |  |  |  |  |
| Stroke | 0.15 | 0.10 | 3.34 | 0.14 | 101.39 | 0.19 | 0.15 | 4.33 | 0.19 | 131.24 | 0.26 | 0.22 | 5.93 | 0.26 | 179.91 | 0.13 | 0.09 | 2.99 | 0.13 | 90.58 |
| Dementia | 0.58 | 0.44 | 4.14 | 0.57 | 69.63 | 0.52 | 0.38 | 3.72 | 0.51 | 62.43 | 0.57 | 0.43 | 4.10 | 0.56 | 68.89 | 0.64 | 0.50 | 4.59 | 0.63 | 77.18 |
| Parkinson’s | 0.04 | 0.03 | 2.92 | 0.04 | 53.05 | 0.04 | 0.03 | 2.94 | 0.04 | 53.45 | 0.03 | 0.02 | 2.25 | 0.03 | 40.83 | 0.05 | 0.03 | 3.34 | 0.04 | 60.66 |
| Epilepsy | 0.21 | -0.02 | 1.10 | 0.19 | 8.65 | 0.21 | 0.02 | 1.09 | 0.19 | 8.59 | 0.23 | 0.04 | 1.20 | 0.21 | 9.43 | 0.22 | 0.02 | 1.11 | 0.19 | 8.72 |
| Colorectal Cancer | 0.02 | 0.01 | 2.47 | 0.02 | 12.56 | 0.02 | 0.01 | 2.64 | 0.02 | 13.45 | 0.03 | 0.02 | 3.00 | 0.02 | 15.25 | 0.02 | 0.01 | 2.59 | 0.02 | 13.18 |

**Table S10. 1- and 5-year adjusted absolute risks (AR), split by age group, for each outcome, and the absolute risk increase (ARI) and relative risk ratios (RR) compared to the wider Scottish population**

|  | Stroke | | | | Dementia | | | | Parkinson’s Disease | | | | Epilepsy | | | | Colorectal cancer | | | |
| --- | --- | --- | --- | --- | --- | --- | --- | --- | --- | --- | --- | --- | --- | --- | --- | --- | --- | --- | --- | --- |
|  | Analytic AR | Population AR | ARI | RR  [95% CI] | Analytic AR | Population AR | ARI | RR  [95% CI] | Analytic AR | Population AR | ARI | RR  [95% CI] | Analytic AR | Population AR | ARI | RR  [95% CI] | Analytic AR | Population AR | ARI | RR  [95% CI] |
| **1-Year Survival** |  |  |  |  |  |  |  |  |  |  |  |  |  |  |  |  |  |  |  |  |
| 21-30 years | <0.01 | <0.01 | <0.01 | 13.22 [6.58-26.58] | <0.01 | <0.01 | <0.01 | 3.09 [0.68-13.94] | <0.01 | <0.01 | <0.01 | 11.33 [1.89-67.80] | 0.04 | 0.01 | 0.03 | 3.13 [2.97-3.31] | <0.01 | <0.01 | <0.01 | 0.53 [0.07-3.89] |
| 31-40 years | <0.01 | <0.01 | <0.01 | 8.68 [5.21-14.46] | <0.01 | <0.01 | <0.01 | 6.87 [2.99-15.80] | <0.01 | <0.01 | <0.01 | 5.80 [1.54-21.85] | 0.05 | 0.02 | 0.03 | 2.27 [2.16-2.38] | <0.01 | <0.01 | <0.01 | 0.40 [0.10-1.63] |
| 41-50 years | <0.01 | <0.01 | <0.01 | 9.36 [7.21-12.16] | <0.01 | <0.01 | <0.01 | 6.70 [4.45-10.07] | <0.01 | <0.01 | <0.01 | 4.59 [2.32-9.05] | 0.05 | 0.03 | 0.02 | 1.80 [1.73-1.88] | <0.01 | <0.01 | <0.01 | 0.65 [0.40-1.08] |
| 51-60 years | <0.01 | <0.01 | <0.01 | 6.94 [5.88-8.18] | <0.01 | <0.01 | <0.01 | 6.40 [5.53-7.42] | <0.01 | <0.01 | <0.01 | 3.73 [2.59-5.37] | 0.05 | 0.03 | 0.01 | 1.47 [1.41-1.53] | <0.01 | <0.01 | <0.01 | 0.47 [0.34-0.63] |
| 61-70 years | 0.01 | <0.01 | 0.01 | 4.63 [4.10-5.23] | 0.02 | 0.01 | 0.02 | 4.60 [4.30-4.92] | <0.01 | <0.01 | <0.01 | 2.84 [2.34-3.45] | 0.04 | 0.03 | 0.01 | 1.17 [1.12-1.22] | <0.01 | <0.01 | <0.01 | 0.53 [0.44-0.64] |
| 71-80 years | 0.02 | 0.01 | 0.01 | 2.82 [2.61-3.06] | 0.09 | 0.04 | 0.05 | 2.44 [2.36-2.52] | 0.01 | <0.01 | <0.01 | 1.58 [1.41-1.77] | 0.04 | 0.04 | <0.01 | 0.90 [0.86-0.94] | <0.01 | 0.01 | <0.01 | 0.46 [0.39-0.53] |
| 81-90 years | 0.03 | 0.02 | 0.01 | 1.64 [1.52-1.76] | 0.19 | 0.15 | 0.05 | 1.33 [1.29-1.36] | 0.01 | 0.01 | <0.01 | 0.84 [0.73-0.95] | 0.03 | 0.04 | -0.02 | 0.66 [0.61-0.70] | 0.01 | 0.01 | <0.01 | 0.51 [0.44-0.60] |
| >=91 years | 0.05 | 0.04 | 0.01 | 1.38 [1.20-1.58] | 0.21 | 0.22 | -0.01 | 0.93 [0.88-0.99] | <0.01 | 0.01 | <0.01 | 0.43 [0.26-0.70] | 0.02 | 0.04 | -0.02 | 0.58 [0.48-0.70] | <0.01 | 0.01 | -0.01 | 0.46 [0.31-0.70] |
| **5-Year Survival** |  |  |  |  |  |  |  |  |  |  |  |  |  |  |  |  |  |  |  |  |
| 21-30 years | <0.01 | <0.01 | <0.01 | 13.89 [9.11-21.17] | <0.01 | <0.01 | <0.01 | 3.85 [1.48-10.01] | <0.01 | <0.01 | <0.01 | 8.01 [2.85-22.45] | 0.16 | 0.04 | 0.12 | 4.42 [4.29-4.56] | <0.01 | <0.01 | <0.01 | 0.66 [0.24-1.77] |
| 31-40 years | <0.01 | <0.01 | <0.01 | 12.21 [9.35-15.93] | <0.01 | <0.01 | <0.01 | 4.82 [2.62-8.85] | <0.01 | <0.01 | <0.01 | 8.22 [3.91-17.26] | 0.20 | 0.06 | 0.14 | 3.25 [3.17-3.34] | <0.01 | <0.01 | <0.01 | 0.97 [0.58-1.60] |
| 41-50 years | 0.01 | <0.01 | 0.01 | 12.47 [10.93-14.22] | <0.01 | <0.01 | <0.01 | 7.48 [5.77-9.70] | <0.01 | <0.01 | <0.01 | 7.96 [5.61-11.30] | 0.22 | 0.08 | 0.14 | 2.71 [2.66-2.77] | <0.01 | <0.01 | <0.01 | 0.84 [0.66-1.07] |
| 51-60 years | 0.02 | <0.01 | 0.02 | 9.71 [8.92-10.56] | 0.02 | <0.01 | 0.01 | 8.54 [7.78-9.39] | <0.01 | <0.01 | <0.01 | 5.61 [4.69-6.72] | 0.21 | 0.09 | 0.12 | 2.31 [2.26-2.36] | <0.01 | 0.01 | <0.01 | 0.80 [0.70-0.93] |
| 61-70 years | 0.04 | 0.01 | 0.03 | 7.10 [6.68-7.55] | 0.09 | 0.01 | 0.07 | 6.76 [6.48-7.04] | 0.02 | <0.01 | 0.01 | 4.95 [4.52-5.43] | 0.19 | 0.10 | 0.09 | 1.92 [1.88-1.97] | 0.01 | 0.01 | <0.01 | 0.87 [0.79-0.95] |
| 71-80 years | 0.10 | 0.02 | 0.08 | 4.25 [4.08-4.42] | 0.42 | 0.10 | 0.31 | 3.99 [3.92-4.07] | 0.05 | 0.02 | 0.03 | 2.93 [2.76-3.11] | 0.20 | 0.12 | 0.07 | 1.57 [1.53-1.61] | 0.02 | 0.03 | -0.01 | 0.78 [0.72-0.84] |
| 81-90 years | 0.24 | 0.10 | 0.14 | 2.39 [2.31-2.48] | 1.32 | 0.61 | 0.71 | 2.17 [NA-NA] | 0.06 | 0.04 | 0.02 | 1.52 [1.41-1.64] | 0.20 | 0.17 | -0.04 | 1.23 [1.18-1.28] | 0.04 | 0.05 | -0.01 | 0.74 [0.68-0.81] |
| >=91 years | 0.64 | 0.31 | 0.33 | 2.07 [1.95-2.19] | 2.73 | 1.72 | 1.01 | 1.59 [NA-NA] | 0.04 | 0.05 | -0.01 | 0.77 [0.56-1.06] | 0.25 | 0.21 | -0.03 | 1.16 [1.03-1.31] | 0.06 | 0.08 | -0.01 | 0.81 [0.63-1.05] |

**Figure S1. Kaplan-Meier curves showing stroke survival for those with and without WMH, lacunes, cortical infarcts, cerebral atrophy, and any/no CCD phenotype**

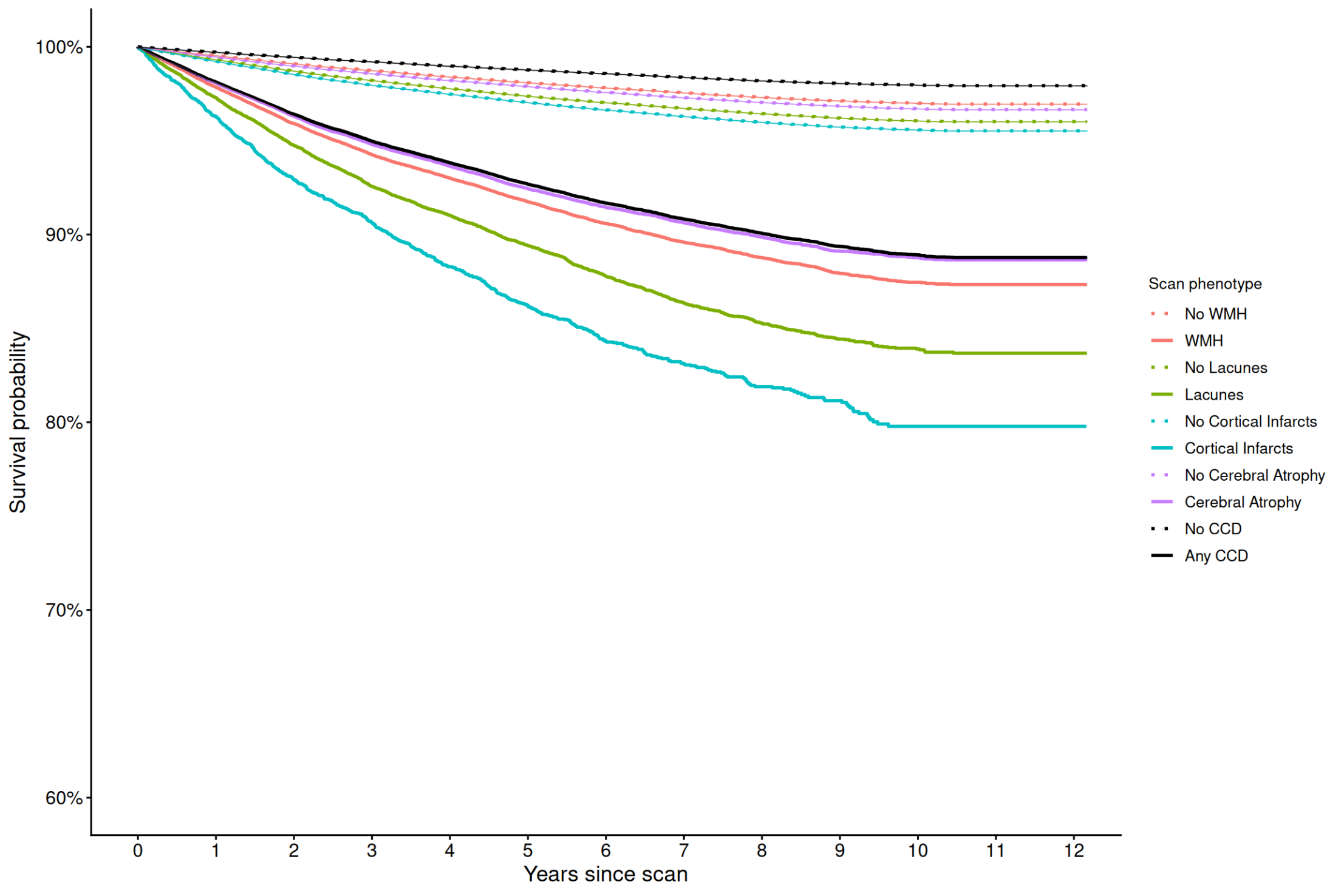

**Figure S2. Kaplan-Meier curves showing dementia survival for those with and without WMH, lacunes, cortical infarcts, cerebral atrophy, and any/no CCD phenotype**
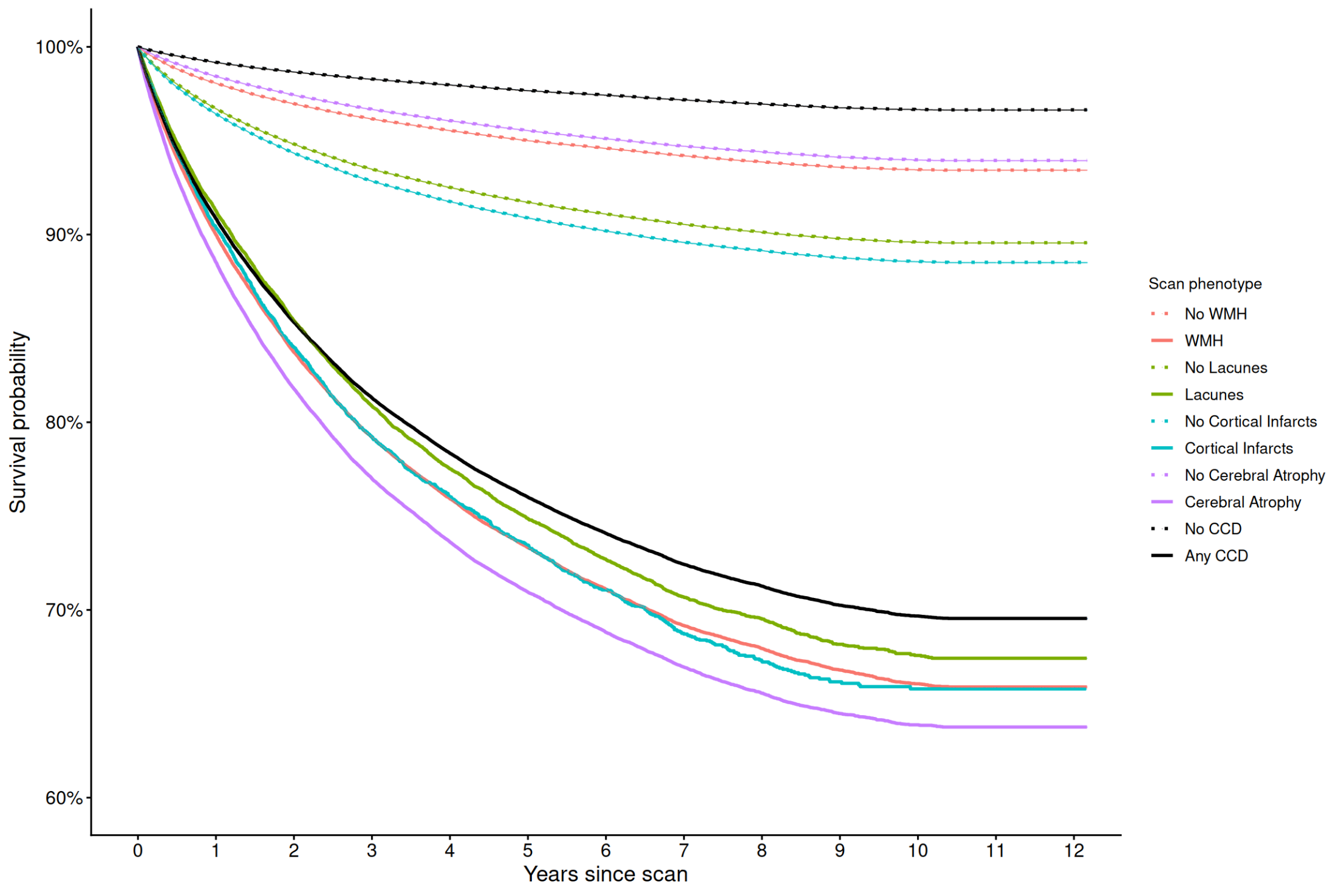

**Figure S3. Kaplan-Meier curves showing Parkinson’s disease survival for those with and without WMH, lacunes, cortical infarcts, cerebral atrophy, and any/no CCD phenotype**

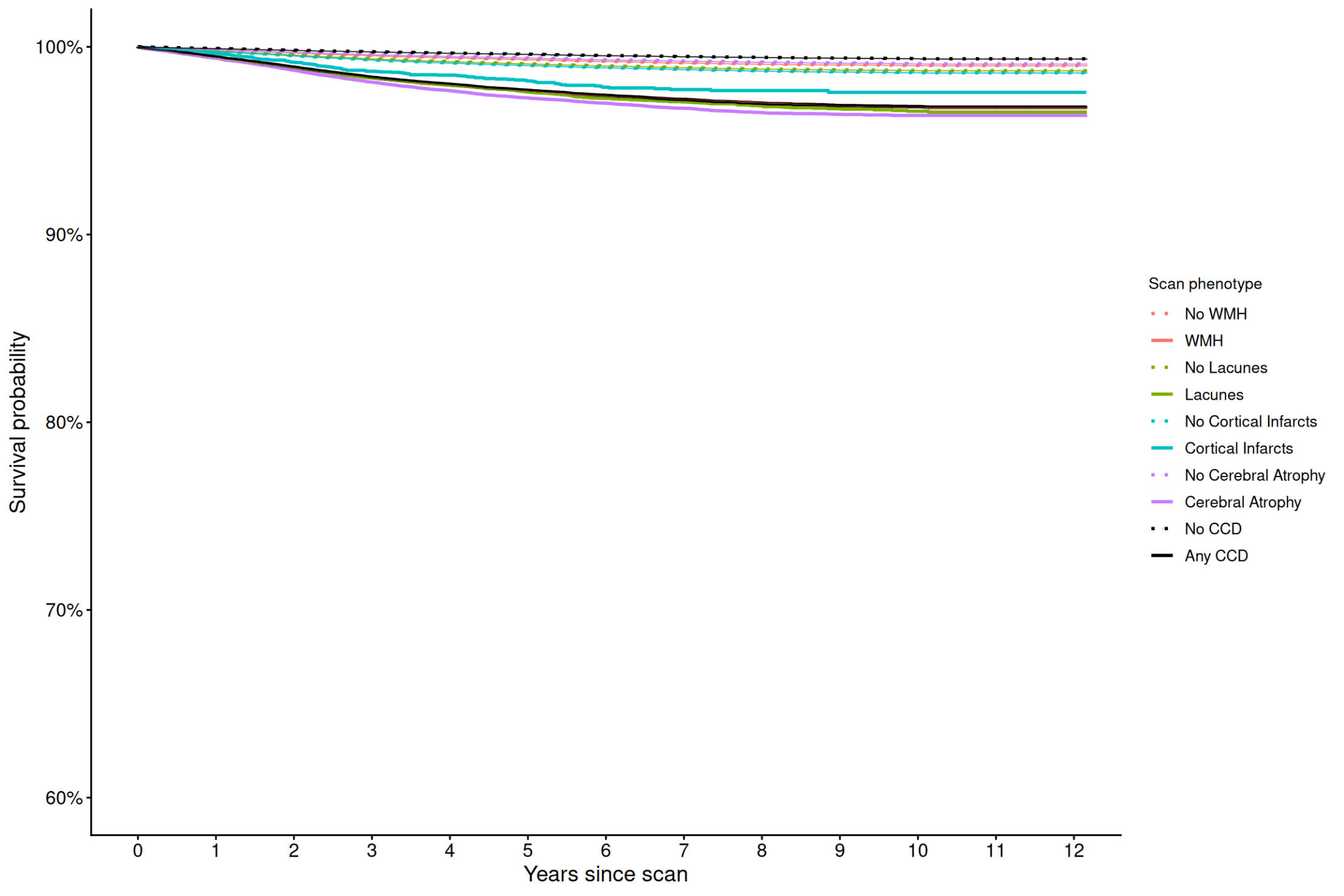

**Figure S4. Kaplan-Meier curves showing epilepsy survival for those with and without WMH, lacunes, cortical infarcts, cerebral atrophy, and any/no CCD phenotype**
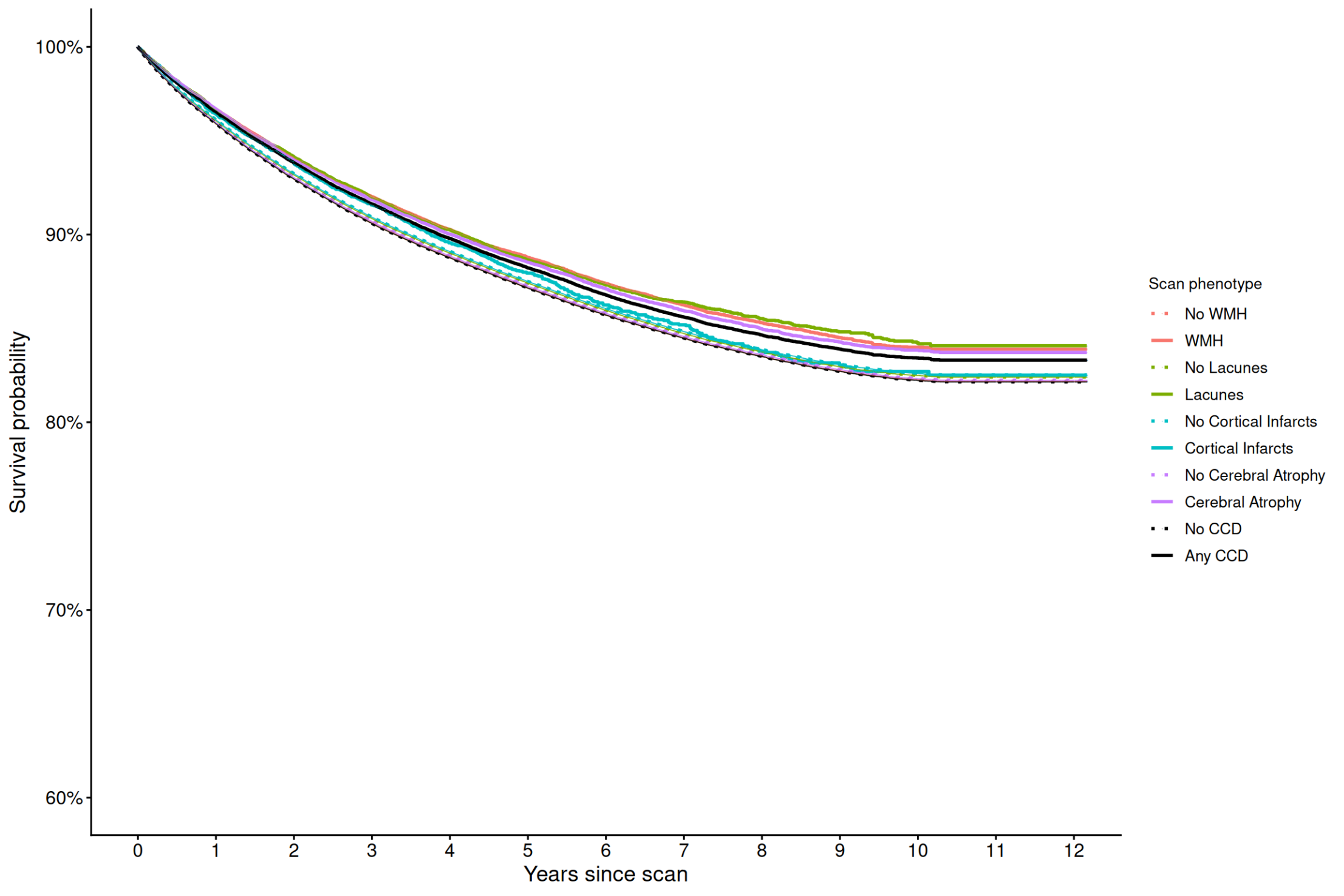

**Figure S5. Kaplan-Meier curves showing colorectal cancer survival for those with and without WMH, lacunes, cortical infarcts, cerebral atrophy, and any/no CCD phenotype**
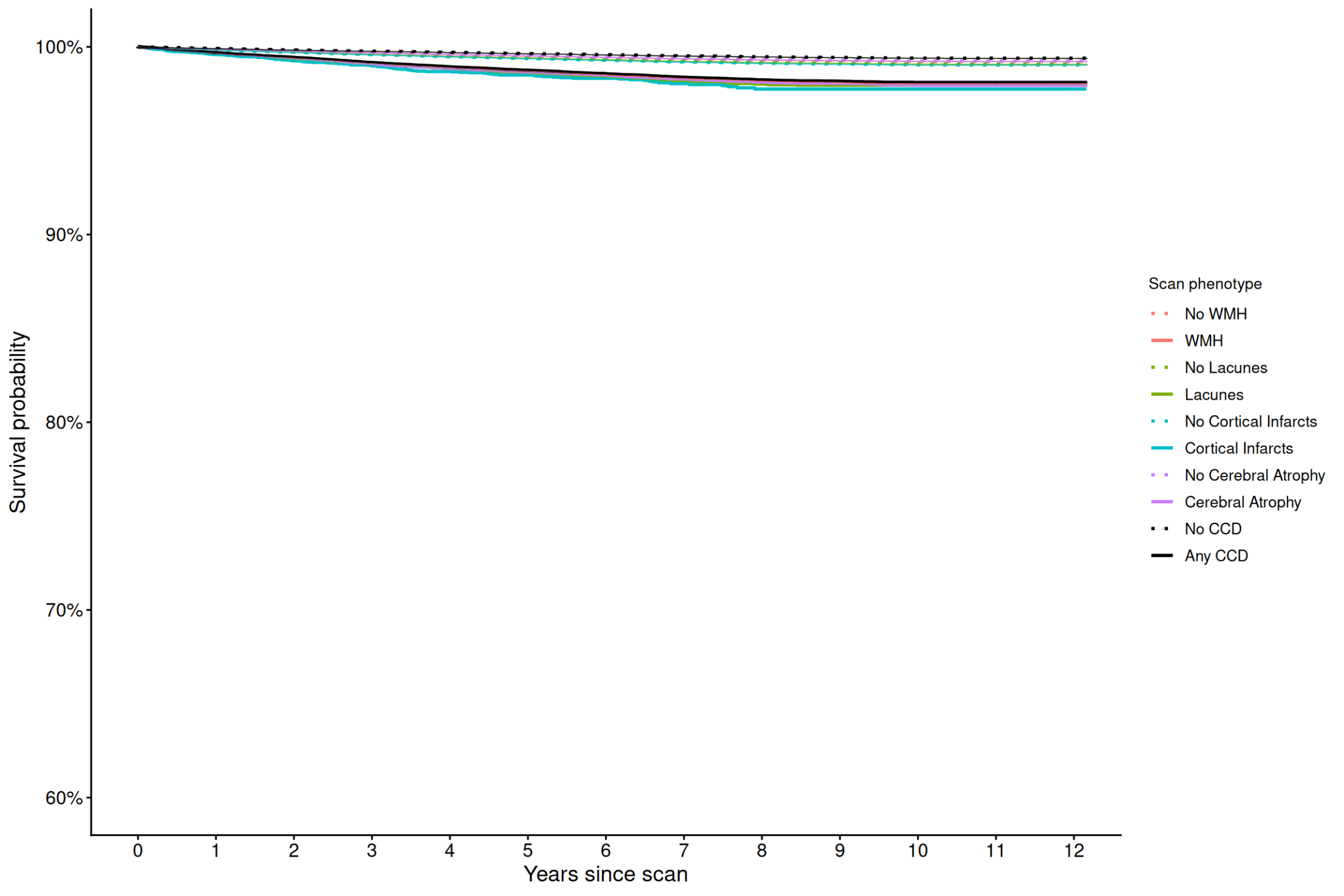

**Table S11. Univariate hazard ratios (HR) and fully-adjusted hazard ratios (aHR), 95% CIs and p-values for each scan phenotype associated with each outcome in the primary analyses**

| **Outcome** | **Scan phenotype** | **HR [95% CI]** | **aHR [95% CI]** | **p-value** |
| --- | --- | --- | --- | --- |
| Stroke | WMH | 4.4 [4.3–4.6] | 1.4 [1.3–1.4] | <0.001 |
|  | Lacune | 4.3 [4.1–4.5] | 1.6 [1.5–1.6] | <0.001 |
|  | Cortical infarct | 4.9 [4.6–5.2] | 1.8 [1.7–1.9] | <0.001 |
|  | Cerebral atrophy | 3.6 [3.5–3.7] | 1.1 [1.0–1.1] | <0.001 |
| Dementia | WMH | 6.1 [5.9–6.2] | 1.3 [1.3–1.3] | <0.001 |
|  | Lacune | 3.3 [3.3–3.4] | 1.0 [1.0–1.0] | 0.37 |
|  | Cortical infarct | 3.3 [3.1–3.4] | 1.1 [1.1–1.2] | <0.001 |
|  | Cerebral atrophy | 7.4 [7.3–7.6] | 1.7 [1.7–1.8] | <0.001 |
| Parkinson’s disease | WMH | 3.4 [3.2–3.6] | 1.1 [1.0–1.2] | 0.03 |
|  | Lacune | 2.7 [2.5–2.9] | 1.1 [1.0–1.2] | 0.16 |
|  | Cortical infarct | 1.8 [1.5–2.2] | 0.7 [0.6–0.9] | <0.01 |
|  | Cerebral atrophy | 4.4 [4.2–4.7] | 1.4 [1.3–1.5] | <0.001 |
| Epilepsy | WMH | 0.9 [0.9–0.9] | 1.0 [1.0–1.0] | 0.83 |
|  | Lacune | 0.9 [0.9–0.9] | 1.0 [0.9–1.0] | 0.04 |
|  | Cortical infarct | 1.0 [0.9–1.0] | 1.0 [1.0–1.1] | 0.55 |
|  | Cerebral atrophy | 0.9 [0.9–0.9] | 1.0 [1.0–1.0] | 0.49 |
| Colorectal cancer | WMH | 2.6 [2.4–2.9] | 0.9 [0.9–1.0] | 0.23 |
|  | Lacune | 2.4 [2.1–2.6] | 1.1 [0.9–1.2] | 0.48 |
|  | Cortical infarct | 2.5 [2.0–3.0] | 1.1 [0.9–1.4] | 0.27 |
|  | Cerebral atrophy | 2.8 [2.6–3.1] | 1.0 [0.9–1.1] | 0.60 |

**Table S12. Univariate hazard ratios (HR) and fully-adjusted hazard ratios (aHR), 95% CIs and p-values for each scan phenotype associated with stroke and dementia subtypes in the primary analyses**

| **Outcome** | **Scan phenotype** | **HR [95% CI]** | **aHR [95% CI]** | **p-value** |
| --- | --- | --- | --- | --- |
| Ischaemic stroke | WMH | 4.0 [3.8–4.2] | 1.3 [1.3–1.4] | <0.001 |
|  | Lacune | 4.1 [3.9–4.3] | 1.5 [1.4–1.6] | <0.001 |
|  | Cortical infarct | 4.9 [4.5–5.4] | 1.9 [1.7–2.0] | <0.001 |
|  | Cerebral atrophy | 3.2 [3.1–3.4] | 1.0 [1.0–1.1] | 0.38 |
| Intracerebral haemorrhage | WMH | 4.4 [4.2–4.7] | 1.5 [1.4–1.6] | <0.001 |
|  | Lacune | 4.2 [3.9–4.5] | 1.6 [1.5–1.7] | <0.001 |
|  | Cortical infarct | 4.4 [3.9–4.9] | 1.7 [1.5–1.9] | <0.001 |
|  | Cerebral atrophy | 3.3 [3.1–3.5] | 1.1 [1.0–1.1] | 0.06 |
| Uncertain stroke | WMH | 4.8 [4.5–5.1] | 1.3 [1.2–1.4] | <0.001 |
|  | Lacune | 4.5 [4.2–4.9] | 1.6 [1.4–1.7] | <0.001 |
|  | Cortical infarct | 5.0 [4.5–5.5] | 1.8 [1.6–2.0] | <0.001 |
|  | Cerebral atrophy | 4.2 [4.0–4.5] | 1.2 [1.1–1.3] | <0.001 |
| Alzheimer’s dementia | WMH | 4.9 [4.6–5.3] | 0.9 [0.9–1.0] | 0.15 |
|  | Lacune | 2.3 [2.0–2.5] | 0.7 [0.7–0.8] | <0.001 |
|  | Cortical infarct | 1.8 [1.5–2.2] | 0.7 [0.6–0.8] | <0.001 |
|  | Cerebral atrophy | 8.3 [7.7–8.9] | 1.9 [1.8–2.1] | <0.001 |
| Vascular dementia | WMH | 9.6 [9.2–10.0] | 2.0 [1.9–2.1] | <0.001 |
|  | Lacune | 6.0 [5.7–6.3] | 1.6 [1.5–1.7] | <0.001 |
|  | Cortical infarct | 6.0 [5.6–6.4] | 1.8 [1.7–2.0] | <0.001 |
|  | Cerebral atrophy | 7.1 [6.8–7.4] | 1.4 [1.3–1.4] | <0.001 |
| Unspecified/rare dementia | WMH | 6.5 [6.2–6.7] | 1.3 [1.3–1.4] | <0.001 |
|  | Lacune | 3.4 [3.3–3.6] | 1.0 [1.0–1.1] | 0.60 |
|  | Cortical infarct | 3.6 [3.3–3.8] | 1.2 [1.1–1.3] | <0.001 |
|  | Cerebral atrophy | 7.1 [6.8–7.4] | 1.5 [1.4–1.6] | <0.001 |

**Table S13. Adjusted hazard ratios (aHR), 95% CIs and p-values for each scan phenotype associated with each outcome after removing the first year of follow-up**

| **Outcome** | **Scan phenotype** | **sHR [95% CI]** | **p-value** |
| --- | --- | --- | --- |
| Stroke | WMH | 1.3 [1.2–1.4] | <0.001 |
|  | Lacune | 1.6 [1.5–1.6] | <0.001 |
|  | Cortical infarct | 1.6 [1.5–1.7] | <0.001 |
|  | Cerebral atrophy | 1.0 [1.0–1.1] | 0.05 |
| Dementia | WMH | 1.2 [1.2–1.3] | <0.001 |
|  | Lacune | 1.0 [1.0–1.0] | 0.90 |
|  | Cortical infarct | 1.0 [1.0–1.1] | 0.52 |
|  | Cerebral atrophy | 1.6 [1.6–1.7] | <0.001 |
| Parkinson’s disease | WMH | 1.1 [1.0–1.1] | 0.19 |
|  | Lacune | 1.0 [0.9–1.1] | 0.49 |
|  | Cortical infarct | 0.7 [0.6–0.9] | <0.001 |
|  | Cerebral atrophy | 1.4 [1.3–1.5] | <0.001 |
| Epilepsy | WMH | 1.0 [1.0–1.0] | 0.73 |
|  | Lacune | 0.9 [0.9–1.0] | <0.01 |
|  | Cortical infarct | 1.0 [0.9–1.0] | 0.33 |
|  | Cerebral atrophy | 1.0 [1.0–1.0] | 0.87 |
| Colorectal cancer | WMH | 0.9 [0.8–1.0] | 0.14 |
|  | Lacune | 1.1 [0.9–1.2] | 0.40 |
|  | Cortical infarct | 1.0 [0.8–1.2] | 0.73 |
|  | Cerebral atrophy | 1.0 [0.9–1.1] | 0.64 |

**Table S14. Adjusted hazard ratios (aHR), 95% CIs and p-values for each scan phenotype associated with each outcome after removing the first 5 years of follow-up**

| **Outcome** | **Scan phenotype** | **aHR [95% CI]** | **p-value** |
| --- | --- | --- | --- |
| Stroke | WMH | 1.2 [1.1–1.3] | <0.001 |
|  | Lacune | 1.5 [1.4–1.7] | <0.001 |
|  | Cortical infarct | 1.3 [1.1–1.6] | <0.001 |
|  | Cerebral atrophy | 0.9 [0.8–1.0] | 0.27 |
| Dementia | WMH | 1.1 [1.1–1.1] | <0.001 |
|  | Lacune | 0.9 [0.9–1.0] | <0.001 |
|  | Cortical infarct | 0.9 [0.8–1.0] | 0.04 |
|  | Cerebral atrophy | 1.4 [1.3–1.5] | <0.001 |
| Parkinson’s disease | WMH | 0.9 [0.8–1.0] | 0.04 |
|  | Lacune | 1.0 [0.8–1.2] | 0.75 |
|  | Cortical infarct | 0.5 [0.4–0.8] | <0.01 |
|  | Cerebral atrophy | 1.3 [1.1–1.4] | <0.001 |
| Epilepsy | WMH | 1.0 [0.9–1.0] | 0.13 |
|  | Lacune | 0.9 [0.8–1.0] | <0.01 |
|  | Cortical infarct | 0.9 [0.8–1.1] | 0.23 |
|  | Cerebral atrophy | 0.9 [0.9–1.0] | <0.01 |
| Colorectal cancer | WMH | 0.9 [0.7–1.0] | 0.10 |
|  | Lacune | 1.1 [0.8–1.3] | 0.67 |
|  | Cortical infarct | 0.7 [0.4–1.1] | 0.15 |
|  | Cerebral atrophy | 0.9 [0.8–1.1] | 0.29 |

**Table S15. Adjusted hazard Ratios (aHR), 95% CIs and p-values for each scan phenotype associated with each outcome, broken down by sex, age group and scan modality**

|  |  | **Main effect** | | **Interaction effect** | |
| --- | --- | --- | --- | --- | --- |
| **Outcome** | **Phenotype** | **aHR [95% CI]** | **p-value** | **aHR [95% CI]** | **p-value** |
| **Stroke** | **Sex** |  |  |  |  |
|  | WMH | 1.3 [1.2–1.4] | <0.001 | 1.1 [1.0–1.2] | <0.01 |
|  | Lacunes | 1.6 [1.5–1.7] | <0.001 | 0.9 [0.9–1.0] | 0.16 |
|  | Cortical Infarcts | 1.7 [1.5–1.8] | <0.001 | 1.2 [1.0–1.3] | 0.02 |
|  | Cerebral atrophy | 1.0 [1.0–1.1] | 0.52 | 1.1 [1.0–1.2] | <0.01 |
|  | *Male* |  |  |  |  |
|  | WMH | 1.4 [1.3–1.4] | <0.001 | - | - |
|  | Lacunes | 1.7 [1.6–1.8] | <0.001 | - | - |
|  | Cortical Infarcts | 1.7 [1.6–1.9] | <0.001 | - | - |
|  | Cerebral atrophy | 1.1 [1.0–1.2] | <0.01 | - | - |
|  | *Female* |  |  |  |  |
|  | WMH | 1.4 [1.3–1.4] | <0.001 | - | - |
|  | Lacunes | 1.5 [1.4–1.6] | <0.001 | - | - |
|  | Cortical Infarcts | 1.9 [1.8–2.1] | <0.001 | - | - |
|  | Cerebral atrophy | 1.1 [1.0–1.1] | <0.01 | - | - |
|  | **Age (linear)** |  |  |  |  |
|  | WMH | 3.7 [2.7–5.0] | <0.001 | 1.1 [1.0–1.1] | <0.001 |
|  | Lacunes | 9.6 [6.8–13.5] | <0.001 | 1.0 [1.0–1.0] | <0.001 |
|  | Cortical Infarcts | 3.4 [2.0–5.9] | <0.001 | 1.0 [1.0–1.0] | 0.01 |
|  | Cerebral atrophy | 1.9 [1.4–2.6] | <0.001 | 1.0 [1.0–1.0] | <0.001 |
|  | *Age <= 30 years* |  |  |  |  |
|  | WMH | NA [NA–NA] | NA | - | - |
|  | Lacunes | NA [NA–NA] | NA | - | - |
|  | Cortical Infarcts | NA [NA–NA] | NA | - | - |
|  | Cerebral atrophy | 4.5 [1.1–18.8] | 0.04 | - | - |
|  | *Age 31-50 years* |  |  |  |  |
|  | WMH | 2.2 [1.6–3.0] | <0.001 | - | - |
|  | Lacunes | 3.1 [2.1–4.5] | <0.001 | - | - |
|  | Cortical Infarcts | 2.8 [1.6–4.9] | <0.001 | - | - |
|  | Cerebral atrophy | 1.4 [1.0–1.9] | 0.04 | - | - |
|  | *Ages 51-70 years* |  |  |  |  |
|  | WMH | 1.7 [1.6–1.8] | <0.001 | - | - |
|  | Lacunes | 2.3 [2.0–2.5] | <0.001 | - | - |
|  | Cortical Infarcts | 1.9 [1.7–2.3] | <0.001 | - | - |
|  | Cerebral atrophy | 1.3 [1.2–1.4] | <0.001 | - | - |
|  | *Ages 71-90 years* |  |  |  |  |
|  | WMH | 1.4 [1.3–1.4] | <0.001 | - | - |
|  | Lacunes | 1.5 [1.4–1.6] | <0.001 | - | - |
|  | Cortical Infarcts | 1.9 [1.7–2.0] | <0.001 | - | - |
|  | Cerebral atrophy | 1.1 [1.1–1.2] | <0.001 | - | - |
|  | *Ages >=91 years* |  |  |  |  |
|  | WMH | 1.0 [0.9–1.2] | 0.68 | - | - |
|  | Lacunes | 1.1 [0.9–1.3] | 0.27 | - | - |
|  | Cortical Infarcts | 1.5 [1.1–2.0] | <0.01 | - | - |
|  | Cerebral atrophy | 1.0 [0.8–1.2] | 0.94 | - | - |
|  | **Modality** |  |  |  |  |
|  | WMH | 1.4 [1.3–1.4] | <0.001 | 1.0 [0.9–1.1] | 0.92 |
|  | Lacunes | 1.5 [1.5–1.6] | <0.001 | 1.1 [1.1–1.5] | <0.01 |
|  | Cortical Infarcts | 1.8 [1.7–1.9] | <0.001 | 1.0 [0.7–1.2] | 0.70 |
|  | Cerebral atrophy | 1.1 [1.0–1.1] | <0.01 | 1.2 [1.1–1.4] | <0.001 |
|  | *CT* |  |  |  |  |
|  | WMH | 1.4 [1.3–1.4] | <0.001 | - | - |
|  | Lacunes | 1.5 [1.5–1.6] | <0.001 | - | - |
|  | Cortical Infarcts | 1.8 [1.7–1.9] | <0.001 | - | - |
|  | Cerebral atrophy | 1.1 [1.0–1.1] | 0.02 | - | - |
|  | *MRI* |  |  |  |  |
|  | WMH | 1.4 [1.3–1.6] | <0.001 | - | - |
|  | Lacunes | 1.9 [1.7–2.3] | <0.001 | - | - |
|  | Cortical Infarcts | 1.7 [1.4–2.2] | <0.001 | - | - |
|  | Cerebral atrophy | 1.3 [1.2–1.5] | <0.001 | - | - |
| **Dementia** | **Sex** |  |  |  |  |
|  | WMH | 1.3 [1.3–1.4] | <0.001 | 1.0 [0.9–1.0] | 0.04 |
|  | Lacunes | 1.0 [1.0–1.1] | 0.52 | 1.0 [0.9–1.1] | 0.91 |
|  | Cortical Infarcts | 1.2 [1.1–1.2] | <0.01 | 1.0 [0.9–1.1] | 0.40 |
|  | Cerebral atrophy | 1.8 [1.7–1.9] | <0.001 | 0.9 [0.9–1.0] | <0.01 |
|  | *Male* |  |  |  |  |
|  | WMH | 1.3 [1.3–1.4] | <0.001 | - | - |
|  | Lacunes | 1.0 [1.0–1.1] | 0.48 | - | - |
|  | Cortical Infarcts | 1.2 [1.1–1.2] | <0.01 | - | - |
|  | Cerebral atrophy | 1.8 [1.7–1.9] | <0.001 | - | - |
|  | *Female* |  |  |  |  |
|  | WMH | 1.3 [1.2–1.3] | <0.001 | - | - |
|  | Lacunes | 1.0 [1.0–1.0] | 0.61 | - | - |
|  | Cortical Infarcts | 1.1 [1.0–1.2] | <0.01 | - | - |
|  | Cerebral atrophy | 1.7 [1.6–1.7] | <0.001 | - | - |
|  | **Age (linear)** |  |  |  |  |
|  | WMH | 3.9 [3.1–5.0] | <0.001 | 1.1 [1.1–1.1] | <0.001 |
|  | Lacunes | 1.2 [0.9–1.7] | 0.22 | 1.0 [1.0–1.0] | 0.23 |
|  | Cortical Infarcts | 2.5 [1.5–4.2] | <0.001 | 1.0 [1.0–1.0] | <0.01 |
|  | Cerebral atrophy | 19.6 [15.7–24.5] | <0.001 | 1.0 [1.0–1.0] | <0.001 |
|  | *Age <= 30 years* |  |  |  |  |
|  | WMH | NA [NA–NA] | NA | - | - |
|  | Lacunes | NA [NA–NA] | NA | - | - |
|  | Cortical Infarcts | NA [NA–NA] | NA | - | - |
|  | Cerebral atrophy | 18.0 [1.5–208.7] | 0.02 | - | - |
|  | *Age 31-50 years* |  |  |  |  |
|  | WMH | 4.8 [2.7–8.4] | <0.001 | - | - |
|  | Lacunes | 2.5 [1.1–5.6] | 0.03 | - | - |
|  | Cortical Infarcts | 1.6 [0.4–7.0] | 0.54 | - | - |
|  | Cerebral atrophy | 6.5 [4.1–10.2] | <0.001 | - | - |
|  | *Age 51-70 years* |  |  |  |  |
|  | WMH | 1.9 [1.8–2.1] | <0.001 | - | - |
|  | Lacunes | 1.2 [1.1–1.3] | <0.01 | - | - |
|  | Cortical Infarcts | 1.5 [1.3–1.7] | <0.001 | - | - |
|  | Cerebral atrophy | 3.6 [3.4–3.9] | <0.001 | - | - |
|  | *Age 71-90 years* |  |  |  |  |
|  | WMH | 1.4 [1.4–1.4] | <0.001 | - | - |
|  | Lacunes | 1.0 [1.0–1.1] | 0.09 | - | - |
|  | Cortical Infarcts | 1.2 [1.1–1.2] | <0.001 | - | - |
|  | Cerebral atrophy | 1.7 [1.7–1.8] | <0.001 | - | - |
|  | *Age >= 91 years* |  |  |  |  |
|  | WMH | 1.1 [1.0–1.3] | 0.01 | - | - |
|  | Lacunes | 1.1 [1.0–1.2] | 0.20 | - | - |
|  | Cortical Infarcts | 0.9 [0.8–1.1] | 0.30 | - | - |
|  | Cerebral atrophy | 1.3 [1.2–1.4] | <0.001 | - | - |
|  | **Modality** |  |  |  |  |
|  | WMH | 1.3 [1.3–1.3] | <0.001 | 1.0 [0.9–1.0] | 0.33 |
|  | Lacunes | 1.0 [1.0–1.0] | 0.44 | 1.0 [0.9–1.2] | 0.81 |
|  | Cortical Infarcts | 1.1 [1.1–1.2] | <0.001 | 1.1 [0.9–1.4] | 0.26 |
|  | Cerebral atrophy | 1.7 [1.7–1.7] | <0.001 | 1.2 [1.1–1.3] | <0.001 |
|  | *CT* |  |  |  |  |
|  | WMH | 1.3 [1.3–1.3] | <0.001 | - | - |
|  | Lacunes | 1.0 [1.0–1.0] | 0.47 | - | - |
|  | Cortical Infarcts | 1.1 [1.1–1.2] | <0.001 | - | - |
|  | Cerebral atrophy | 1.7 [1.7–1.7] | <0.001 | - | - |
|  | *MRI* |  |  |  |  |
|  | WMH | 1.3 [1.2–1.5] | <0.001 | - | - |
|  | Lacunes | 1.0 [0.9–1.2] | 0.77 | - | - |
|  | Cortical Infarcts | 1.3 [1.0–1.6] | 0.03 | - | - |
|  | Cerebral atrophy | 2.1 [1.9–2.4] | <0.001 | - | - |
| **Epilepsy** | **Sex** |  |  |  |  |
|  | WMH | 1.0 [1.0–1.1] | 0.09 | 0.9 [0.9–1.0] | 0.05 |
|  | Lacunes | 1.0 [0.9–1.1] | 0.74 | 0.9 [0.9–1.0] | 0.09 |
|  | Cortical Infarcts | 1.0 [0.9–1.1] | 0.66 | 1.0 [0.9–1.1] | 0.88 |
|  | Cerebral atrophy | 1.1 [1.1–1.2] | <0.001 | 0.8 [0.8–0.9] | <0.001 |
|  | *Males* |  |  |  |  |
|  | WMH | 1.0 [1.0–1.1] | 0.29 | - | - |
|  | Lacunes | 1.0 [0.9–1.0] | 0.54 | - | - |
|  | Cortical Infarcts | 1.0 [0.9–1.1] | 0.73 | - | - |
|  | Cerebral atrophy | 1.1 [1.0–1.1] | <0.001 | - | - |
|  | *Females* |  |  |  |  |
|  | WMH | 1.0 [1.0–1.0] | 0.69 | - | - |
|  | Lacunes | 0.9 [0.9–1.0] | 0.01 | - | - |
|  | Cortical Infarcts | 1.0 [0.9–1.1] | 0.78 | - | - |
|  | Cerebral atrophy | 0.9 [0.9–1.0] | <0.01 | - | - |
|  | **Age (linear)** |  |  |  |  |
|  | WMH | 1.2 [1.0–1.5] | 0.03 | 1.0 [1.0–1.0] | <0.01 |
|  | Lacunes | 1.0 [0.8–1.2] | 0.77 | 1.0 [1.0–1.0] | 0.95 |
|  | Cortical Infarcts | 0.8 [0.5–1.2] | 0.32 | 1.0 [1.0–1.0] | 0.26 |
|  | Cerebral atrophy | 1.9 [1.7–2.2] | <0.001 | 1.0 [1.0–1.0] | <0.001 |
|  | *Age <= 30 years* |  |  |  |  |
|  | WMH | 1.6 [1.1–2.4] | 0.01 | - | - |
|  | Lacunes | 0.8 [0.4–1.6] | 0.48 | - | - |
|  | Cortical Infarcts | 2.4 [0.8–7.5] | 0.13 | - | - |
|  | Cerebral atrophy | 1.2 [0.9–1.5] | 0.31 | - | - |
|  | *Age 31-50 years* |  |  |  |  |
|  | WMH | 1.0 [0.9–1.1] | 0.89 | - | - |
|  | Lacunes | 0.9 [0.7–1.0] | 0.14 | - | - |
|  | Cortical Infarcts | 1.0 [0.8–1.4] | 0.78 | - | - |
|  | Cerebral atrophy | 1.3 [1.2–1.4] | <0.001 | - | - |
|  | *Age 51-70 years* |  |  |  |  |
|  | WMH | 1.0 [1.0–1.0] | 0.99 | - | - |
|  | Lacunes | 1.0 [0.9–1.0] | 0.19 | - | - |
|  | Cortical Infarcts | 0.9 [0.7–1.0] | 0.01 | - | - |
|  | Cerebral atrophy | 1.0 [1.0–1.1] | 0.64 | - | - |
|  | *Age 71-90 years* |  |  |  |  |
|  | WMH | 0.9 [0.9–1.0] | 0.02 | - | - |
|  | Lacunes | 0.9 [0.9–1.0] | 0.06 | - | - |
|  | Cortical Infarcts | 1.1 [1.0–1.2] | 0.04 | - | - |
|  | Cerebral atrophy | 0.9 [0.8–0.9] | <0.001 | - | - |
|  | *Age >= 91 years* |  |  |  |  |
|  | WMH | 1.0 [0.7–1.3] | 0.88 | - | - |
|  | Lacunes | 1.0 [0.7–1.3] | 0.82 | - | - |
|  | Cortical Infarcts | 1.2 [0.7–1.9] | 0.50 | - | - |
|  | Cerebral atrophy | 1.1 [0.9–1.5] | 0.34 | - | - |
|  | **Modality** |  |  |  |  |
|  | WMH | 1.0 [1.0–1.0] | 0.40 | 1.1 [1.0–1.1] | 0.03 |
|  | Lacunes | 1.0 [0.9–1.0] | 0.12 | 1.0 [0.8–1.1] | 0.54 |
|  | Cortical Infarcts | 1.0 [0.9–1.1] | 0.61 | 1.0 [0.8–1.3] | 0.85 |
|  | Cerebral atrophy | 1.0 [1.0–1.0] | 0.61 | 1.1 [1.0–1.2] | <0.01 |
|  | *CT* |  |  |  |  |
|  | WMH | 1.0 [1.0–1.0] | 0.46 | - | - |
|  | Lacunes | 1.0 [0.9–1.0] | 0.13 | - | - |
|  | Cortical Infarcts | 1.0 [0.9–1.1] | 0.62 | - | - |
|  | Cerebral atrophy | 1.0 [1.0–1.0] | 0.58 | - | - |
|  | *MRI* |  |  |  |  |
|  | WMH | 1.1 [1.0–1.1] | 0.06 | - | - |
|  | Lacunes | 0.9 [0.8–1.0] | 0.19 | - | - |
|  | Cortical Infarcts | 1.0 [0.9–1.3] | 0.71 | - | - |
|  | Cerebral atrophy | 1.1 [1.0–1.2] | <0.01 | - | - |
| **Parkinson’s disease** | **Sex** |  |  |  |  |
|  | WMH | 1.1 [1.0–1.2] | 0.01 | 0.9 [0.8–1.1] | 0.20 |
|  | Lacunes | 1.1 [1.0–1.3] | 0.02 | 0.8 [0.7–1.0] | 0.05 |
|  | Cortical Infarcts | 0.6 [0.5–0.8] | <0.001 | 1.7 [1.1–2.4] | <0.01 |
|  | Cerebral atrophy | 1.5 [1.4–1.7] | <0.001 | 0.9 [0.7–1.0] | 0.03 |
|  | *Males* |  |  |  |  |
|  | WMH | 1.1 [1.0–1.2] | 0.02 | - | - |
|  | Lacunes | 1.1 [1.0–1.3] | 0.03 | - | - |
|  | Cortical Infarcts | 0.6 [0.5–0.8] | <0.001 | - | - |
|  | Cerebral atrophy | 1.5 [1.4–1.6] | <0.001 | - | - |
|  | *Females* |  |  |  |  |
|  | WMH | 1.1 [0.9–1.2] | 0.41 | - | - |
|  | Lacunes | 1.0 [0.8–1.1] | 0.55 | - | - |
|  | Cortical Infarcts | 1.0 [0.8–1.4] | 0.80 | - | - |
|  | Cerebral atrophy | 1.3 [1.2–1.5] | <0.001 | - | - |
|  | **Age (linear)** |  |  |  |  |
|  | WMH | 3.1 [1.6–6.0] | <0.01 | 1.0 [1.0–1.1] | <0.01 |
|  | Lacunes | 0.6 [0.2–1.5] | 0.25 | 1.0 [1.0–1.0] | 0.19 |
|  | Cortical Infarcts | 0.2 [<0.1–1.7] | 0.15 | 1.0 [1.0–1.0] | 0.26 |
|  | Cerebral atrophy | 12.4 [6.8–22.7] | <0.001 | 1.0 [1.0–1.0] | <0.001 |
|  | *Age <= 30 years* |  |  |  |  |
|  | WMH | NA [NA–NA] | NA | - | - |
|  | Lacunes | NA [NA–NA] | NA | - | - |
|  | Cortical Infarcts | NA [NA–NA] | NA | - | - |
|  | Cerebral atrophy | NA [NA–NA] | NA | - | - |
|  | *Age 31-50 years* |  |  |  |  |
|  | WMH | 1.4 [0.4–4.6] | 0.55 | - | - |
|  | Lacunes | NA [NA–NA] | NA | - | - |
|  | Cortical Infarcts | NA [NA–NA] | NA | - | - |
|  | Cerebral atrophy | 3.0 [1.3–6.6] | <0.01 | - | - |
|  | *Age 51-70 years* |  |  |  |  |
|  | WMH | 1.6 [1.4–1.8] | <0.001 | - | - |
|  | Lacunes | 1.2 [1.0–1.5] | 0.12 | - | - |
|  | Cortical Infarcts | 0.7 [0.5–1.2] | 0.19 | - | - |
|  | Cerebral atrophy | 2.6 [2.3–2.9] | <0.001 | - | - |
|  | *Age 71-90 years* |  |  |  |  |
|  | WMH | 1.0 [0.9–1.1] | 0.67 | - | - |
|  | Lacunes | 1.1 [1.0–1.2] | 0.31 | - | - |
|  | Cortical Infarcts | 0.7 [0.6–0.9] | <0.01 | - | - |
|  | Cerebral atrophy | 1.2 [1.1–1.3] | <0.001 | - | - |
|  | *Age >= 91 years* |  |  |  |  |
|  | WMH | 1.1 [0.5–2.3] | 0.86 | - | - |
|  | Lacunes | 1.0 [0.4–2.2] | 0.93 | - | - |
|  | Cortical Infarcts | 0.5 [0.1–3.4] | 0.45 | - | - |
|  | Cerebral atrophy | 1.5 [0.7–3.3] | 0.31 | - | - |
|  | **Modality** |  |  |  |  |
|  | WMH | 1.1 [1.0–1.2] | <0.01 | 0.8 [0.7–1.0] | 0.02 |
|  | Lacunes | 1.1 [1.0–1.2] | 0.12 | 0.9 [0.6–1.2] | 0.47 |
|  | Cortical Infarcts | 0.8 [0.6–0.9] | <0.01 | 0.8 [0.4–1.4] | 0.38 |
|  | Cerebral atrophy | 1.4 [1.3–1.5] | <0.001 | 1.4 [1.1–1.6] | <0.001 |
|  | *CT* |  |  |  |  |
|  | WMH | 1.1 [1.0–1.2] | 0.02 | - | - |
|  | Lacunes | 1.1 [1.0–1.2] | 0.19 | - | - |
|  | Cortical Infarcts | 0.7 [0.6–0.9] | <0.01 | - | - |
|  | Cerebral atrophy | 1.3 [1.2–1.4] | <0.001 | - | - |
|  | *MRI* |  |  |  |  |
|  | WMH | 1.0 [0.9–1.2] | 0.83 | - | - |
|  | Lacunes | 1.0 [0.8–1.4] | 0.84 | - | - |
|  | Cortical Infarcts | 0.6 [0.3–1.1] | 0.09 | - | - |
|  | Cerebral atrophy | 2.1 [1.7–2.5] | <0.001 | - | - |
| **Colorectal cancer** | **Sex** |  |  |  |  |
|  | WMH | 1.1 [0.9–1.2] | 0.27 | 0.8 [0.6–0.9] | <0.01 |
|  | Lacunes | 1.0 [0.8–1.2] | 0.99 | 1.1 [0.9–1.4] | 0.45 |
|  | Cortical Infarcts | 1.2 [0.9–1.5] | 0.16 | 0.8 [0.5–1.3] | 0.39 |
|  | Cerebral atrophy | 1.0 [0.9–1.1] | 0.90 | 1.1 [0.9–1.3] | 0.42 |
|  | *Male* |  |  |  |  |
|  | WMH | 1.1 [0.9–1.2] | 0.30 | - | - |
|  | Lacunes | 1.0 [0.9–1.2] | 0.97 | - | - |
|  | Cortical Infarcts | 1.2 [0.9–1.5] | 0.15 | - | - |
|  | Cerebral atrophy | 1.0 [0.9–1.1] | 0.97 | - | - |
|  | *Female* |  |  |  |  |
|  | WMH | 0.8 [0.7–0.9] | <0.01 | - | - |
|  | Lacunes | 1.1 [0.9–1.3] | 0.34 | - | - |
|  | Cortical Infarcts | 1.0 [0.7–1.4] | 0.91 | - | - |
|  | Cerebral atrophy | 1.1 [0.9–1.2] | 0.48 | - | - |
|  | **Age (linear)** |  |  |  |  |
|  | WMH | 2.0 [0.9–4.2] | 0.07 | 1.0 [1.0–1.1] | 0.19 |
|  | Lacunes | 1.6 [0.6–4.3] | 0.39 | 1.0 [1.0–1.0] | 0.43 |
|  | Cortical Infarcts | 2.4 [0.4–13.3] | 0.32 | 1.0 [1.0–1.0] | 0.38 |
|  | Cerebral atrophy | 1.0 [0.5–2.1] | 0.95 | 1.0 [1.0–1.0] | 0.90 |
|  | *Age <= 30 years* |  |  |  |  |
|  | WMH | NA [NA–NA] | NA | - | - |
|  | Lacunes | NA [NA–NA] | NA | - | - |
|  | Cortical Infarcts | NA [NA–NA] | NA | - | - |
|  | Cerebral atrophy | NA [NA–NA] | NA | - | - |
|  | *Age 31-50 years* |  |  |  |  |
|  | WMH | 1.4 [0.5–3.9] | 0.51 | - | - |
|  | Lacunes | 2.0 [0.6–6.9] | 0.25 | - | - |
|  | Cortical Infarcts | 1.6 [0.2–13.1] | 0.64 | - | - |
|  | Cerebral atrophy | 1.3 [0.6–2.9] | 0.57 | - | - |
|  | *Age 51-70 years* |  |  |  |  |
|  | WMH | 1.3 [1.1–1.5] | <0.01 | - | - |
|  | Lacunes | 1.1 [0.8–1.4] | 0.57 | - | - |
|  | Cortical Infarcts | 1.6 [1.0–2.3] | 0.03 | - | - |
|  | Cerebral atrophy | 1.3 [1.1–1.5] | <0.01 | - | - |
|  | *Age 71-90 years* |  |  |  |  |
|  | WMH | 0.9 [0.8–1.0] | 0.19 | - | - |
|  | Lacunes | 1.1 [0.9–1.3] | 0.24 | - | - |
|  | Cortical Infarcts | 1.0 [0.8–1.3] | 0.76 | - | - |
|  | Cerebral atrophy | 1.0 [0.9–1.1] | 0.61 | - | - |
|  | *Age >= 90 years* |  |  |  |  |
|  | WMH | 0.7 [0.4–1.1] | 0.13 | - | - |
|  | Lacunes | 0.5 [0.3–1.2] | 0.12 | - | - |
|  | Cortical Infarcts | 1.1 [0.4–3.1] | 0.86 | - | - |
|  | Cerebral atrophy | 1.1 [0.6–1.9] | 0.77 | - | - |
|  | **Modality** |  |  |  |  |
|  | WMH | 0.9 [0.8–1.0] | 0.11 | 1.1 [0.9–1.4] | 0.26 |
|  | Lacunes | 1.0 [0.9–1.2] | 0.67 | 1.2 [0.8–1.7] | 0.37 |
|  | Cortical Infarcts | 1.2 [0.9–1.4] | 0.19 | 0.8 [0.4–1.5] | 0.49 |
|  | Cerebral atrophy | 1.0 [0.9–1.2] | 0.38 | 0.9 [0.7–1.2] | 0.45 |
|  | *CT* |  |  |  |  |
|  | WMH | 0.9 [0.8–1.0] | 0.07 | - | - |
|  | Lacunes | 1.0 [0.9–1.2] | 0.73 | - | - |
|  | Cortical Infarcts | 1.2 [0.9–1.4] | 0.20 | - | - |
|  | Cerebral atrophy | 1.0 [0.9–1.1] | 0.54 | - | - |
|  | *MRI* |  |  |  |  |
|  | WMH | 1.1 [0.9–1.4] | 0.30 | - | - |
|  | Lacunes | 1.2 [0.9–1.7] | 0.22 | - | - |
|  | Cortical Infarcts | 0.9 [0.5–1.7] | 0.77 | - | - |
|  | Cerebral atrophy | 1.0 [0.8–1.2] | 0.94 | - | - |

**Figure S6. Forest plot summarising event rates, hazard ratios and 95% CIs for each scan phenotype in the main analyses (Baseline), sex-specific models, age-specific models, and modality-specific models, split by outcome: stroke (A), dementia (B), Parkinson’s disease (C), epilepsy (D), and colorectal cancer (E)**

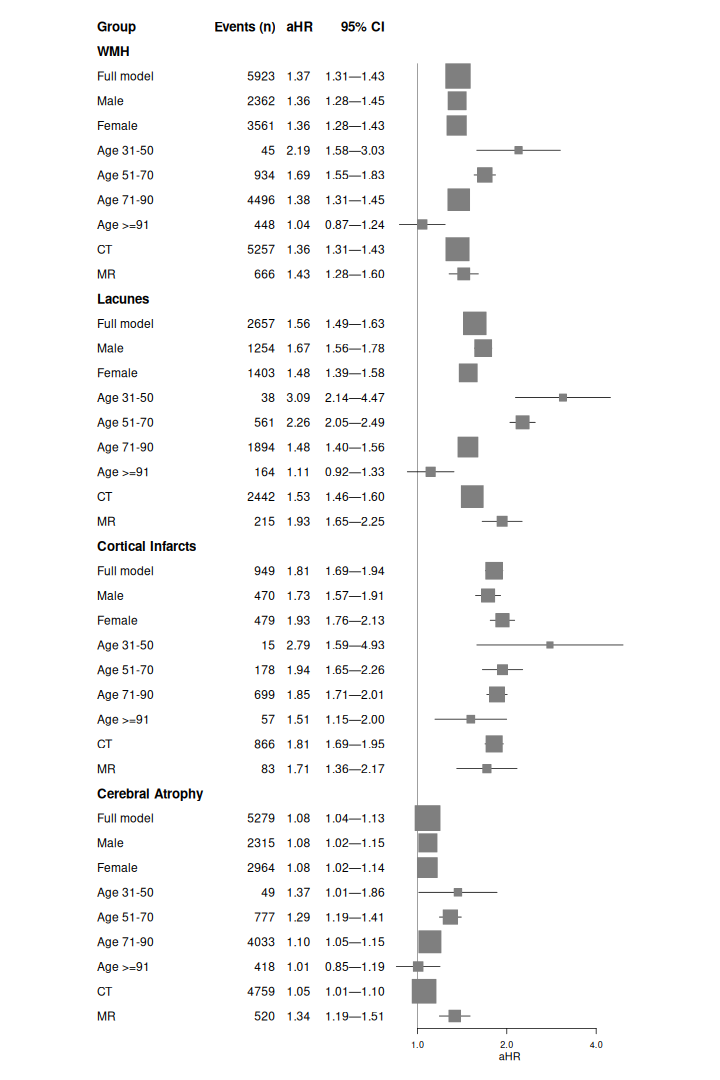

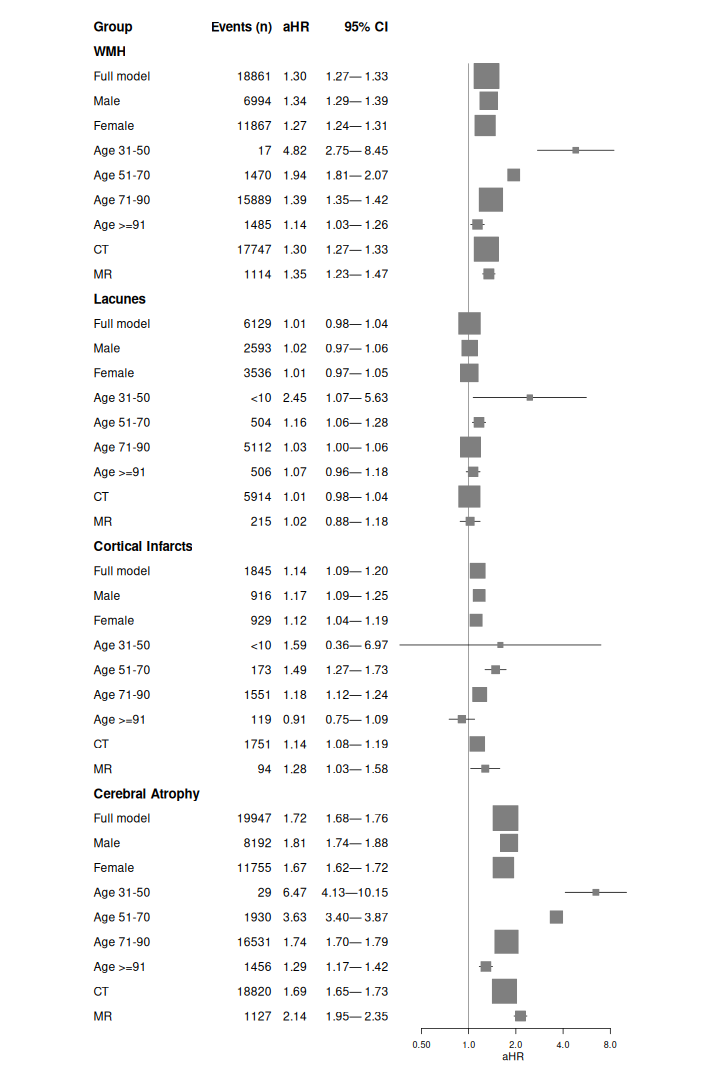

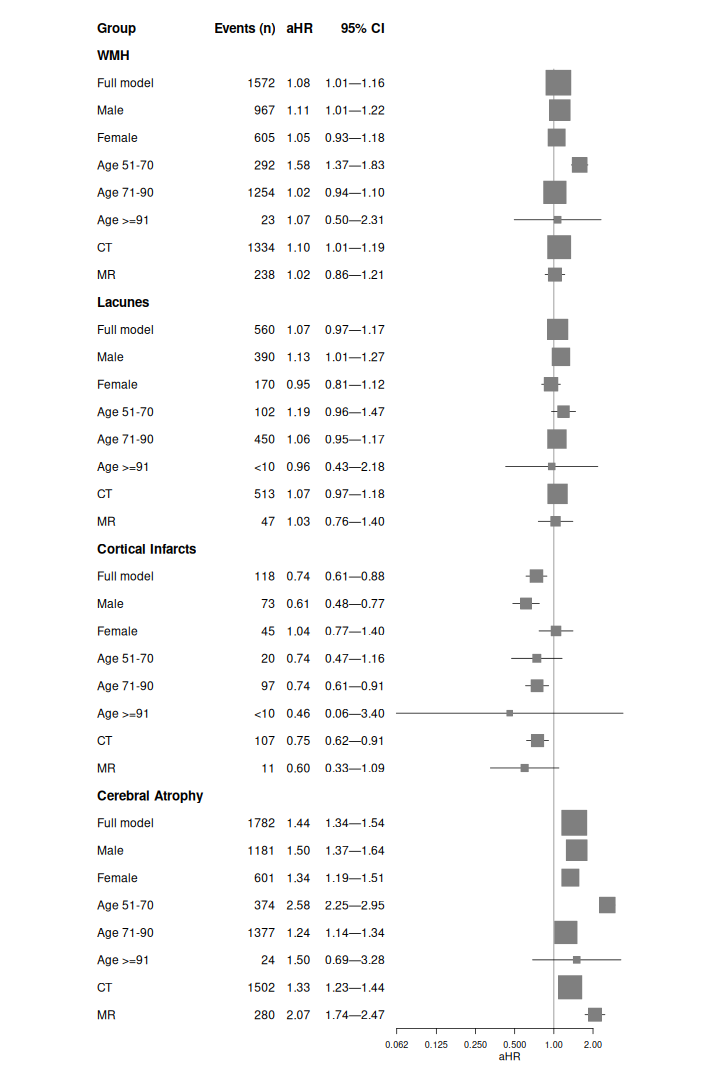

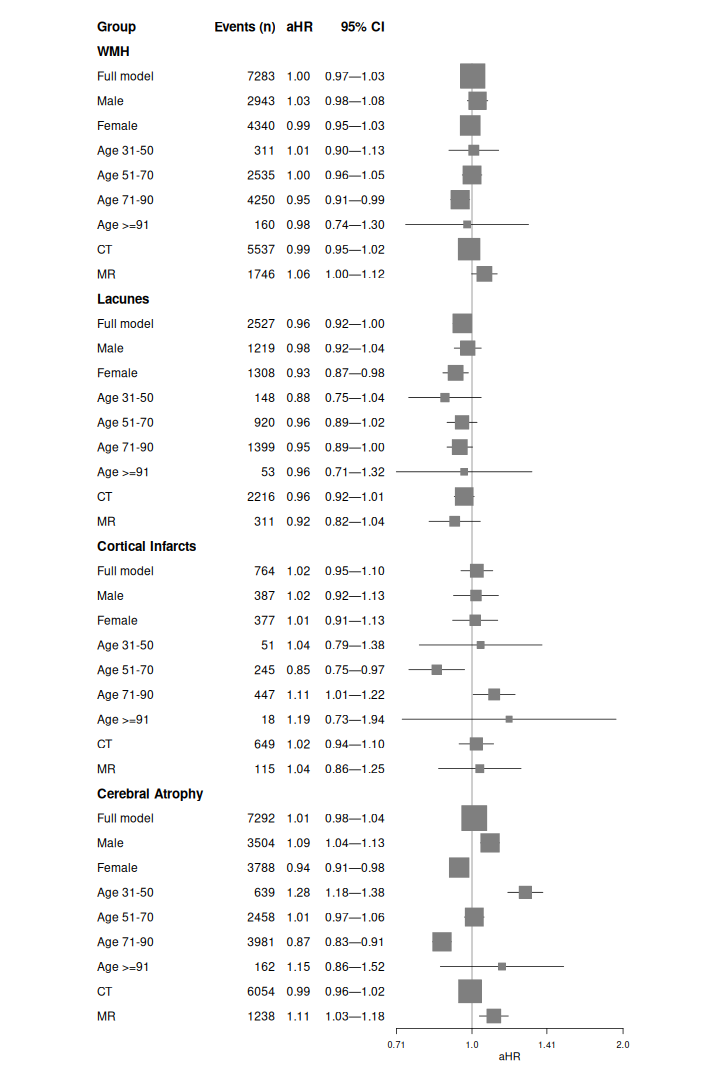

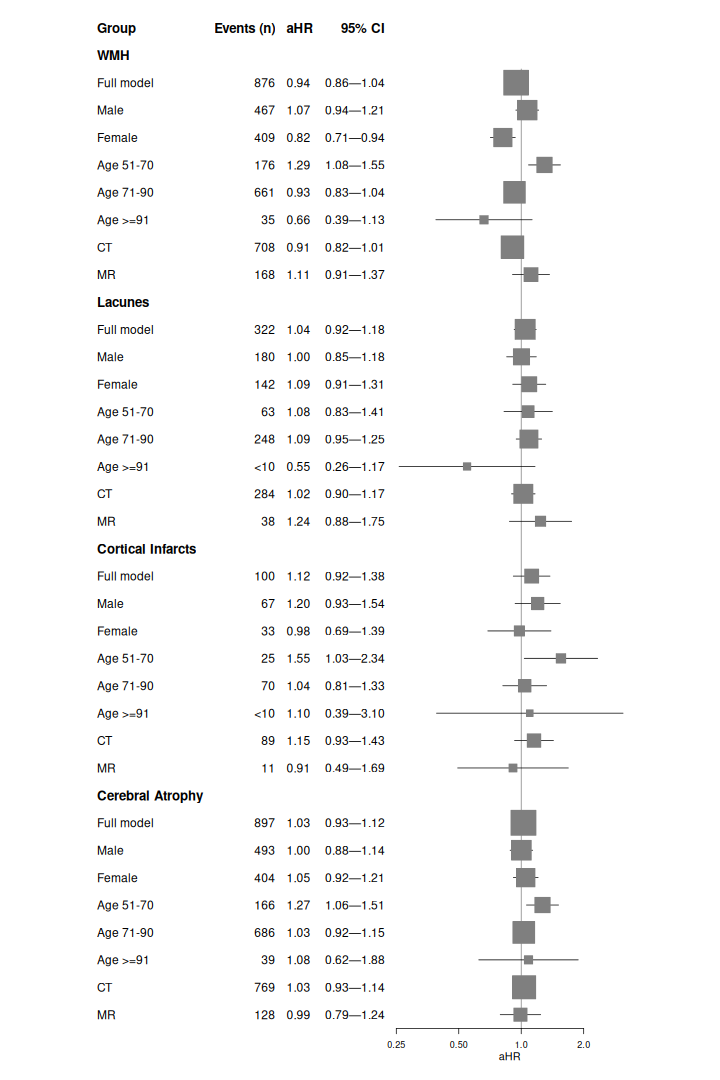

**Figure S7. Forest plot showing the event rate and adjusted hazard ratio (aHR) and 95% CIs per number of exposures (WMH, lacunes, cortical infarcts, and cerebral atrophy) and their association with risk of each outcome across follow-up.**

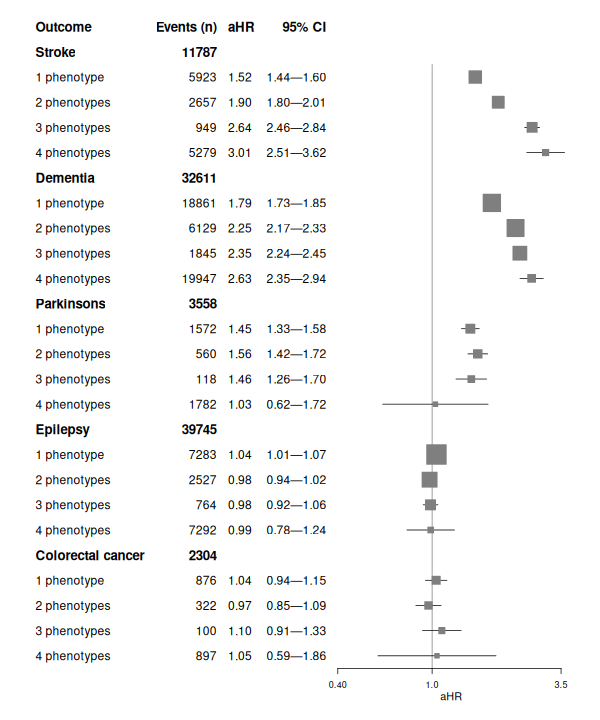

**Figure S8. Forest plot showing the event rate and adjusted hazard ratio (aHR) and 95% CIs per number of exposures (WMH, lacunes, cortical infarcts, and cerebral atrophy) and their association with risk of a) stroke subtypes and b) dementia subtypes across follow-up.**

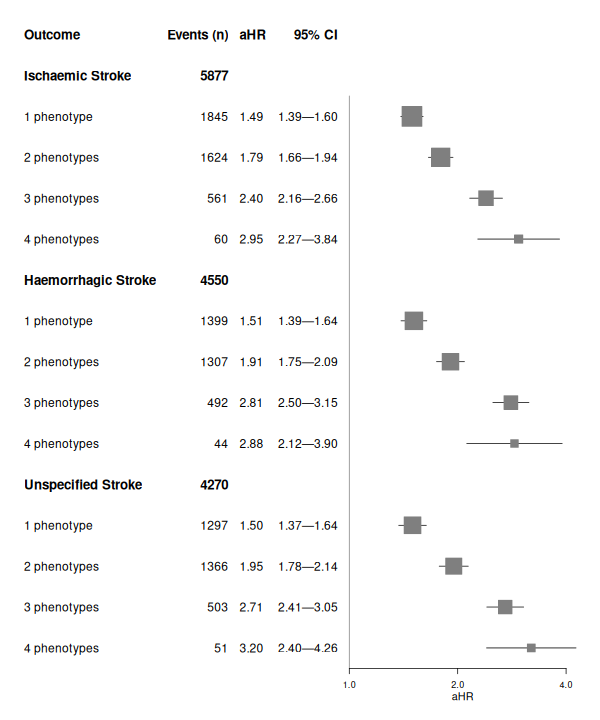

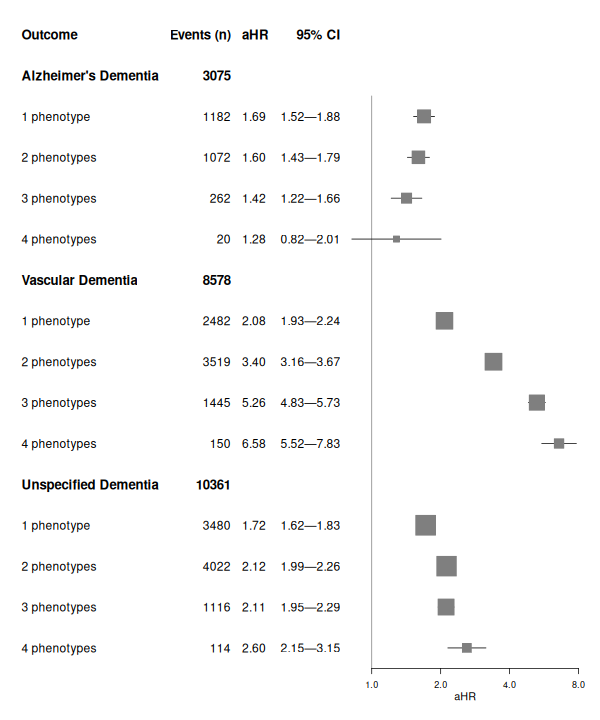
